# Considerations in the Reliability and Fairness Audits of Predictive Models for Advance Care Planning

**DOI:** 10.1101/2022.07.10.22275967

**Authors:** Jonathan Lu, Amelia Sattler, Samantha Wang, Ali Raza Khaki, Alison Callahan, Scott Fleming, Rebecca Fong, Benjamin Ehlert, Ron C. Li, Lisa Shieh, Kavitha Ramchandran, Michael F. Gensheimer, Sarah Chobot, Stephen Pfohl, Siyun Li, Kenny Shum, Nitin Parikh, Priya Desai, Briththa Seevaratnam, Melanie Hanson, Margaret Smith, Yizhe Xu, Arjun Gokhale, Steven Lin, Michael A. Pfeffer, Winifred Teuteberg, Nigam H. Shah

## Abstract

Multiple reporting guidelines for artificial intelligence (AI) models in healthcare recommend that models be audited for reliability and fairness. However, there is a gap of operational guidance for performing reliability and fairness audits in practice.

Following guideline recommendations, we conducted a reliability audit of two models based on model performance and calibration as well as a fairness audit based on summary statistics, subgroup performance and subgroup calibration. We assessed the Epic End-of-Life (EOL) Index model and an internally developed Stanford Hospital Medicine (HM) Advance Care Planning (ACP) model in 3 practice settings: Primary Care, Inpatient Oncology and Hospital Medicine, using clinicians’ answers to the surprise question (“Would you be surprised if [patient X] passed away in [Y years]?”) as a surrogate outcome.

For performance, the models had positive predictive value (PPV) at or above 0.76 in all settings. In Hospital Medicine and Inpatient Oncology, the Stanford HM ACP model had higher sensitivity (0.69, 0.89 respectively) than the EOL model (0.20, 0.27), and better calibration (O/E 1.5, 1.7) than the EOL model (O/E 2.5, 3.0). The Epic EOL model flagged fewer patients (11%, 21% respectively) than the Stanford HM ACP model (38%, 75%). There were no differences in performance and calibration by sex. Both models had lower sensitivity in Hispanic/Latino male patients with Race listed as “Other.”

10 clinicians were surveyed after a presentation summarizing the audit. 10/10 reported that summary statistics, overall performance, and subgroup performance would affect their decision to use the model to guide care; 9/10 said the same for overall and subgroup calibration. The most commonly identified barriers for routinely conducting such reliability and fairness audits were poor demographic data quality and lack of data access. This audit required 115 person-hours across 8-10 months.

Our recommendations for performing reliability and fairness audits include verifying data validity, analyzing model performance on intersectional subgroups, and collecting clinician-patient linkages as necessary for label generation by clinicians. Those responsible for AI models should require such audits before model deployment and mediate between model auditors and impacted stakeholders.

**Contribution to the Field Statement:** Artificial intelligence (AI) models developed from electronic health record (EHR) data can be biased and unreliable. Despite multiple guidelines to improve reporting of model fairness and reliability, adherence is difficult given the gap between what guidelines seek and operational feasibility of such reporting. We try to bridge this gap by describing a reliability and fairness audit of AI models that were considered for use to support team-based advance care planning (ACP) in three practice settings: Primary Care, Inpatient Oncology, and Hospital Medicine. We lay out the data gathering processes as well as the design of the reliability and fairness audit, and present results of the audit and decision maker survey. We discuss key lessons learned, how long the audit took to perform, requirements regarding stakeholder relationships and data access, and limitations of the data. Our work may support others in implementing routine reliability and fairness audits of models prior to deployment into a practice setting.

## Introduction

Concern about the reliability and fairness of deployed artificial intelligence (AI) models trained on electronic health record (EHR) data is growing. EHR-based AI models have been found to be unreliable, with decreased performance and calibration across different geographic locations and over time; for example, an Epic sepsis prediction algorithm had reduced performance when validated by University of Michigan researchers (1) and acute kidney injury models have shown worsening calibration over time (2). AI models have also been found to be unfair, with worse performance and calibration for historically marginalized subgroups; for example, widely used facial recognition algorithms have lower performance on darker-skinned females (3); and widely used health insurance algorithms underrate the disease status of Black patients compared with similar White patients (4). Despite lacking evidence of reliability and fairness, algorithms are still being deployed (5).

To promote improved reliability and fairness of deployed EHR models, at least 15 different model reporting guidelines have been published (6–20). Some commonly included items related to reliability in these guidelines include external validation (6,8–10,14–17,19); multiple performance metrics such as Area Under Receiver Operating Curve (AUROC) (6,8–12,14–18), positive predictive value (PPV) (9–12,14,16–18), sensitivity (8–12,14,16–18), and specificity (8–12,14,17,18); confidence intervals or another measure of variability of the performance (6,8–12,15,18–20); and calibration plots (6,8–10,12,14). Some commonly included items related to fairness include summary statistics (10,11,15,17,18,20), like the distribution of demographics such as sex (11,15,17,20) and race/ethnicity (15,17,20), as well as subgroup analyses that investigate how a model performs for specific subpopulations (7,9,11–13,15,18,20). Nevertheless, many of these items are infrequently reported for both published (21) and deployed EHR models (22).

Several efforts seek to address this reporting gap. For example, there is an existing auditing framework that supports AI system development end-to-end and links development decisions to organizational values/principles (23). There is also currently an open-source effort to better understand, standardize and implement algorithmic audits (24).

In this work, we illustrate a reliability/fairness audit of 12-month mortality models considered for use in supporting team-based ACP in three practice settings (Primary Care, Inpatient Oncology, Hospital Medicine) at a quaternary academic medical center in the United States (25–27) (Figure 1). We 1) design and report a reliability/fairness audit of the models following existing reporting guidelines, 2) survey decision makers about how the results impacted their decision of whether to use the model, and 3) quantify the time, workflow and data requirements for performing this audit. We discuss key drivers and barriers to making these audits standard practice. We believe this may aid other decision makers and informaticists in operationalizing regular reliability and fairness audits (22, 23).

**Figure 1:**
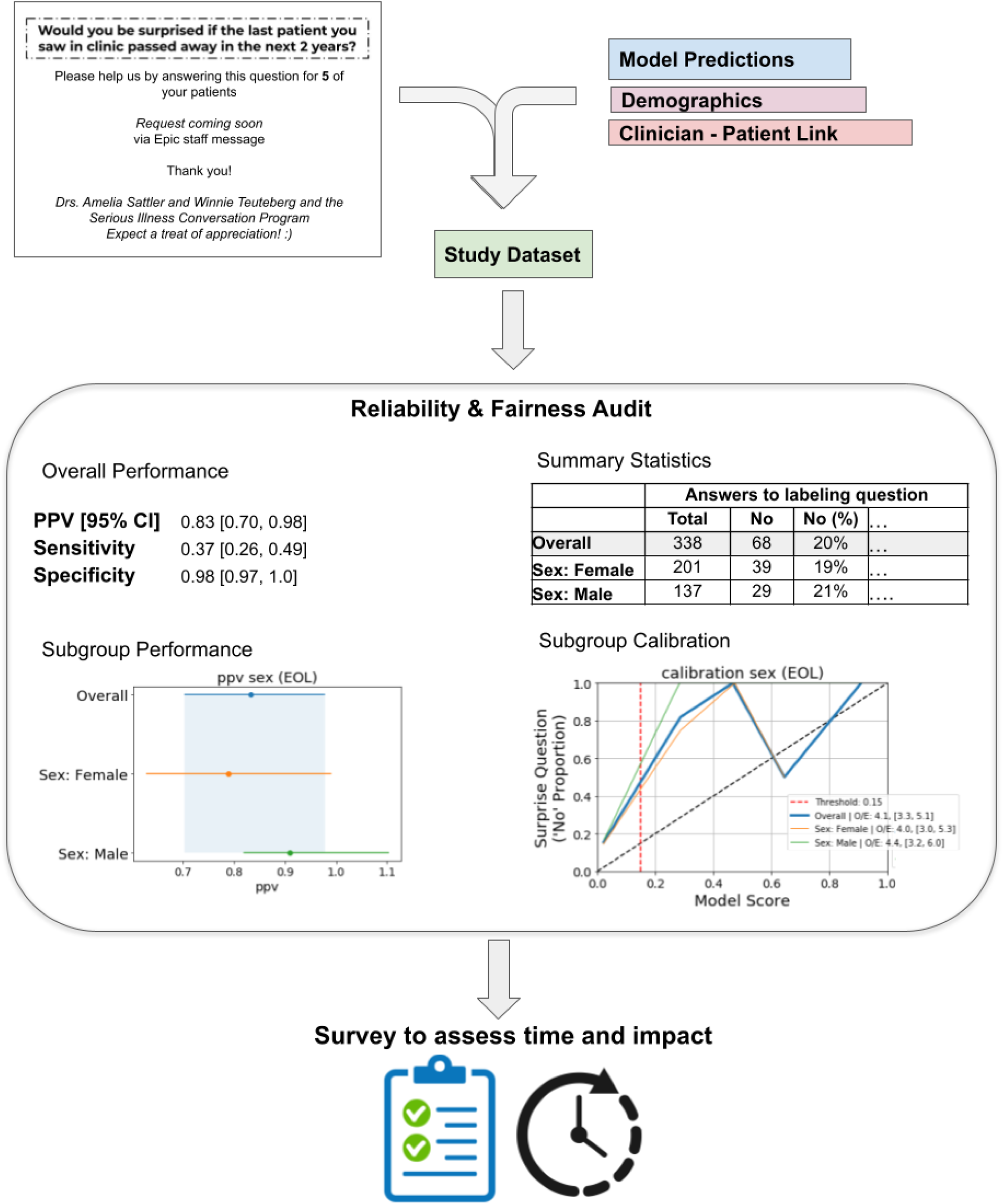
Overview of Audit Process. Results and plots listed are for the Epic End-of-life Index Low Threshold for Primary Care. The “labeling question” under Summary Statistics is “Would you be surprised if this patient passed away in 2 years?”

*Note: we use recorded race/ethnicity in the EHR as a way to measure how models may perform across such groupings, as recommended* (*15, 21*)*. Importantly, race/ethnicity is not used as an input for any of the models and we do not use it as a “risk factor” for health disparities* (*28–30*)*. We recognize race/ethnicity has widely varying definitions* (*31*) *and is more a social construct* (*32*) *than a biological category* (*30*)*. We also caution that studies have found poor concordance of race/ethnicity data as recorded in the EHR with the patient’s self-identification* (*33*) (*34*)*. However, performance by race/ethnicity subgroups is a recommended analysis in reporting guidelines*.

### Background on Advance Care Planning and Model Usage

Much of care for patients at the end of their lives is not goal-concordant, i.e. not consistent with the patients’ goals and values. For example, a survey (35) of Californians’ attitudes towards death and dying found that 70% would prefer to die at home. Despite this, only 30% of all deaths happened at home in 2009. Meanwhile 60% occurred in a hospital or nursing home (26).

In 2018 the Stanford Department of Medicine began implementation of Ariadne Labs’ Serious Illness Care Program (SICP) (36) to promote goal-concordant care by improving timing and quality of advance care planning conversations. By following best practices (37), the Stanford SICP trained and supported clinicians in using the structured Serious Illness Conversation Guide (SICG) in their practice.

Through the duration of this audit, *Primary Care* and *Inpatient Oncology* were developing implementation plans, while *Hospital Medicine* had an active implementation after SICG training of key physicians and staff members using a 12-month mortality model to generate patient prognoses that were shared with the entire clinical team (25). Two models were considered: 1) the 12-month mortality model which runs only on currently hospitalized patients only and is currently used by the Hospital Medicine SICP team (HM ACP), and 2) the Epic End-of-life (EOL) Index, which unlike HM ACP, runs for all patients receiving care in the health system, not just hospitalized patients.

We assessed these models by performing a reliability audit (model performance and calibration) and fairness audit (summary statistics, subgroup performance, subgroup calibration) to ascertain whether the Epic EOL Index appropriately prioritize patients for ACP in *Primary Care*, and which of the two models appropriately prioritizes patients for ACP in *Inpatient Oncology* and *Hospital Medicine*.

## Methods

We first provide details on the two models and then summarize the processes required to complete the fairness and reliability audit. We describe the metrics that comprised the quantitative aspect of the audit. We then describe the methods we used to identify and gather the data needed to complete the audit, including calculating the minimum sample size of ground truth labels required for model evaluation, obtaining those ground truth labels by clinician review, and merging those labels with patient records to create the audit dataset. Lastly, we describe the methods used to compute the audit metrics, and how we presented the results of the audit to clinicians to obtain feedback.

### AI Models

We audited two models currently deployed at Stanford Health Care: the Epic EOL Index model and Stanford HM ACP model (Table 1).

**Table 1:** Model Information for each setting.

|  |  |  |  |  |  |
| --- | --- | --- | --- | --- | --- |
| **Setting** | **Primary Care** | **Inpatient Oncology** | **Inpatient Oncology** | **Hospital Medicine** | **Hospital Medicine** |
| **Model** | Epic EOL - Low Threshold | Epic EOL - High Threshold | Stanford HM ACP | Epic EOL - High Threshold | Stanford HM ACP |
| **Features** | Demographics (Age, Sex, Insurance status), Labs (Albumin, RDW), Comorbidities (Cancer, Neuro., Psych., ....Cardio., Resp., . ....), Medications (many) | Demographics (Age, Sex, Insurance status), Labs (Albumin, RDW), Comorbidities (Cancer, Neuro., Psych., ....Cardio., Resp., . ....), Medications (many) | Demographics (Age, Sex), Lab/Procedure Orders (done in the last year) | Demographics (Age, Sex, Insurance status), Labs (Albumin, RDW), Comorbidities (Cancer, Neuro., Psych., ....Cardio., Resp., . ....), Medications (many) | Demographics (Age, Sex), Lab/Procedure Orders (done in the last year) |
| **# Features** | 46 | 46 | 13189 | 46 | 13189 |
| **Model Type** | Logistic Regression | Logistic Regression | Gradient boosted Tree | Logistic Regression | Gradient boosted Tree |
| **Output** | One-year Mortality Risk | One-year Mortality Risk | One-year Mortality Risk | One-year Mortality Risk | One-year Mortality Risk |
| **Predictions Available For:** | All adult patients within health system | All adult patients within health system | All currently hospitalized adult patients | All adult patients within health system | All currently hospitalized adult patients |
| **Threshold** | 0.15 (Low) | 0.45 (High) | 0.25 (HM Implementation Threshold) | 0.45 (High) | 0.25 (HM Implementation Threshold) |
| **Source of Model Information** | Epic Cognitive Computing Model Brief: End of Life Index (Galaxy, PDF) | Epic Cognitive Computing Model Brief: End of Life Index (Galaxy, PDF) | AI ACP Technical Details | Epic Cognitive Computing Model Brief: End of Life Index (Galaxy, PDF) | AI ACP Technical Details |
| **Time of Model Predictions** | 11/16/2021 | 6/14/2021 | 8/15/2021 - 3/19/2022 | 1/31/2022 | 2/21/2022, 2/23/2022, 3/1/2022, 3/4/2022, 3/7/2022, 3/14/2022, 3/21/2022 |
| **Notes on Time of Model Predictions** | Daily predictions are performed, but were not available to be extracted or retrospectively pulled, so we only used a one-time pull on 11/16/2021 | Daily predictions are performed, but were not available to be extracted or retrospectively pulled, so we only used a one-time pull on 6/14/2021 | Daily predictions were performed and the most recent model prediction on or before the date of the clinician label was used. | Daily predictions are performed, but were not available to be extracted or retrospectively pulled, so we only used a one-time pull on 1/31/2022 | Daily predictions were performed and were stored before sending out email requesting clinicians to label. |
| **Location of Model Predictions** | Box Folder: Epic EoL Index Validation | Box Folder: Epic EoL Index Validation | shahlab secure server: /data4/AI-ACP/predictions/ngb_hist | Box Folder: Epic EoL Index Validation | shahlab secure server: /data4/AI-ACP/predictions/ngb_hist |

The Epic EOL Index model (38) is a logistic regression model that predicts risk of 12-month mortality (Table 1). It takes in 46 input features including demographics (e.g., age, sex, insurance status), labs (e.g., albumin, RDW), comorbidities (e.g., such as those relating to cancer, neurological diagnoses, cardiologic diagnoses, and more), and medications. While organizations using the Epic EHR software are able to set any threshold for converting the model output into a flag to indicate an action is recommended, two thresholds are pre-specified by Epic: a *low threshold* of 0.15 selected based on sensitivity (38), and a *high threshold* of 0.45 selected based on positive predictive value (38). We decided to audit the Epic EOL Index with the low threshold in Primary Care (given lower patient acuity) and with the high threshold in Inpatient Oncology and Hospital Medicine. We retrieved scores on November 16 2021 for Primary Care, June 14 2021 for Inpatient Oncology and January 31 2022 for Hospital Medicine.

The Stanford HM ACP model is a gradient boosted tree model (39) that predicts risk of 3-12 month mortality (Table 1). It takes 13,189 input features including demographics (e.g., age, sex), lab orders (e.g., complete blood count with differential, arterial blood gas) and procedure orders (e.g., ventilation, respiratory nebulizer) for all hospitalizations within the last year and is run daily on patients admitted to the Hospital. Patients with a model output probability above 0.25 are flagged in a “Recommended for Advance Care Planning” column in Epic available to all clinicians at Stanford (25, 26). On a retrospective cohort involving 5965 patients with 12-month mortality labels (prevalence of 24%), this model flagged 23% of patients and had a PPV of 61% (25). For Inpatient Oncology and Hospital Medicine, we retrieved scores for patients on the day of the clinician’s label for that patient.

### Audit metrics

In previous work (22) we synthesized items that were suggested for reporting by model reporting guidelines to identify the most relevant items for reliability and fairness.

To quantify model reliability, we computed sensitivity, specificity and PPV as these estimate a model’s diagnostic capabilities. We computed 95% confidence intervals for each of these metrics using the empirical bootstrap (40) with 1000 bootstrap samples. We also assessed model calibration using calibration plots and the Observed events/Expected events (O/E) ratio (see details below in the section titled Performing the Audit).

To quantify model fairness, we computed summary statistics across subgroups, defined by sex, race/ethnicity, and age as well as the intersection of race/ethnicity and sex. We also evaluated the model’s performance metrics and calibration in each of these subgroups (see details below in the section titled Performing the Audit).

### Gathering the data required for the audit

#### Sample Size Calculation

We calculated a minimum necessary sample size for external validation of the two prediction models, based on a desired level of calibration (41). We measured calibration as O/E and used the delta method for computing a confidence interval for O/E (41). Assuming a perfect O/E value being 1.0, we aimed for a 95% confidence interval width of [0.74, 1.34]. Based on clinician feedback, in Primary Care, we assumed a 20% prevalence of the positive label; in Inpatient Oncology, we assumed a 70% prevalence of the positive label. In Hospital Medicine, we assumed a 40% prevalence of the positive label.

#### Obtaining Ground Truth Labels

We used a validated instrument, the *surprise question* (42), to assign ground truth labels for patients. The surprise question asks “Would you be surprised if [patient X] passed away in [Y years]?” An answer of “no” to the surprise question for a given patient constitutes a positive label (for example, if the treating physician would not be surprised if a patient died in 1 year, we assume that the patient is at high risk of dying and should be labeled as “recommended for advance care planning”). A recent meta-analysis (43) found that among 16 studies, the 6-to-12-month surprise question’s sensitivity (using records of 12-month mortality as ground truth) ranged from 12% to 93%; specificity ranged from 14% to 98%, PPV ranged from 15% to 79%, and c-statistic ranged from 0.51 to 0.82. In other words, we used the answer to the surprise question as a *proxy* for Y-year mortality in our patient population, because waiting the Y years to ascertain whether patients passed away would have greatly extended the timeframe required to complete the audit. Our audit thus assessed model performance based on concordance of model predictions with clinician-generated assessments of patient mortality via the surprise question.

We specified Y = 1 year for the surprise question for Inpatient Oncology and Hospital Medicine patients and Y = 2 years for the Primary Care setting, given lower acuity of patients in Primary Care clinics (Table 2).

**Table 2:**
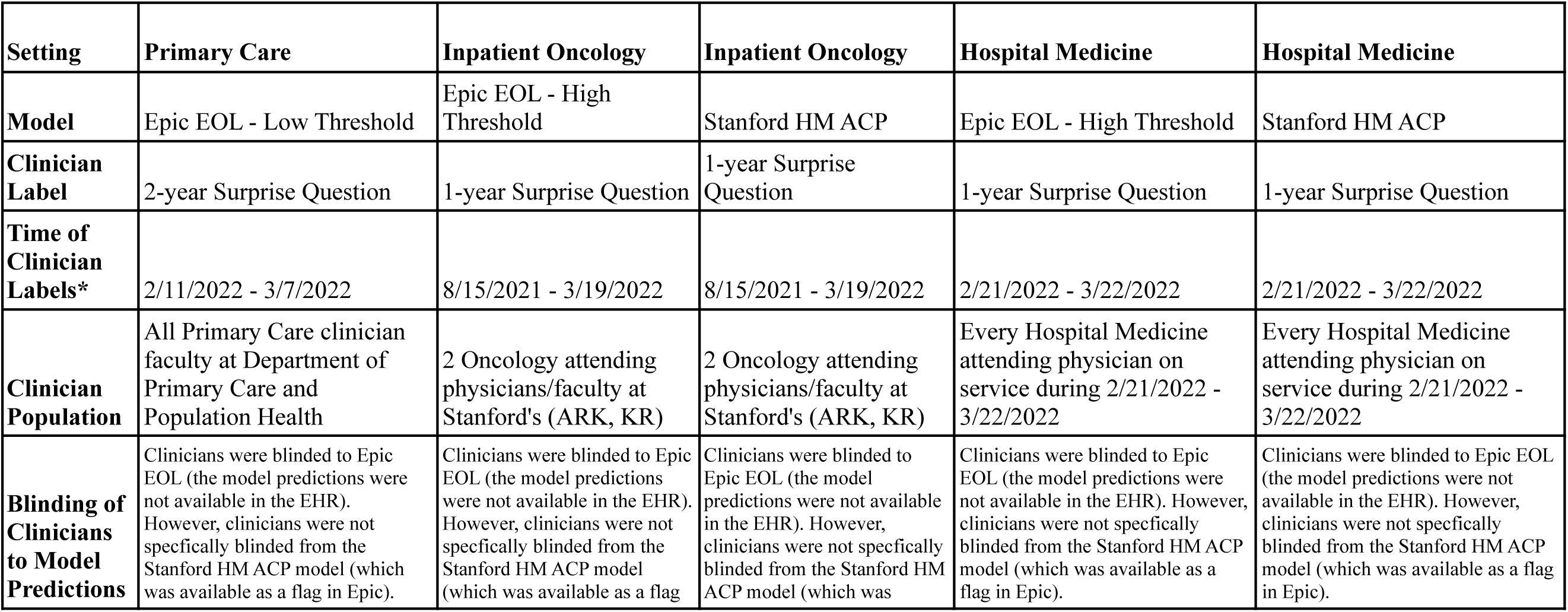
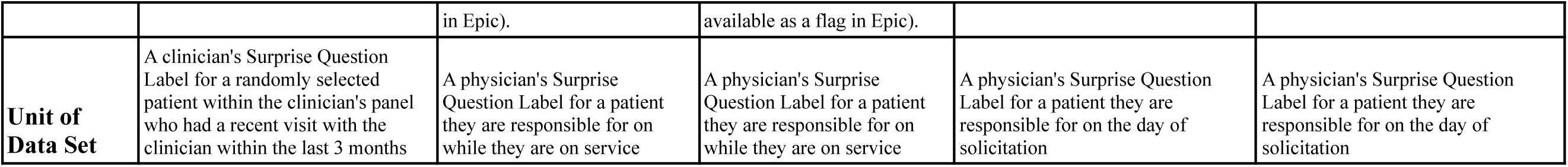
Clinician Label Information.

To obtain answers to the surprise question for Primary Care patients, we first selected from patients who had a visit with a provider between October 7 2021 and January 7 2022. We then randomly sampled 5 unique patients to generate a list for each provider; if there were fewer than 5 unique patients, all patients were kept in the provider’s list. We then sent personalized messages using our EHR’s messaging system to each provider asking them to answer the surprise question for each randomly selected patient (Table 3, Supplemental Figure 1). For Hospital Medicine, we identified providers who were on service between February 21 2022 and March 21 2022, and sent them a message once a week during that period requesting them to answer the surprise question for the patients they had been responsible for during their shifts in that period (Table 3, Supplemental Figure 2). For both Primary Care and Hospital Medicine, we incentivize providers to answer the surprise question by offering chocolates to those who received the message. For Inpatient Oncology we selected patients who were seen by either co-author ARK or KR between August 15 2021 and March 19 2022. ARK and KR answered the 1-year surprise question for all patients they were responsible for while on hospital service during that period (Table 2).

**Table 3:** Solicitation of Clinician Labels.

| Setting | Primary Care | Inpatient Oncology | Inpatient Oncology | Hospital Medicine | Hospital Medicine |
| --- | --- | --- | --- | --- | --- |
| Model | Epic EOL - Low Threshold | Epic EOL - High Threshold | Stanford HM ACP | Epic EOL - High Threshold | Stanford HM ACP |
| Sample Size Required to achieve calibration 95% O/E CI of [0.74, 1.34] (assuming true O/E = 1) | 176 assuming prevalence of 20% | 19 assuming prevalence of 70% | 19 assuming prevalence of 70% | 66 assuming prevalence of 40% | 66 assuming prevalence of 40% |
| Solicitation of Clinician Labels | Epic Staff Message sent 2/11/2022 | N/A (Physician answered surprise question for all patients responsible for each morning on service) | N/A (Physician answered surprise question for all patients each morning on service) | Secure Emails sent 2/21/2022, 2/23/2022, 3/1/2022, 3/4/2022, 3/7/2022, 3/14/2022, 3/21/2022 | Secure Emails sent 2/21/2022, 2/23/2022, 3/1/2022, 3/4/2022, 3/7/2022, 3/14/2022, 3/21/2022 |
| Generation of Solicitations | 1. Link visits at a primary care visit site since 09/2021 with patient demographics<br>2. Filter to visits after 10/72/2021<br>3. For each provider: filter to visits with the provider that were with patients within their panel<br>4. Remove visits for providers on days where that provider had more than 30 visits (assume this is artifact of data base)<br>5. Randomly sample 5 patients of remaining | N/A | N/A | 1. For each attending physician on service, generate an email asking them to answer the surprise question for all patients they are responsible for that day | 1. For each attending physician on service, generate an email asking them to answer the surprise question for all patients they are responsible for that day |
| Example Solicitation | Link | N/A | N/A | Link | Link |
| Announcement of Solicitation | Slide in Division Meeting | N/A | N/A | Email at week start | Email at week start |
| Incentive with Solicitation | Bag of Ghirardelli Chocolates personally addressed, thanking for answering the surprise question | N/A | N/A | Bag of Ghirardelli Chocolates personally addressed, thanking for answering the surprise question | Bag of Ghirardelli Chocolates personally addressed, thanking for answering the surprise question |
| Location of Code to Generate Solicitations | shahlab secure server: /data4/jhlu/EOL/[2022-02-01 using concept] pcph_merge_visits_generate_validation_lists_and_plausibility_lists.ipynb | N/A | N/A | shahlab secure server: /data4/jhlu/hm-surprise-gathering/PROD | shahlab secure server: /data4/jhlu/hm-surprise-gathering/PROD |
| # Clinicians Solicited | 79 | N/A | N/A | 22 | 22 |
| Size of Solicitations | 386 | N/A | N/A | 545 | 545 |

Note that the physicians were blinded to Epic EOL Index model predictions, but they were not blinded to the Stanford HM ACP Flag as the flag was available in Epic and in active use at the time of the audit. Co-author ARK reported occasionally referencing the flag when answering the surprise question for patients with rarer cancers. While we recognize this biases our results in favor of the Stanford HM ACP model, we also did not have the ability to suppress the flag just for those clinicians.

#### Creating the Audit Data Set

Each patient’s surprise question ground truth labels were linked with their corresponding patient records from our clinical data warehouse (44), which included patient demographics (sex, date of birth, race, ethnicity), and with the two models’ output predictions (Figure 1).

We excluded all patients where their provider had not answered the surprise question during the response period. For Inpatient Oncology, we also excluded all patients for which a medical record number was not available. The number of patients excluded for these reasons are provided in the Results.

Finally, we converted patient demographic data into one-hot encoded columns. For sex, we assigned this value based on biological sex (45) (and did one-hot encodings of the potential values). For age, we computed the patient’s age at the time of the clinician’s surprise question assessment by subtracting their date of birth; we then generated age subgroups by decade of life e.g. (10,20], (20, 30] etc. For ethnicity/race, we pulled the ethnicity variable and the race variable, both based on Office of Management and Budget variables (46). We then performed one-hot encoding of the ethnicity and race variables separately, and used a logical AND to generate the ethnicity/race variable: e.g. a Hispanic or Latino, White patient. Lastly, for ethnicity/race and sex, we created intersectional combinations using a logical AND to identify all observed permutations of these variables.

### Performing the Audit

After we generated the audit data set, we first computed summary statistics. Specifically, for each demographic variable (sex, age, ethnicity/race, and the intersection of ethnicity/race and sex), we computed the counts of each subgroup within that demographic, as well as the % of the count within the entire data set, and the number and % of positive ground truth labels. We also computed a 95% confidence interval on the positive ground truth label prevalence in each subgroup, using the Clopper-Pearson interval (47) and determined if it overlapped with the confidence interval of the overall positive label prevalence; this evaluated whether ground truth labels were consistent across different demographic subgroups.

We next evaluated model performance. With the ground truth labels and model flags, we computed the following metrics: number of flagged patients, PPV, sensitivity, and specificity. We computed 95% confidence intervals on the performance metrics using the empirical bootstrap: we generated 1000 bootstrap samples of the data set. For each sample, we computed the performance metrics, and computed the difference between each metric from the bootstrap sample and that from the overall study group. (Note the metric on the bootstrap sample may have been null due to dividing by zero, e.g. for PPV if there were no patients that were flagged by the model) We used these differences to generate a distribution of 1000 bootstrap differences, computed the 2.5^th^ and 97.5^th^ percentiles of the differences (excluding null values), and subtracted these from each metric to generate the empirical bootstrap confidence interval for each metric.

We also evaluated model performance for the subgroups defined by the demographic variables above by computing PPV, sensitivity, and specificity. We computed 95% confidence intervals for each subgroup as above, replacing “overall study group” with the subgroup. We then check if the confidence intervals overlap. Note that resulting confidence intervals had values in some cases that were above 1 or below 0, due to large differences resulting from wide variation in the metric over the bootstrap sampling (40).

We evaluated the models’ calibration using calibration plots. A calibration plot provides a visual assessment of how well predicted risk probabilities are aligned with observed outcomes. To generate the calibration plots, we grouped predicted probabilities into quintiles, and within each quintile, computed the average of the predicted risks. We then plotted the averaged predicted risk for each quintile on the x-axis and proportion of positive ground truth labels for each quintile on the y-axis (6,8–10,12,14). We also computed the Observed events/Expected events ratio O/E, which measures the overall calibration of risk predictions, which is computed as the ratio of the total number of observed to predicted events. We computed O/E by dividing the total number of positive ground truth labels by the sum of model output probabilities and used the delta method for computing a 95% confidence interval on O/E (50). The ideal value for O/E is 1; a value < 1 or > 1 implies that the model over or under predicts the number of events, respectively (41).

We evaluated subgroup calibration by generating calibration plots and by computing the O/E for each subgroup, again using the delta method to compute a 95% confidence interval on O/E (50). Note: because this method’s standard error formula for ln(O/E) has O in the denominator, the interval is undefined if O = 0.

### Presenting Audit Results to Decision Makers

We presented the results of our audit to decision makers in Primary Care (co-authors AS, WT), Inpatient Oncology (co-authors ARK, WT, SC, KR, MG), and Hospital Medicine (co-authors SW, LS, RL), in a separate presentation for each setting. Each presentation first gave context to the audit, including sharing previous findings that AI models have been unreliable (5, 48) or unfair (4), as well as that race/ethnicity data in the EHR is known to have inaccuracies (33). Then, we shared the summary statistics, model performance, model calibration, subgroup performance and subgroup calibration.

We also designed a survey for the decision makers to complete at the end of each presentation (<u>Supplemental Methods</u>). In the survey, we assessed their understanding of reliability/fairness by asking “What does it mean to you for a model to be reliable/fair?” and “What are the first thoughts that came to your mind on seeing the results of the reliability and fairness audit?” We also assessed whether specific components of the reliability/fairness audit would or would not affect decision making, and asked if there would be any other information they believe should be included in the audit. Example surveys were shared with several decision makers (co-authors WT, SW, AS), informaticists (co-authors AG, AC) and the director of operations of an AI research & implementation team (co-author MS) for feedback prior to giving the survey.

After we received the survey responses, we reviewed and summarized the most common structured responses. We also read the free text responses, identified themes (ensuring that every response had at least one theme represented) and categorized responses by the themes. JL was the sole coder, and performed inductive thematic analysis to generate codes.

## Results

### Reliability And Fairness Audit

We report the reliability and fairness audits below. For simplicity, all confidence intervals are listed in the Tables. Also, only statistically significant results are listed in the Tables; full results including those without statistically significant differences are listed in the Supplemental Tables.

#### Primary Care

We calculated we would need a sample size of 176 to achieve an O/E 95% confidence interval of [0.74, 1.34], assuming a 20% prevalence of the positive label. We solicited 79 clinicians for 386 labels of their patients (2-year surprise question answers). 70 clinicians responded with 344 labels (89% response rate). Six of the response labels were “Y/N” or “DECEASED” and were filtered out, leaving 338 labels fitting the schema.

##### Epic EOL Low Threshold in Primary Care

The final data set size for the Epic EOL - Low Threshold model in Primary Care was 338 with 68 positive labels after we linked the 338 clinician labels fitting the schema with Epic EOL model predictions and patient demographics (Table 4).

**Table 4:**
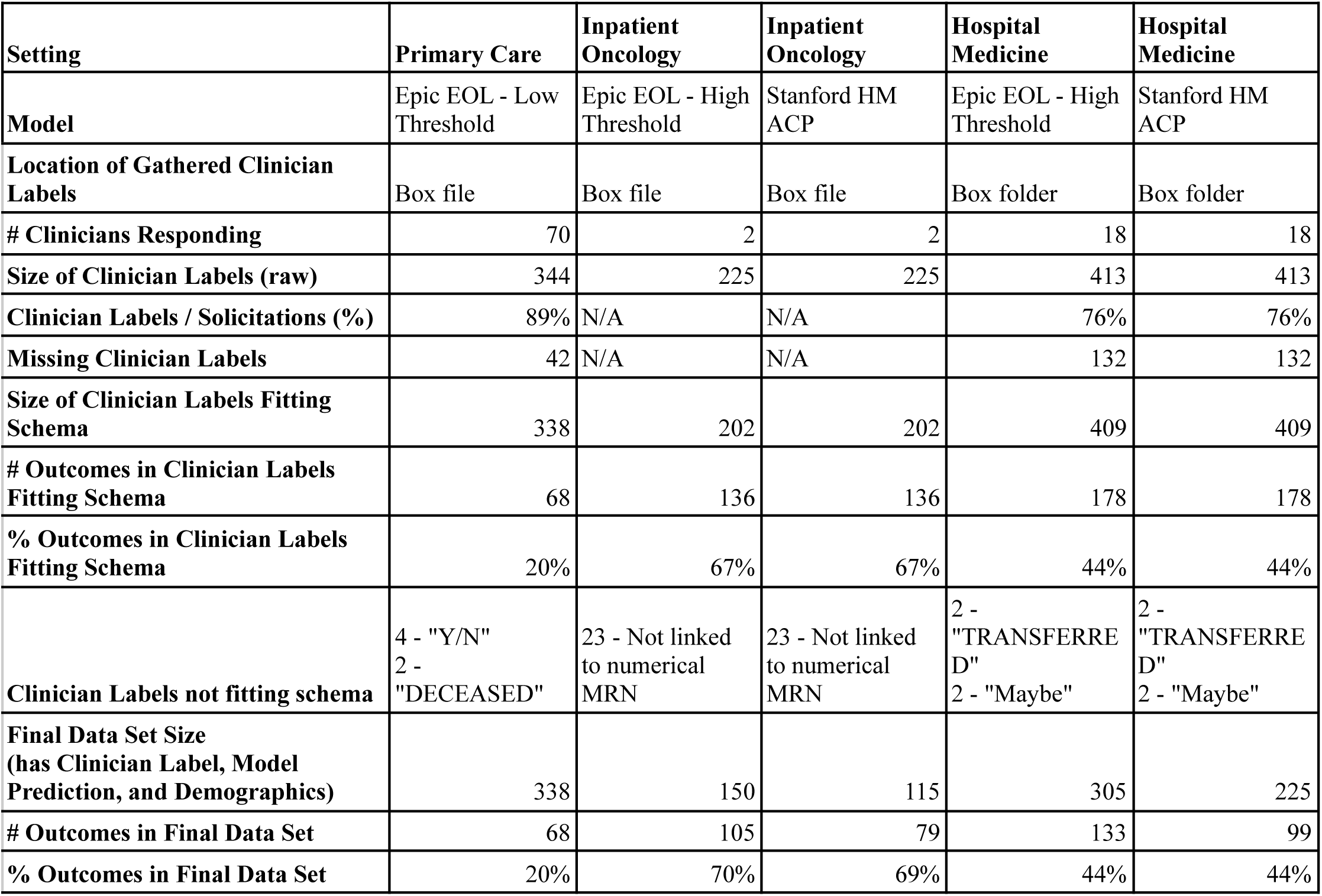
Processing and Final Data Sets.

| **Setting** | **Primary Care** | **Inpatient Oncology** | **Inpatient Oncology** | **Hospital Medicine** | **Hospital Medicine** |
| --- | --- | --- | --- | --- | --- |
| **Model** | Epic EOL - Low Threshold | Epic EOL - High Threshold | Stanford HM ACP | Epic EOL - High Threshold | Stanford HM ACP |
| **Location of Gathered Clinician Labels** | Box file | Box file | Box file | Box folder | Box folder |
| **# Clinicians Responding** | 70 | 2 | 2 | 18 | 18 |
| **Size of Clinician Labels (raw)** | 344 | 225 | 225 | 413 | 413 |
| **Clinician Labels / Solicitations (%)** | 89% | N/A | N/A | 76% | 76% |
| **Missing Clinician Labels** | 42 | N/A | N/A | 132 | 132 |
| **Size of Clinician Labels Fitting Schema** | 338 | 202 | 202 | 409 | 409 |
| **# Outcomes in Clinician Labels Fitting Schema** | 68 | 136 | 136 | 178 | 178 |
| **% Outcomes in Clinician Labels Fitting Schema** | 20% | 67% | 67% | 44% | 44% |
| **Clinician Labels not fitting schema** | 4 - "Y/N"<br>2 - "DECEASED" | 23 - Not linked to numerical MRN | 23 - Not linked to numerical MRN | 2 - "TRANSFERRED"<br>2 - "Maybe" | 2 - "TRANSFERRED"<br>2 - "Maybe" |
| **Final Data Set Size (has Clinician Label, Model Prediction, and Demographics)** | 338 | 150 | 115 | 305 | 225 |
| **# Outcomes in Final Data Set** | 68 | 105 | 79 | 133 | 99 |
| **% Outcomes in Final Data Set** | 20% | 70% | 69% | 44% | 44% |

The overall prevalence was 0.2. There was significantly higher prevalence for Age: (80, 90] at 0.55. There was significantly lower prevalence for Age: (20,30] at 0 and Age: (30, 40] at 0. There were no significant differences in prevalence found by Sex, Ethnicity/Race, or the intersection of Ethnicity/Race and Sex (Table 5, <u>Supplemental Tables 1-4</u>).

**Table 5:**
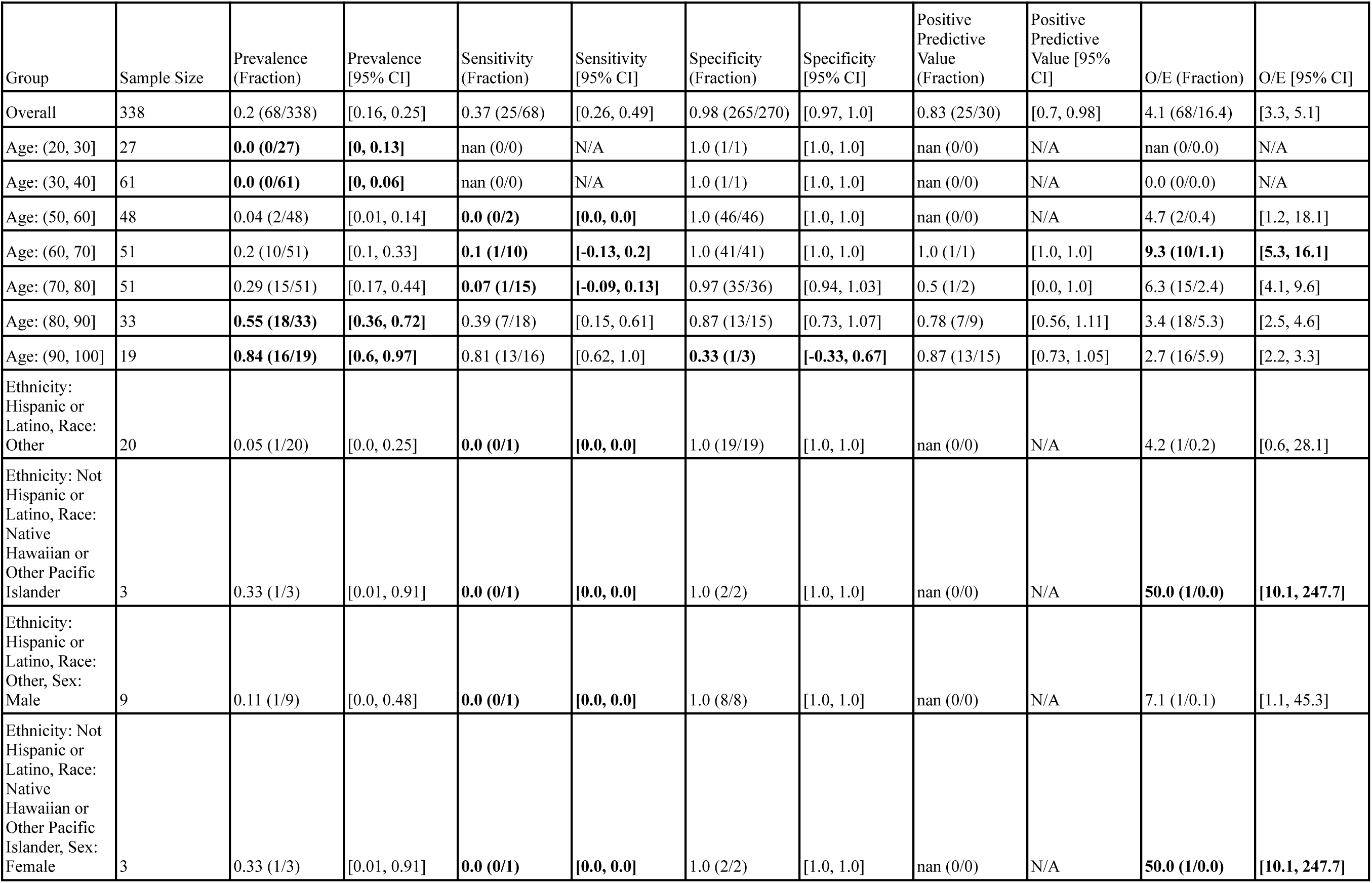
Epic EOL Low Threshold in Primary Care: Reliability and Fairness Audit with significant results. Prevalence, performance and calibration is presented for the overall cohort and for subgroups with significant differences in prevalence, significantly lower performance, or significantly higher O/E (bolded). For the full set of results, see Supplemental Tables 1-4.

The model flagged 30 patients out of 338 (9%), exhibiting low sensitivity (0.37), high specificity (0.98), and high PPV (0.83). The model also underpredicted events relative to clinicians by a factor of O/E = 4.1. There was significantly lower sensitivity for Age: (60, 70] at 0.1 and Age: (70, 80] at 0.07. The model also underpredicted events more for Age: (60, 70], by a factor of O/E = 9.3 (Table 5). For several other groups, there were statistically significant differences in prevalence, performance or O/E, but these subgroups had less than 10 patients to calculate the metric for, making results inconclusive (Table 5).

#### Inpatient Oncology

We calculated we would need a sample size of 19 to achieve an O/E 95% confidence interval of [0.74, 1.34], assuming a 70% prevalence of the positive label. Two clinicians (ARK, KR) completed 225 labels for patients they saw while on service (1-year surprise question answers). Note: each data point corresponds with a unique patient encounter (some patients were included multiple times due to re-hospitalization). Of the 225 labels, 23 did not have a numerical MRN associated and were filtered out, leaving 202 clinician labels fitting the schema.

##### Epic EOL High Threshold in Inpatient Oncology

The final data set size for the Epic EOL - High Threshold model in Inpatient Oncology, was 150 with 105 positive labels after we linked the 202 clinician labels fitting the schema with Epic EOL model predictions and patient demographics (Table 4).

The overall prevalence was 0.7. There was significantly lower prevalence for younger patients (0.23 for Age: (20, 30]). There were no significant differences in prevalence by Sex, Ethnicity/Race, and the intersection of Ethnicity/Race and Sex (Table 6).

**Table 6:**
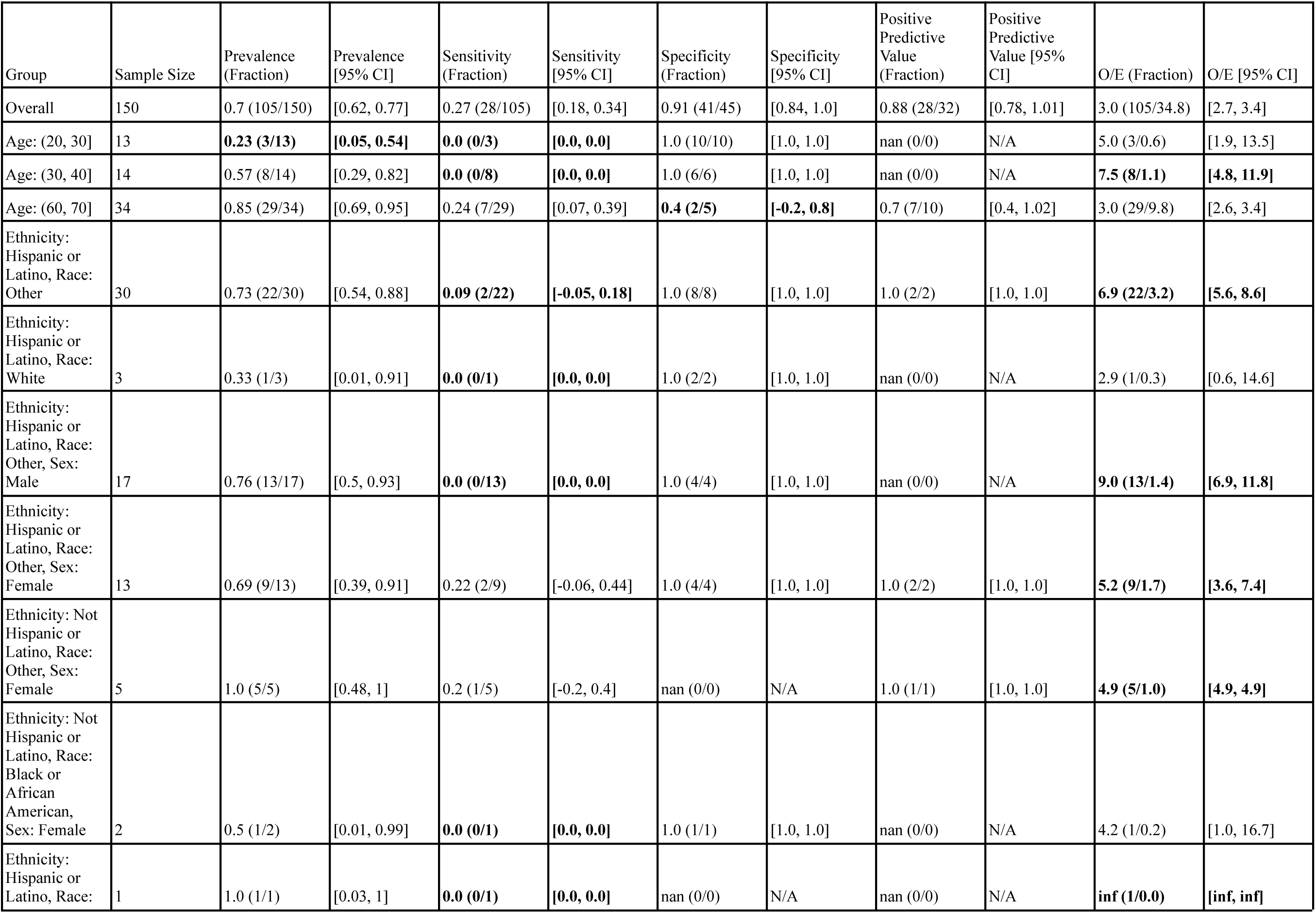
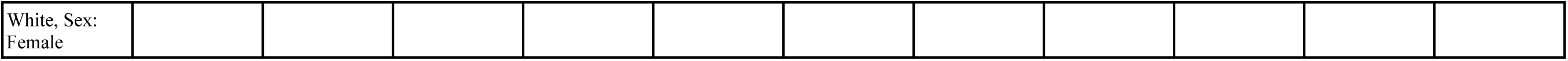
Epic EOL High Threshold in Inpatient Oncology: Reliability and Fairness Audit, significant results. Prevalence, performance and calibration is presented for the overall cohort and for subgroups with significant differences in prevalence, significantly lower performance, or significantly higher O/E (bolded). For the full set of results, see Supplemental Tables 5-8.

The model flagged 32 patients out of 150 (21%) with a sensitivity of 0.27, specificity of 0.91, and PPV of 0.88. The model predicted many fewer events relative to the number of positive clinician labels, with an O/E ratio of 3. Sensitivity for Hispanic or Latino patients with Race “Other” (0.09) was significantly lower than the model’s overall sensitivity (0.27). This was also true for Hispanic or Latino Males with Race “Other” specifically, for which the model’s sensitivity was 0. The model significantly underpredicted events for both subgroups relative to clinicians, with O/E ratios of 6.9 and 9, respectively. Several other subgroups exhibited statistically significant differences in model performance or O/E, but these subgroups had less than 10 patients to calculate the metric for, making such claims inconclusive. See Table 6 for details.

##### Stanford HM ACP in Inpatient Oncology

The final data set size for the Stanford HM ACP model in Inpatient Oncology was 114 with 79 positive labels after we linked the 202 clinician labels fitting the schema with Stanford HM ACP model predictions and patient demographics (Table 4).

The overall prevalence was 0.69. There were no significant differences in prevalence amongst the demographic subgroups considered.

The Stanford HM ACP model flagged 85 patients out of 114 (75%) with sensitivity 0.89, specificity 0.57, and PPV 0.82. The model moderately underestimated events relative to clinicians, with an O/E of 1.7. Model performance and O/E appeared to differ for some subgroups, but these subgroups had less than 10 patients to calculate the metric for, making any associated claims inconclusive. See Table 7 for details.

**Table 7:**
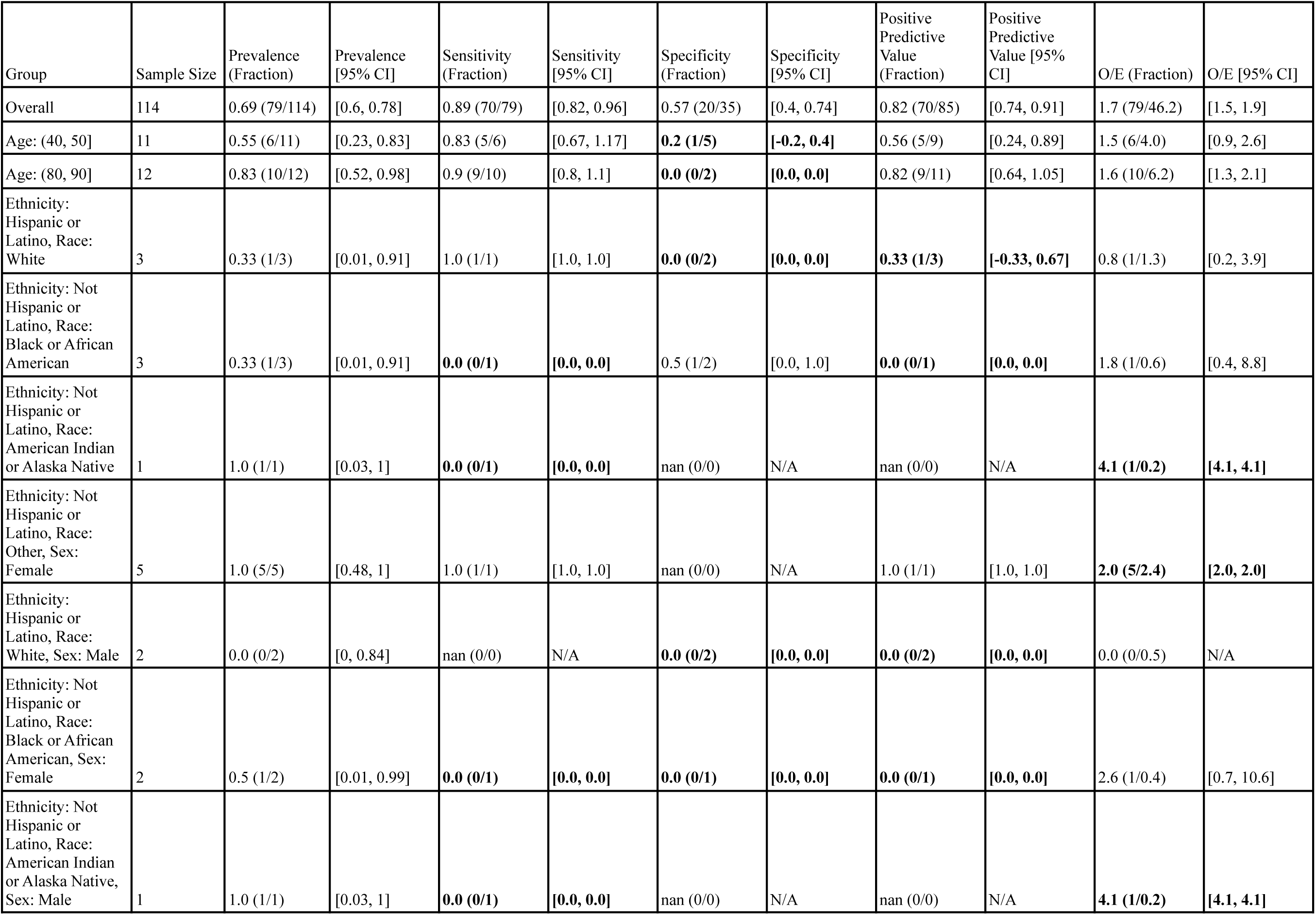
Stanford HM ACP in Inpatient Oncology: Reliability and Fairness Audit with significant results. Prevalence, performance and calibration is presented for the overall cohort and for subgroups with significant differences in prevalence, significantly lower performance, or significantly higher O/E (bolded). For the full set of results, see Supplemental Tables 9-12.

##### Model Comparison in Inpatient Oncology

Comparing model performance in Inpatient Oncology, the Stanford HM ACP model flagged more patients (75% vs 21%), had significantly higher sensitivity (0.89 vs 0.27), and exhibited similar PPV (0.82 vs 0.88, 95% confidence intervals overlap). The Epic EOL High Threshold model had significantly higher specificity (0.91 vs 0.57). Comparing model calibration, the Stanford HM ACP model had significantly better calibration in terms of O/E (1.7 vs 3).

#### Hospital Medicine

We calculated we would need a sample size of 66 to achieve an O/E confidence interval of [0.74, 1.34], assuming a 40% prevalence of the positive label. We solicited 22 clinicians for 545 labels of their patients seen while they were on service (1-year surprise question answers). 18 clinicians responded with 413 labels (76% response rate). Note: each data point corresponds with a unique patient encounter (some patients were included multiple times due to long hospital stays). Four of these were “Maybe” or “TRANSFERRED” and were filtered out, leaving 409 clinician labels fitting the schema.

##### Epic EOL High Threshold in Hospital Medicine

The final data set size for the Epic EOL - High Threshold model in Hospital Medicine, was 305 with 133 positive labels after we linked the 409 clinician labels fitting the schema with Epic EOL model predictions and patient demographics (Table 4).

The overall prevalence was 0.44. Prevalence did not differ by sex, but was significantly higher for older patients (0.76 for Age: (80, 90] and 0.94 for Age: (90, 100]) and significantly lower for younger patients (0.12 for Age: (20, 30] and 0.15 for Age: (30, 40]). Prevalence was also significantly higher for Non-Hispanic Asian patients (0.68) but significantly lower for Hispanic or Latino patients with Race “Other” (0.18) and, in particular, Hispanic or Latino Males of Race “Other” (0.14).

The model flagged 34 out of 305 patients (11%). The model demonstrated a sensitivity of 0.2, specificity of 0.95, and PPV of 0.76. The model underpredicted events relative to clinicians (O/E ratio of 2.5). In particular, the model underestimated events relative to clinicians for Non-Hispanic White Females (O/E = 3.7). Differences in performance and O/E were statistically significant for other subgroups, but these subgroups had less than 10 patients to calculate the metric for, preventing conclusive statements regarding disparate performance. See Table 8 for details.

**Table 8:**
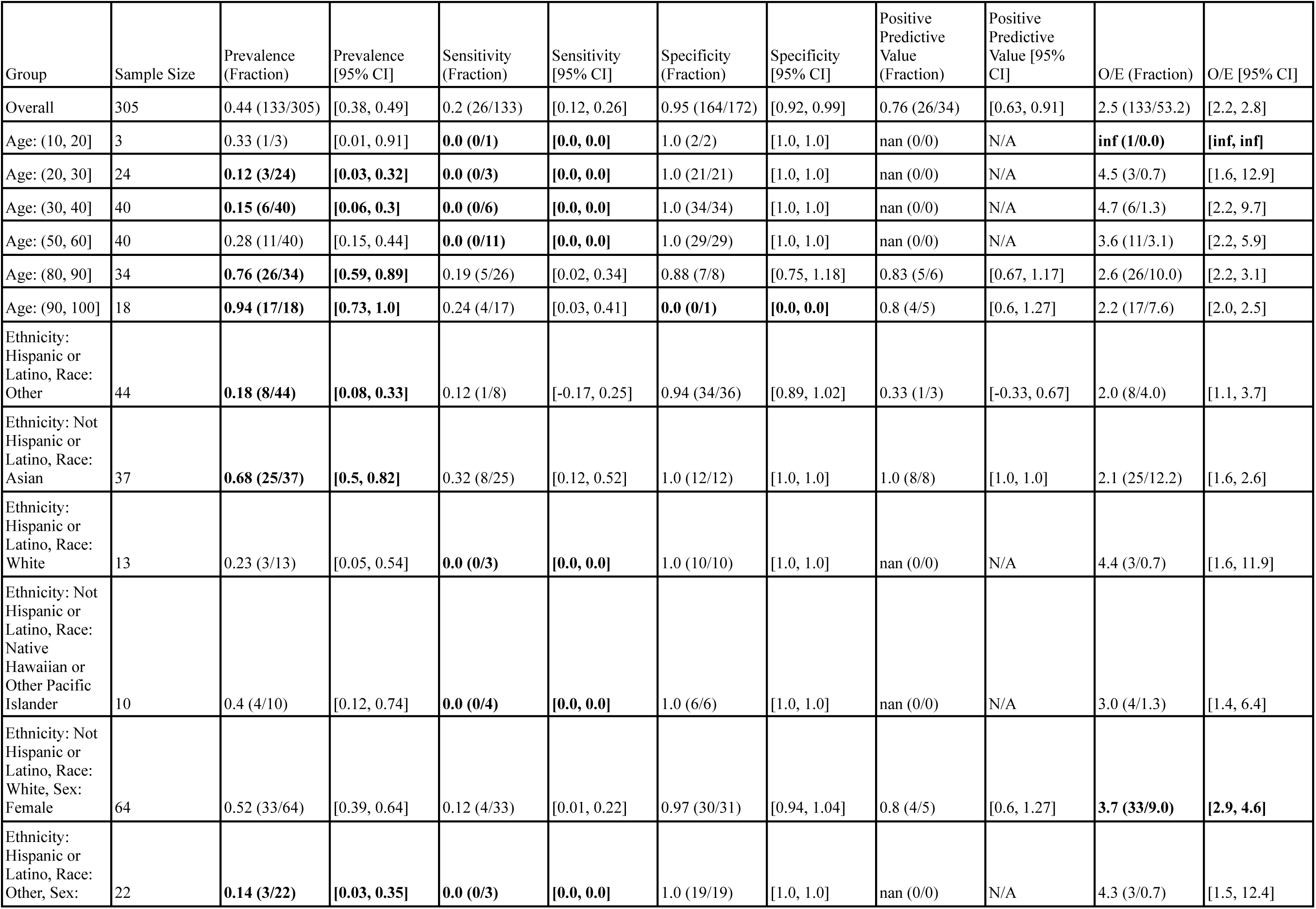
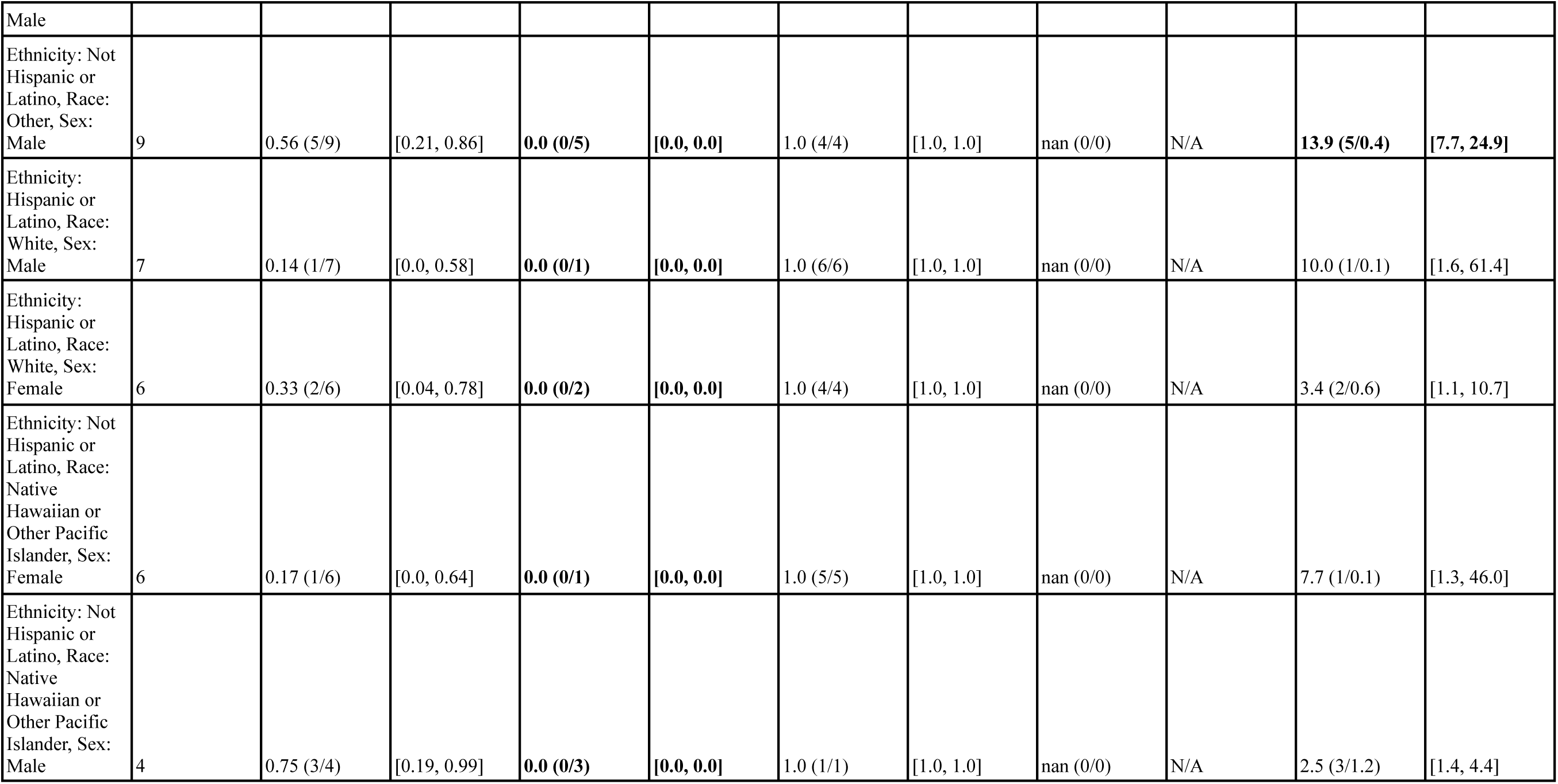
Epic EOL High Threshold in Hospital Medicine: Reliability and Fairness Audit with significant results. Prevalence, performance and calibration is presented for the overall cohort and for subgroups with significant differences in prevalence, significantly lower performance, or significantly higher O/E (bolded). For the full set of results, see Supplemental Tables 13-16.

##### Stanford HM ACP in Hospital Medicine

The final data set size for the Stanford HM ACP model in Hospital Medicine, was 225 with 99 positive labels after we linked the 409 clinician labels fitting the schema with Stanford HM ACP model predictions and patient demographics (Table 4).

The overall prevalence was 0.44. Prevalence was significantly higher for older patients (0.8 for Age: (80, 90], 0.92 for Age: (90, 100]) and significantly lower for younger patients (0.11 for Age: (30, 40]). Prevalence was also significantly lower for Hispanic or Latino patients with Race “Other” (0.16) and significantly higher for Non-Hispanic Asian patients (0.7), especially Non-Hispanic Asian Males (0.81).

The Stanford HM ACP model flagged 85 out of 225 patients (38%), with sensitivity 0.69, specificity 0.87, and PPV 0.8. Relative to clinicians, the model underestimated events by a factor of O/E = 1.5. For patients Age: (90, 100], this underestimation was even more substantial with an O/E ratio of 2.5. Sensitivity was significantly lower (0.17) for patients Age: (50, 60].

Specificity was lower (0.57) for Age: (70, 80]. Relative to the model’s overall PPV, the PPV for Hispanic or Latino patients with Race “Other” was significantly lower (0.29 vs. 0.8). Model performance disparities in other subgroups were inconclusive given they had less than 10 patients to calculate the metric for. See Table 9 for details.

**Table 9:**
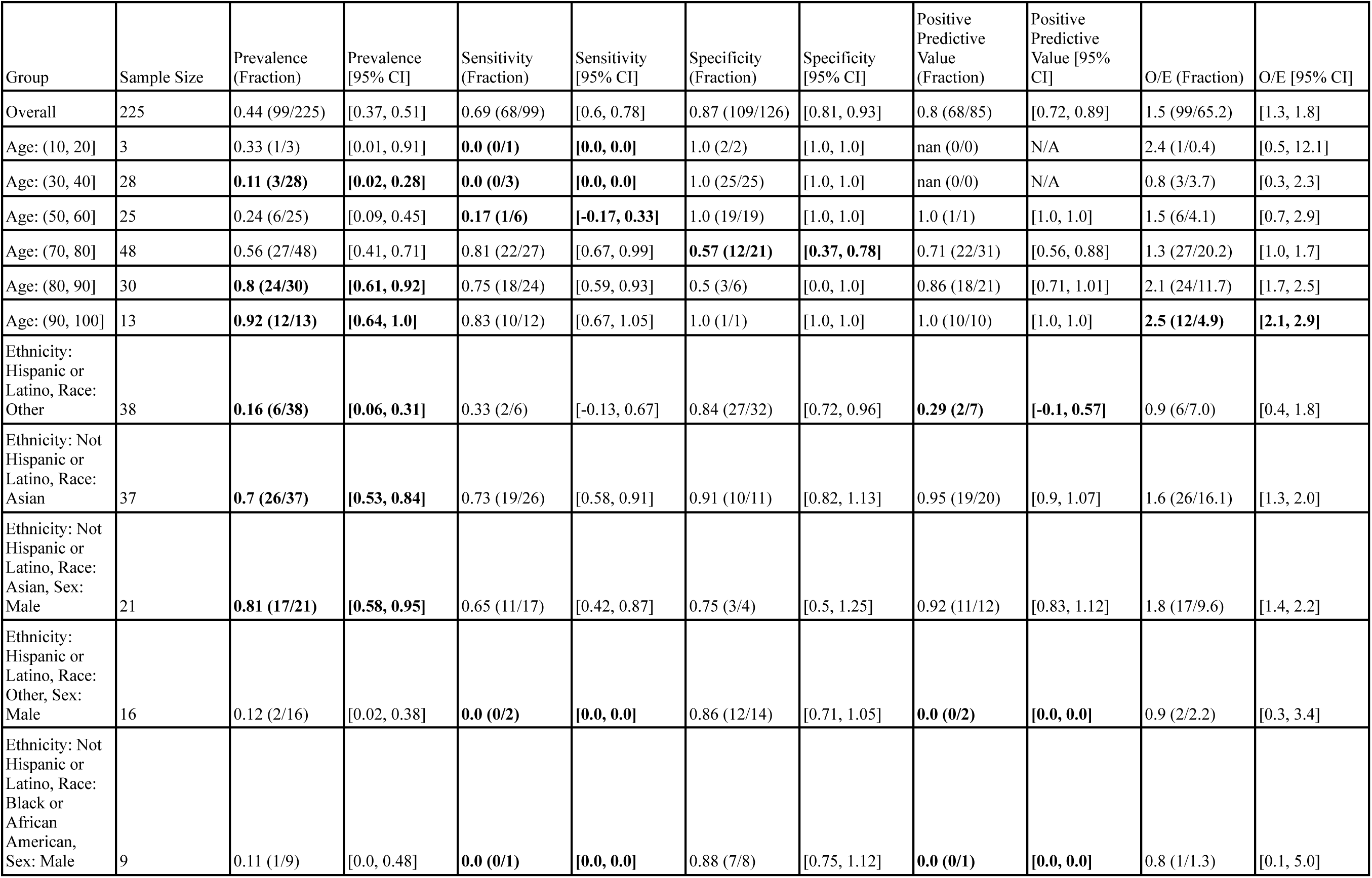
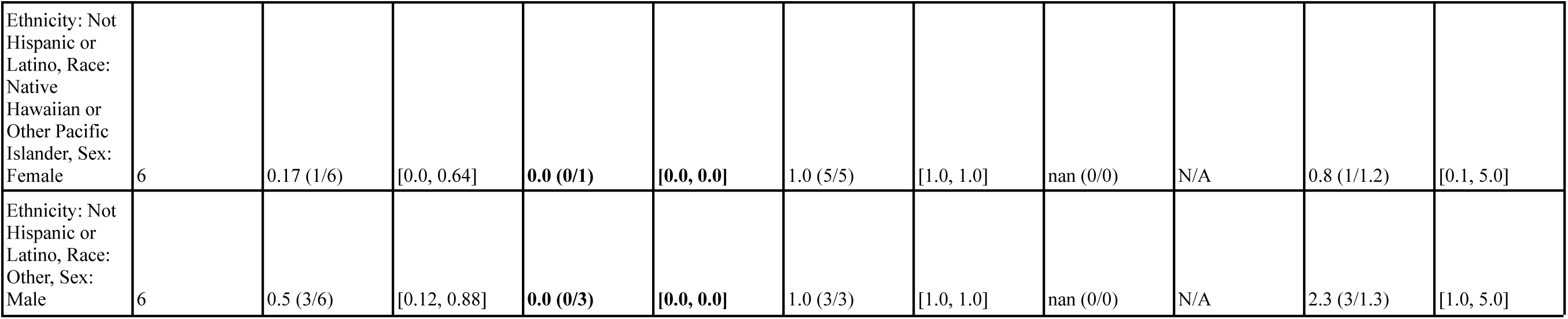
Stanford HM ACP in Hospital Medicine: Reliability and Fairness Audit with significant results. Prevalence, performance and calibration is presented for the overall cohort and for subgroups with significant differences in prevalence, significantly lower performance, or significantly higher O/E (bolded). For the full set of results, see Supplemental Tables 17-20.

##### Model Comparison in Hospital Medicine

Comparing model performance in Hospital Medicine, relative to the Epic EOL – High Threshold model the Stanford HM ACP model flagged more patients (38% vs 11%), had significantly higher sensitivity (0.69 vs 0.2), similar specificity (0.87 vs 0.95, 95% confidence intervals overlap), and similar PPV (0.8 vs 0.76, 95% confidence intervals overlap). Comparing model calibration, the Stanford HM ACP model had significantly better calibration in O/E (1.5 vs 2.5).

### Survey of Decision Makers

After the presentations, we administered a survey about how the audit impacted decision makers’ decision to use the model. We gathered 10 responses: 2 for Primary Care, 5 for Inpatient Oncology and 3 for Hospital Medicine. 7 responses were from Attending Physicians, 1 was from a Physician Assistant, and 2 were from the Lead for the Serious Illness Care Program.

#### Understandings of Reliable/Fair Models

Decision makers used themes of **Accurate** (9/10) and **Consistent** (5/10) when asked to describe what it meant to them for a model to be reliable (Table 10). For example, one response said: “*not brittle (doesn’t give really weird answers if some data are missing)*.”

**Table 10:**
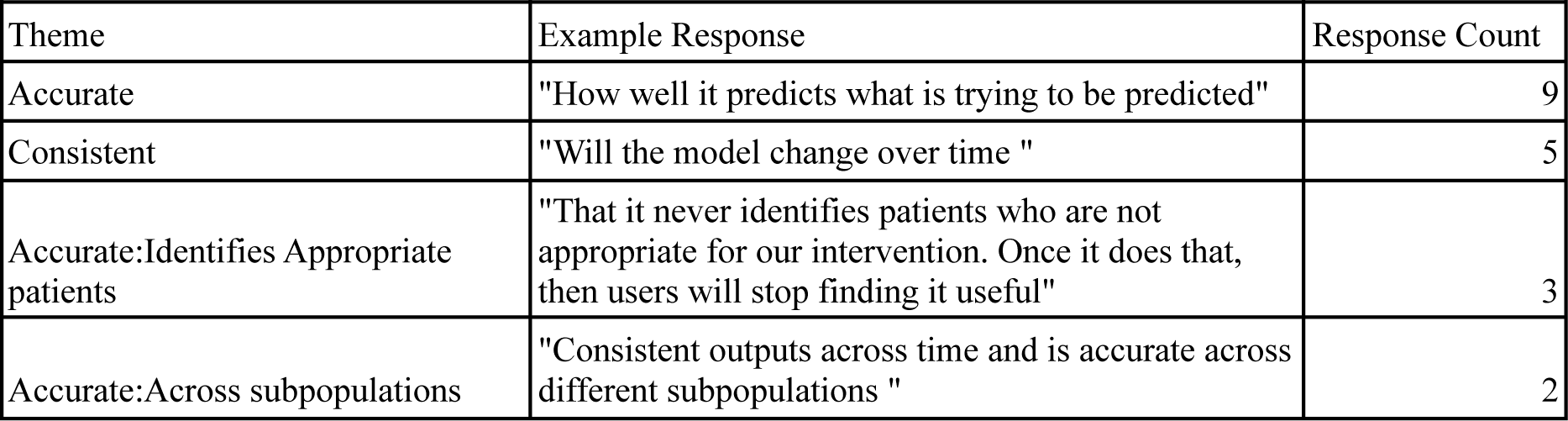
Survey responses to “What does it mean to you for a model to be reliable?”

| Theme | Example Response | Response Count |
| --- | --- | --- |
| Accurate | "How well it predicts what is trying to be predicted" | 9 |
| Consistent | "Will the model change over time " | 5 |
| Accurate:Identifies Appropriate patients | "That it never identifies patients who are not appropriate for our intervention. Once it does that, then users will stop finding it useful" | 3 |
| Accurate:Across subpopulations | "Consistent outputs across time and is accurate across different subpopulations " | 2 |

When asked to describe what it meant to them for a model to be fair, they tended to use themes of **Similar Model Performance across demographics** (6/10) often specifically citing **Race/Ethnicity** (4/10) and **Sex** (4/10) (Table 11). Another common theme was **Depends on How Model is Used** (2/10). For example, one response said: “*… In one context, being more sensitive for patients of a certain group could be good (fair) for those patients, in another context it could be bad (unfair).”*

**Table 11:**
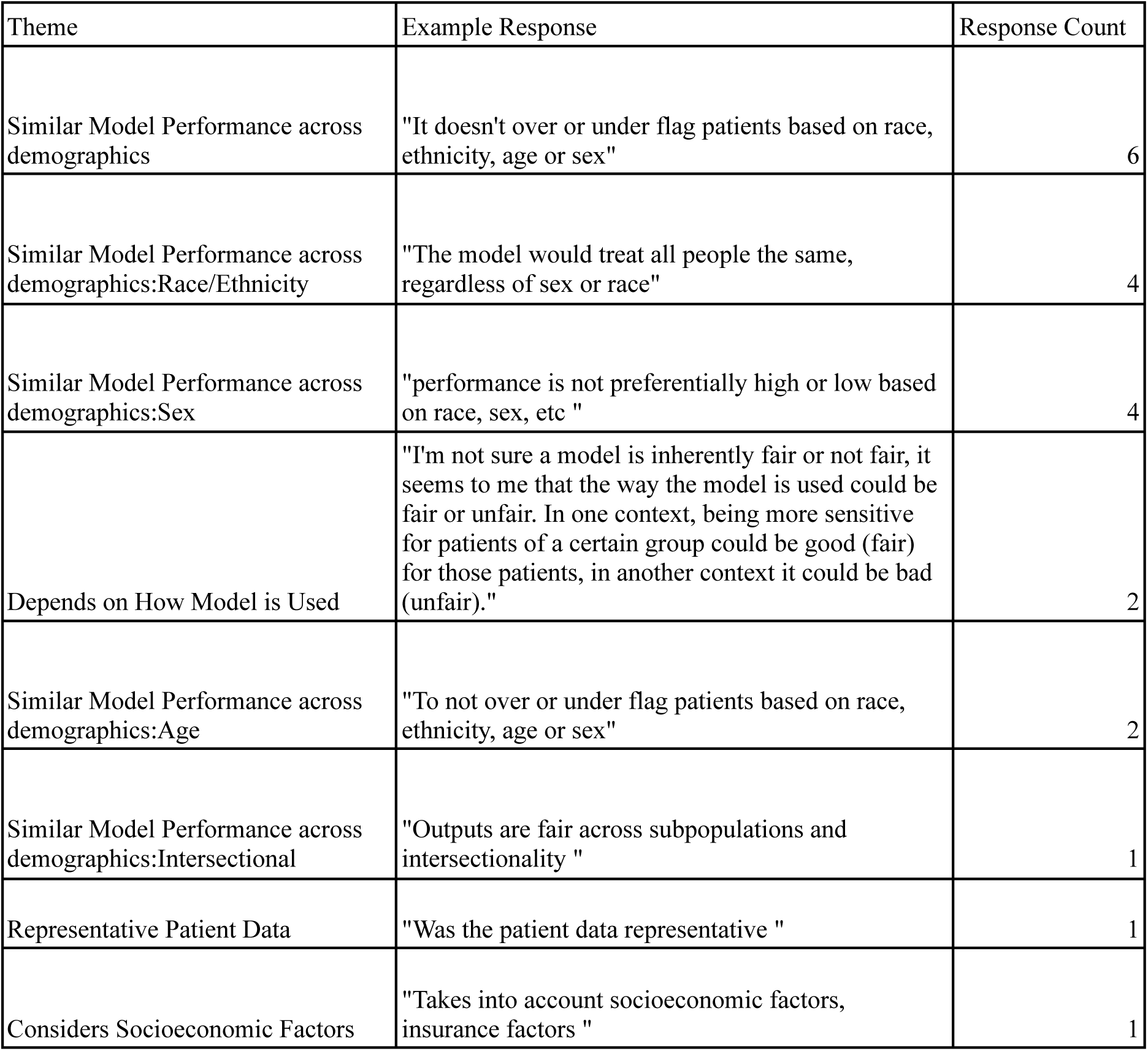
Survey responses to “What does it mean to you for a model to be fair?”

| Theme | Example Response | Response Count |
| --- | --- | --- |
| Similar Model Performance across demographics | "It doesn't over or under flag patients based on race, ethnicity, age or sex" | 6 |
| Similar Model Performance across demographics:Race/Ethnicity | "The model would treat all people the same, regardless of sex or race" | 4 |
| Similar Model Performance across demographics:Sex | "performance is not preferentially high or low based on race, sex, etc " | 4 |
| Depends on How Model is Used | "I'm not sure a model is inherently fair or not fair, it seems to me that the way the model is used could be fair or unfair. In one context, being more sensitive for patients of a certain group could be good (fair) for those patients, in another context it could be bad (unfair)." | 2 |
| Similar Model Performance across demographics:Age | "To not over or under flag patients based on race, ethnicity, age or sex" | 2 |
| Similar Model Performance across demographics:Intersectional | "Outputs are fair across subpopulations and intersectionality " | 1 |
| Representative Patient Data | "Was the patient data representative " | 1 |
| Considers Socioeconomic Factors | "Takes into account socioeconomic factors, insurance factors " | 1 |

Decision makers used a variety of themes to describe their first thoughts on seeing the results of the reliability and fairness audit (<u>Supplemental Table 21</u>). In Primary care, the decision makers used **Excitement** and **Trust to Use the Model For Intended Purpose** (2/2), whereas in Hospital Medicine, they used **Interesting** (3/3). In Inpatient Oncology, 2 of 5 responses referred to **Low Sample Size**, for example “.*..There may be some signals of differences based on age and race/ethnicity groups, but I wonder if this is in part limited by low power.*”

#### Audit Components Affecting Decision making

Decision makers felt that every component of the audit would affect their decision to deploy the model, including Summary Statistics, Performance, and Subgroup Performance (10/10); and Calibration and Subgroup Calibration (both 9/10). When asked for any other information they would want included in the audit to support their decision on whether to deploy a model (<u>Supplemental Table 22</u>), decision makers most commonly responded with **more reliable race data in EHR** (2/10).

#### Drivers and Barriers for Audits and AI model use

Decision makers identified **Findings that AI models are not fair** (10/10), **Findings that AI models are not reliable** (9/10), and **Academic medicine’s push toward racial equity** (9/10) as key drivers to making reliability and fairness audits standard practice (<u>Supplemental Table 23</u>). For key barriers, they tended to identify **Poor demographic data quality** (8/10), **Poor data quality** (6/10), and **Lack of data access** (5/10) (Supplemental Table 24).

Decision makers largely saw **Helps triage patients and identify who would benefit the most** (10/10) and **Shared understanding of patients for our whole care team** (9/10) as key advantages of using AI to support their work (<u>Supplemental Table 25</u>). When asked what cons they see in using an AI model to support their work, decision makers tended to respond with **Lack of transparency of the model** (5/10) and **Takes effort to maintain** (4/10) (<u>Supplemental Table 26</u>).

### Time and Resources required to Perform Audit

We documented the main tasks, persons performing each task, and estimated time required to perform each task in <u>Supplemental File 1</u>, summarizing in Table 12. Note: we estimated response time per clinician using the median time per surprise question from our decision maker survey responses: 1 minute for Primary Care and for Hospital Medicine, and 2 minutes for Inpatient Oncology.

**Table 12:**
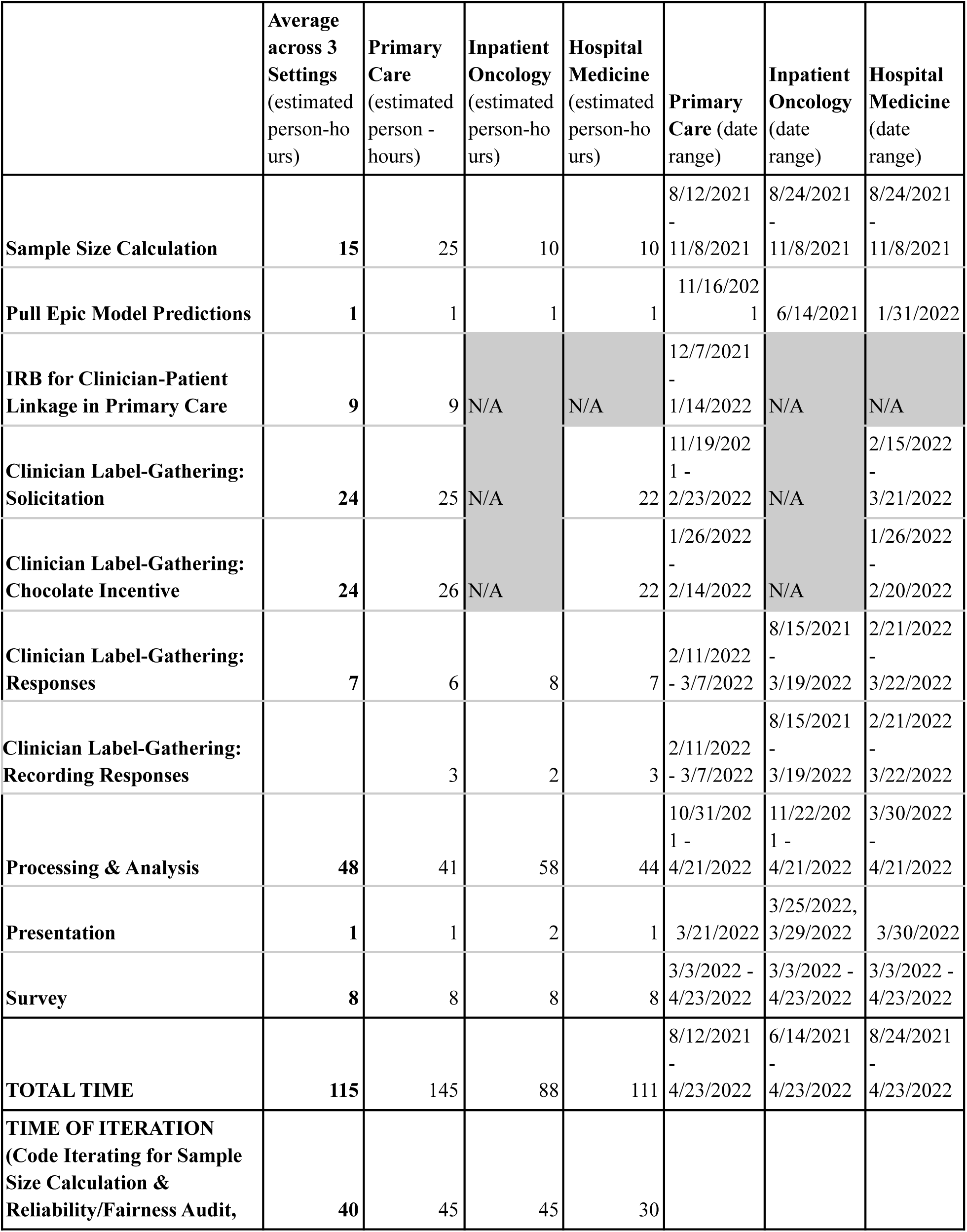
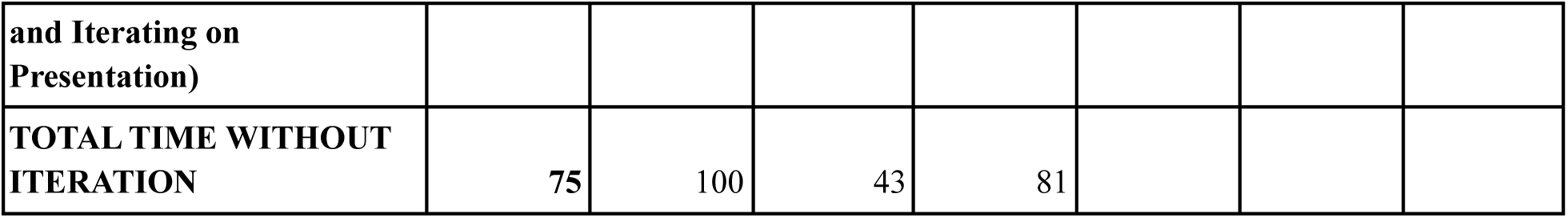
Time and Requirements to generate Reliability and Fairness Audits. For further detail, see <u>Supplemental File 1</u>.

Averaged across the three settings, we spent 115 hours on the audit. Some of the most time-intensive tasks involved processing and analysis of the data (48 person-hours), soliciting clinician labels (24 person-hours), designing and implementing an incentive program to support gathering clinician labels (24 person-hours), and calculating the required sample size (15 person-hours). Notably, the actual responses by the clinicians and recording of responses by the clinicians required less time (9 person-hours), as did designing and implementing the survey (8 person-hours) and presenting to the decision makers (1 person-hours).

Of the 115 hours, we classified 40 (35%) of these hours as iteration time – time that JL spent mainly on iterating on writing code (e.g., for calculating required sample sizes and estimating model performance for each subgroup) or drafting presentation material. If we were to do the same study again at this point, presuming we could bypass the iteration time, the audit could likely be done in 75 hours (65% of total hours).

In calendar time, the audits were completed 8-10 months from the start, underscoring the need for balancing competing priorities amongst both study designers and participants, building relationships among team members to enable the project, and waiting for clinicians to respond.

Lastly, we emphasize key requirements in two categories: *stakeholder relationships* and *data access*. On stakeholder relationships, physicians’ understanding of the best way to communicate with their colleagues and designing appropriate incentives (e.g., chocolate) were crucial to ensure a high response rate. On data access, there were multiple data sources with different access requirements. Some required healthcare system employees to use their privileged access. For example, KS had to extract Epic model predictions from our EHR for us to perform the audit. Similarly, multiple IT subunits had to coordinate to deliver patient panels for us.

Alternatively, other data sources could be accessed using existing data infrastructure. Crucially, our patient demographics and patient visits were already available in a common data format (OMOP-CDM) (44). This allowed iterative querying and refinement to ensure we were pulling the most relevant patients and patient information. Having existing access to a daily hospital census feed and having query access to the hospitalist attending schedules were critical in enabling our hospital medicine clinician labeling workflow (26).

## Discussion

We operationalized reliability and fairness audits of predictive models in ACP, with the best attempt to adhere to model reporting guidelines (22). We highlight key insights and themes across audits below and conclude with recommendations for informaticists and decision makers.

### Key Insights from Model Fairness Audits

We use the Epic EOL High Threshold’s performance for Hispanic patients in Inpatient Oncology as an illustrative example (Figure 2) to show the value of reporting summary statistics, subgroup performance and subgroup calibration. (Note: the specific group is Hispanic/Latino patients with Race listed as Other, but we denote them as “Hispanic” patients here for simplicity).

**Figure 2:**
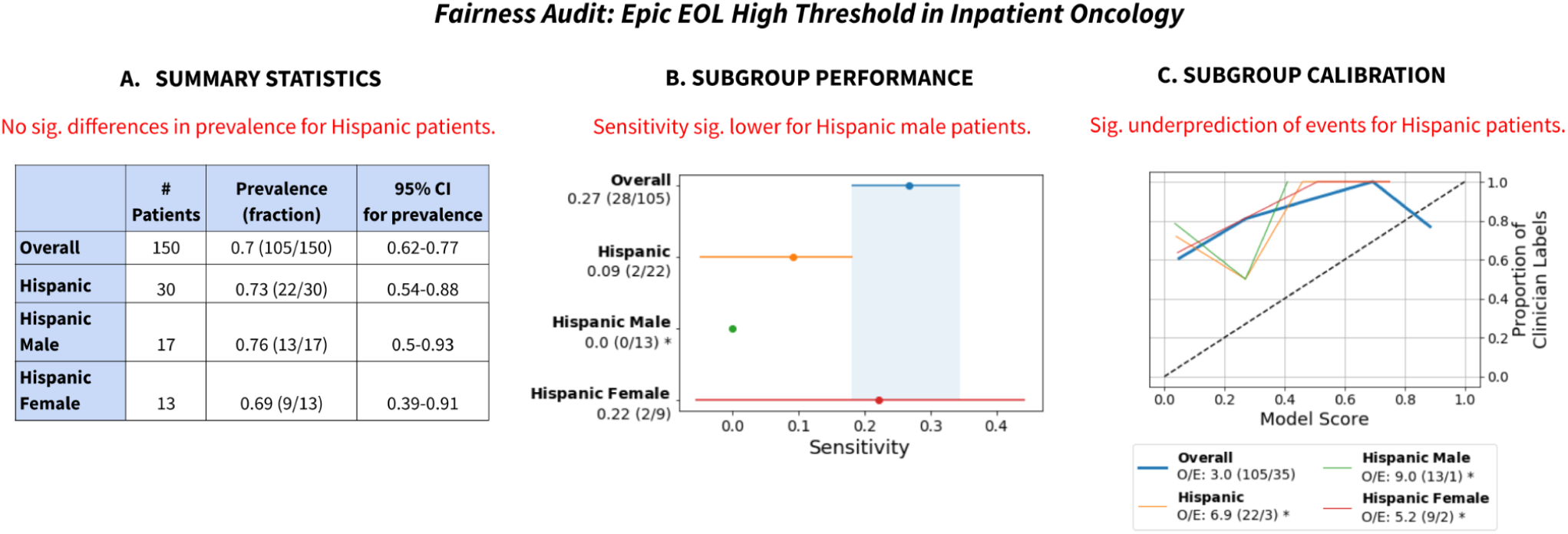
Summary of Fairness Audit Findings, for Hispanic patients for the Epic EOL High Threshold in Inpatient Oncology. A) Summary statistics revealed no significant difference in prevalence for Hispanic patients. B) Subgroup performance revealed decreased sensitivity on Hispanic patients and that it was especially low for Hispanic male patients. C) Subgroup calibration revealed significantly greater miscalibration for Hispanic patients, including for Hispanic female patients and especially for Hispanic male patients. Note: we refer to Hispanic patients with Race listed as Other as Hispanic patients. “Sig.” stands for “statistically significant.”

First, summary statistics revealed no significant differences in prevalence of the outcome label for Hispanic patients, including after disaggregating by the intersection of race and sex (Figure 2A). Assuming no systematic differences in mortality risk or appropriateness of ACP for Hispanic patients vs. Non-Hispanic patients, this reassured us that our surrogate outcome exhibited no obvious signs of bias.

Second, despite insignificant differences in clinician label prevalence, the Epic EOL – High Threshold revealed reduced sensitivity (0.09) for Hispanic patients (Figure 2B). The model only flagged 2 of 22 positive patients identified by clinician review. Disaggregation by the intersection of Race/Ethnicity and Sex revealed that the model had significantly reduced sensitivity (0.0) for Hispanic male patients specifically, flagging 0 of 13 positive patients. This demonstrates the value of analyzing model performance for different subgroups (49) and intersectional subgroups (3).

Third, subgroup calibration revealed significant underprediction of events (O/E: 6.9) for Hispanic patients (Figure 2C), especially Hispanic male patients (O/E: 9.0). The subgroup calibration shows that the model was systematically giving lower scores to Hispanic patients relative to clinicians, which is potentially linked to the model’s lower sensitivity for those groups. Again, this shows how subgroup calibration aids understanding algorithms’ impacts on different groups (4).

Differences in the Epic EOL model’s sensitivity for Hispanics vs. Non-Hispanics and the model’s O/E ratio relative to clinicians for this subgroup also highlights one of the key challenges in using surrogate outcomes (e.g., clinician responses to the surprise question) for reliability and fairness audits. Was the Epic EOL model’s sensitivity low for Hispanic Males because it underestimated true risk, or was it that clinicians overestimated risk for those Hispanic Male patients that the model did not flag? Given the consistency of clinician labels across subgroups, we lean toward the former interpretation, but it is impossible to say with certainty in the absence of an objective ground truth label.

Lastly, in all three cases, reporting numerators and denominators put the metrics in context. There were many otherwise seemingly significant results that were marred by low number of patients to calculate the metric for (e.g. for sensitivity, there may be few patients with the positive label). This is especially true for intersectional subgroups that have low representation in the data set (e.g., American Indian or Alaska Native Males).

### Consistent Themes across Audits

Considering the summary statistics of the data sets, there were generally no differences in prevalence of clinician-generated positive labels by Sex, Race/Ethnicity or Race/Ethnicity and Sex. Out of 5 data sets considered, 4 showed either significantly higher prevalence of positive labels for older patients (Age:(70,80], Age:(80,90], Age:(90,100]) or significantly lower prevalence for younger patients (Age:(20,30], Age:(30,40]). This is consistent with the older patients having worse prognosis than younger patients and thus was not a cause for concern with respect to label bias. However, it was surprising that for the two Hospital Medicine data sets, there was a higher prevalence of positive labels for non-Hispanic Asian patients (including specifically for those of Male sex) and lower prevalence for Hispanic patients for whom Race was listed as Other (including specifically those of Male sex).

Considering the model performance and calibration, in every setting, all models had high PPV at 0.76 or above; several of our clinicians considered this the most important metric, roughly corresponding to “would a clinician agree if the model flagged a patient?”. In Hospital Medicine and Inpatient Oncology, the Epic EOL model at High Threshold tended to flag fewer patients (11%, 21% respectively) than the Stanford HM ACP model (38%, 75%). Meanwhile, the Stanford HM ACP model had higher sensitivity (0.69, 0.89 vs 0.20, 0.27), and better calibration (O/E 1.5, 1.7) than the Epic EOL model (O/E 2.5, 3.0).

Beyond that, the models often had low sensitivities or PPVs or high rate of underprediction (O/E) for several patient subgroups that had less than 10 patients to compute the metric for in the data set. We emphasize that there is a need to increase representation for these groups so that accurate values can be obtained. Such subgroups include Native Hawaiian or Other Pacific Islander patients, American Indian or Alaska Native patients, Hispanic or Latino patients with race “White” or “Other”, and Black or African American patients, among others.

Decision makers overall felt every component of the audit would affect their decision to turn on the model. They most often responded with themes of **Accurate** and **Consistent** for “What does it mean to you for a model to be reliable?”. They most often responded with **Similar Model Performance across demographics**, especially for **Race/Ethnicity** and **Sex** for “What does it mean to you for a model to be fair?”. The most commonly identified key barriers for making reliability and fairness audits standard practice were **Poor demographic data quality, Poor data quality,** and **Lack of data access.**

#### Recommendations for Informaticists

##### 1. Invest in Checking and Improving Data Validity

Our audit was influenced by multiple unreliable data cascades (50) that hindered our ability to draw decisive conclusions regarding model fairness and reliability. Firstly, it is likely that the race/ethnicity variables were inaccurate, given widespread low concordance with patients’ self-identified race/ethnicity found in one of our family medicine clinics (33) and other data sets (34). Thus, a prerequisite for reporting summary statistics and model subgroup performance, as recommended by many model reporting guidelines (9,11–13,15,17,18,20,51), would be better collection of race/ethnicity data. We also again emphasize that race/ethnicity is more a social construct than fixed biological category (32) and the goal of the fairness audit is to understand the demographics of who is represented in data sets and how models impact them. Another data cascade we experienced was large loss of clinician labels after linking these to model predictions and patient demographics (25-27% for the Epic EOL and 44-45% for the Stanford HM ACP, in Inpatient Oncology and Hospital Medicine).

Lastly, it is important to verify the validity of source data in detail i.e., via manual inspection of the raw data, summary statistics, and metadata for all variables used in the audit. For example, the Sex variable we used from the patient demographic table came from a column called “gender_source_value”; OMOP-CDM documentation (45) clarified “*The Gender domain captures all concepts about the sex of a person, denoting the biological and physiological characteristics. In fact, the Domain (and field in the PERSON table) should probably should be called ‘sex’ rather than ‘gender’, as gender refers to behaviors, roles, expectations, and activities in society.*” Relatedly, we found hundreds of visits on a single day for two of the Primary Care providers in the visits table. Our frontline clinicians advised this was likely an artifact given the unrealistic number (AS, WT), so we filtered those two days out.

##### 2. Perform Intersectional Analyses

Intersectional analyses proved crucial as they often lended greater clarity to specific subgroups that were being impacted. For example, in Inpatient Oncology, the Epic EOL-High Threshold had low sensitivity (2/22) for Hispanic patients and when disaggregated, specifically had a sensitivity of 0% (0/13) for Hispanic male patients. This would not have been recognized if only looking at sex or race/ethnicity individually. This phenomenon has been discussed in Kimberlé Crenshaw’s pioneering intersectionality research to specifically address discrimination against Black women, who often face distinct barriers and challenges relative to White women or Black men (15, 52).

Intersectional subgroup analyses are not difficult to perform, as generating intersectional demographics from one-hot encoded columns only requires performing a logical intersection operation between demographic one-hot encoded columns. However, care must be taken in interpretation of these subgroup analyses as many intersectional subgroups will have poor representation even in large overall sample sizes. Below, we discuss strategies to aid in interpreting results from less frequently represented subgroups.

##### 3. Contextualize Small Sample Sizes By Calculating Confidence Intervals and Reporting Metrics as Fractions

Small sample sizes of certain subgroups should not be a reason to not consider the subgroups. Proper interpretation of subgroup audit results can be supported by 1) using confidence intervals (e.g. via the bootstrap or exact analytical approaches) to appropriately capture sampling variation and 2) reporting metrics with the involved whole numbers (e.g. numerator and denominator, or number of patients) so that if values are extreme, they can be considered in context. For example, several of our bootstrap confidence intervals did not have any width due to there only being one data point from which to resample. (In future work, we would use analytical methods to calculate exact confidence intervals for small sample sizes, such as the Clopper-Pearson interval (47)).

It is especially important to not ignore small sample sizes as doing so can contribute to understudying patient subgroups, especially those that are underrepresented in healthcare data sets due to societal inequities and structural racism. For example, Indigenous peoples have regularly been excluded from COVID-19 data (53) and American Indian and Alaska Native Peoples have often been ignored in data sets due to aggregate analyses (54). Devising sampling strategies in advance to account for known underrepresented populations can help mitigate these issues (e.g., by oversampling underrepresented minorities or increasing sample sizes so that tests for model performance discrepancies between subgroups are adequately powered).

##### 4. Provider-Patient Linkage are Necessary Data to Perform Audits Using Expert-Generated Labels

Before performing the audit, we did not realize how important it was to be able to generate a list of relevant patients for whom the clinicians would feel comfortable answering the surprise question. Concretely, our clinician annotators felt most comfortable providing labels (the “surprise question”) for patients that they had cared for recently. For Primary Care, this required finding recent visits (available in our OMOP-CDM infrastructure) and linking that with patient panels (which we retrieved from business analysts). For Hospital Medicine, this required linking a daily hospital census feed that had assigned treatment teams, with attending-treatment teams. Informatics teams should view clinician-patient linkage as necessary to perform audits in cases where clinician-generated labels are required.

#### Recommendations for Decision Makers

##### 1. Acknowledge Limits on Data Quality for Evaluation

Decision makers should recognize the limitations of data quality when performing audits. Race/ethnicity data is likely inaccurate unless proven otherwise given the widespread low concordance with patients’ self-identification, as found in our and other data sets (33, 34). Surrogate clinician-generated outcomes used may also be imperfect: our clinician surprise question (a surrogate outcome for appropriateness of an ACP consultation) did not include blinding to the Stanford HM ACP model because it was actively in use as an Epic column as part of the Hospital Medicine SICP implementation. Moreover, while our clinician surprise question generally did not exhibit any obvious differences across ethnicity/race, other studies have found that using surrogate outcomes (e.g., health spending as a proxy for health risk) can exacerbate existing disparities in health (e.g., by estimating that Black patients are at lower health risk because health spending for Black patients has historically been lower than for White patients) (4). Lastly, there were many dropped patients due to lack of an associated model prediction which, if not missing at random, could affect the reliability of our audit.

##### 2. Require Reliability and Fairness Audits of Models Before Deployment

Our work demonstrates that it is feasible to do thorough reliability and fairness audits of models according to model reporting guidelines, despite low adherence to such guidelines for many deployed models (22). In particular, beyond the usual aggregate model performance metrics, it is straightforward to perform pre-study sample size calculations (41), to report confidence intervals on performance metrics (e.g. using bootstrap sampling), to report summary statistics of the evaluation dataset by subgroup, to share calibration plots and calibration measures, and to do subgroup and intersectional subgroup analyses (3, 15). 90% of our decision makers felt that summary statistics, model performance, model calibration, model subgroup performance and model subgroup calibration affected their decision on whether to turn on the model.

Such audits can be performed by internal organizational teams responsible for deploying predictive models in healthcare (23, 55), with the caveat that internal audits may have limited independence and objectivity (23). Alternatively, regulators may conduct such audits, such as the Food and Drug Administration (FDA)’s proposed Digital Health Software Precertification Program which evaluates real world performance of software as a medical device (56). A more likely scenario is the emergence of community standards (57) that provide consensus guidance on responsible use of AI in Healthcare. We propose that the cost of performing such audits be included in the operating cost of running a care program in a manner similar to how IT costs are currently paid for, with a specific carveout to ensure audits are performed and needed resources are fundedT.

##### 3. Enable Audits via connecting Impacted Stakeholders and Informaticists

Our decision makers facilitated relationships with their colleagues in Primary Care, Inpatient Oncology and Hospital Medicine that enabled generation of sufficient clinician labels for us to perform our external validation with excellent response rates. This shows the value of interdisciplinary teams and how important it is to honor the trust that comes with personal connections (27,58,59). Without this strong relationship, we would have been unable to perform our analysis.

##### 4. Interpret Fairness Audits in Context of the Broader Sociotechnical System

Fairness is not solely a property of a model but rather encopmpases the broader sociotechnical system in which people are using a model (60). As one of the decision makers noted, “*I’m not sure a model is inherently fair or not fair,. In one context, being more sensitive for patients of a certain group could be good (fair) for those patients, in another context it could be bad (unfair).*” Furthermore, fairness is not just a mathematical property, but it involves process, is contextual, and can be contested (60). Thus, we note that a fairness audit depicting a model in a favorable light does not by itself *prevent* unfair treatment of patients nor guarantee that use of the model will reduce health disparities.

## Conclusion

Despite frequent recommendations by model reporting guidelines, reliability and fairness audits are not often performed for AI models used in health care (21, 22). With respect to reliability, there is a gap in reporting external validation with performance metrics, confidence intervals, and calibration plots. With respect to fairness, there is a gap in reporting summary statistics, subgroup performance and subgroup calibration.

In this work, we audited two AI models, the Epic EOL Index and a Stanford HM ACP model, which were considered for use to support ACP in three care settings: *Primary Care, Inpatient Oncology and Hospital Medicine*. We calculated minimum necessary sample sizes, gathered ground truth labels from clinicians, and merged those labels with model predictions and patient demographics to create the audit data set. In terms of reliability, all models exhibited a PPV of 0.76 or above in all settings, which clinicians identified as the most important metric. In Inpatient Oncology and Hospital Medicine, the Stanford HM ACP model had higher sensitivity and calibration. Meanwhile, the Epic EOL model flagged fewer patients than the Stanford HM ACP model. In terms of fairness, the clinician-generated data set exhibited few differences in prevalence by sex or ethnicity/race. In Primary Care, Inpatient Oncology, and Hospital medicine the Epic EOL model tended to have lower sensitivity in Hispanic/Latino Male patients with Race listed as “Other”. The Stanford HM ACP model similarly had low sensitivity for this subgroup in Hospital Medicine but not in Inpatient Oncology.

The audit required 115 person-hours, but every component of the audit was valuable, affecting decision makers’ consideration on whether to turn on the models. Key requirements for the audit were 1) stakeholder relationships, which enabled gathering ground truth labels and presenting to decision makers, and 2) data access, especially establishing linkages between providers and patients under their care. For future audits, we recommend recognizing data issues upfront (especially race/ethnicity data), handling small sample sizes by showing confidence intervals and reporting metrics as fractions, and performing intersectional subgroup analyses. Above all, we recommend that decision makers require reliability and fairness audits before using AI models to guide care. With established processes, the 8-10 month calendar time can be compressed to a few weeks given that actual person hours were approximately 3 weeks of effort.

## Data Availability

The data sets containing summary statistics and model performance and calibration metrics for this study can be found in the following attached Zip folder:

https://drive.google.com/file/d/1-VB-kuvK2dFy5evCZbS2qe6DkWXmISv_/view?usp=sharing

## Author Contributions

JL, AS, SW, ARK, BE, RCL, LS, KR, MFG, SL, WT and NHS contributed to study conception and design. MH, BS and WT engaged key clinical stakeholders. JL performed sample size calculation, advised by YX and SP. KS pulled Epic model predictions. NP and PD supported data access.

AS, SW and ARK designed clinician labeling processes for Primary Care, Hospital Medicine and Inpatient Oncology, respectively. AS, SW, ARK, RCL and LS engaged clinicians to perform labeling. AS, SW, ARK, RCL, LS, KR, and others performed labeling.

RF implemented the incentive process for clinician labeling, advised by AS, BS and WT. RF and JL solicited and recorded labels for Primary Care and Hospital Medicine, respectively.

JL and SL performed data set linking. JL performed quantitative data analysis, advised by AC, SF, BE, SP, YX and NHS.

JL designed survey, advised by SW, AC, MS, AG and WT. JL presented results and surveyed AS, SW, ARK, RCL, LS, KR, MFG, SC and WT. JL performed qualitative data analysis, advised by MS and WT.

JL wrote first draft of manuscript. JL, AC, SF, MF, and BE wrote and edited sections of manuscript. All authors contributed to manuscript revision, read, and approved the submitted version.

## Data Availability

The data sets containing summary statistics and model performance and calibration metrics for this study can be found in the following attached Zip folder: https://drive.google.com/file/d/1-VB-kuvK2dFy5evCZbS2qe6DkWXmISv_/view?usp=sharing

https://drive.google.com/file/d/1-VB-kuvK2dFy5evCZbS2qe6DkWXmISv_/view?usp=sharing

## Acknowledgments

We would like to thank the primary care faculty at the Stanford Division of Primary Care and Population Health and hospital medicine attending physicians at the Stanford Department of Medicine for their support, time and expertise in generating the labels. We would like to thank Victor Cheng, Henry Nguyen and Ryan Bencharit for support for delivering the Primary Care patient panel.

We would like to thank Julian Genkins, Naveed Rabbani, Richard Yoo, and Lance Downing for advising on this work. We would like to thank Randy Nhan, Samantha Lane and Nicholas Kenji Taylor for their poster presentation with Amelia Sattler, and for advising regarding the validity of race/ethnicity in the EHR. We would like to thank Anand Avati and Sehj Kashyap for designing the initial hospital medicine ACP email system, and Anand specifically for supporting with initial suggestions about the sample size analysis.

