## Supplement for "Considerations in the Reliability and Fairness Audits of Predictive Models for Advance Care Planning"

[1] Center for Biomedical Informatics Research, Department of Medicine, Stanford University School of Medicine
[2] Division of Primary Care and Population Health, Department of Medicine, Stanford University School of Medicine
[3] Division of Hospital Medicine, Department of Medicine, Stanford University School of Medicine
[4] Division of Oncology, Department of Medicine, Stanford University School of Medicine
[5] Serious Illness Care Program, Department of Medicine, Stanford University School of Medicine
[6] Department of Radiation Oncology, Stanford University School of Medicine
[7] Stanford Health Care
[8] Technology & Digital Solutions, Stanford Health Care and Stanford University School of Medicine
[9] Stanford Healthcare AI Applied Research Team, Stanford University School of Medicine
[10] Clinical Excellence Research Center, Stanford University School of Medicine

**Supplemental Methods** 3
Survey 3
Supplemental Methods: Survey Instrument. 9
Definitions of Metrics 9

**Supplemental Files:** 9
Supplemental File 1: Time and Requirements to Generate Reliability/Fairness Audit. 9

**Supplemental Figures:** 9
Supplemental Figure 1: Message to Primary Care clinicians soliciting surprise questions, staff message. 10
Supplemental Figure 2: Message to Hospital Medicine physicians soliciting surprise questions, email. 11

**Supplemental Tables:** 12
Epic EOL Low Threshold in Primary Care 12
Supplemental Table 1: Epic EOL Low Threshold in Primary Care: Reliability and Fairness Audit by Sex. Significant differences in prevalence, significantly lower performance, or significantly higher O/E are bolded. 12
Supplemental Table 2: Epic EOL Low Threshold in Primary Care: Reliability and Fairness Audit by Age. Significant differences in prevalence, significantly lower performance, or significantly higher O/E are bolded. 13

Supplemental Table 3: Epic EOL Low Threshold in Primary Care: Reliability and Fairness Audit by Ethnicity/Race. Significant differences in prevalence, significantly lower performance, or significantly higher O/E are bolded. 14
Supplemental Table 4: Epic EOL Low Threshold in Primary Care: Reliability and Fairness Audit by Ethnicity/Race and Sex. Significant differences in prevalence, significantly lower performance, or significantly higher O/E are bolded. 16
Epic EOL High Threshold in Inpatient Oncology 16
Supplemental Table 5: Epic EOL High Threshold in Inpatient Oncology: Reliability and Fairness Audit by Sex. Prevalence, performance and calibration is presented for the overall cohort and for subgroups with significant differences in prevalence, significantly lower performance, or significantly higher O/E (bolded). 16
Supplemental Table 6: Epic EOL High Threshold in Inpatient Oncology: Reliability and Fairness Audit by Age. Significant differences in prevalence, significantly lower performance, or significantly higher O/E are bolded. 17
Supplemental Table 7: Epic EOL High Threshold in Inpatient Oncology: Reliability and Fairness Audit by Ethnicity/Race. Significant differences in prevalence, significantly lower performance, or significantly higher O/E are bolded. 18
Supplemental Table 8: Epic EOL High Threshold in Inpatient Oncology: Reliability and Fairness Audit by Ethnicity/Race and Sex. Significant differences in prevalence, significantly lower performance, or significantly higher O/E are bolded. 19
Stanford HM ACP in Inpatient Oncology 19
Supplemental Table 9: Stanford HM ACP in Inpatient Oncology: Reliability and Fairness Audit by Sex. Significant differences in prevalence, significantly lower performance, or significantly higher O/E are bolded. 19
Supplemental Table 10: Stanford HM ACP in Inpatient Oncology: Reliability and Fairness Audit by Age. Significant differences in prevalence, significantly lower performance, or significantly higher O/E are bolded. 20
Supplemental Table 11: Stanford HM ACP in Inpatient Oncology: Reliability and Fairness Audit by Ethnicity/Race. Significant differences in prevalence, significantly lower performance, or significantly higher O/E are bolded. 21
Supplemental Table 12: Stanford HM ACP in Inpatient Oncology: Reliability and Fairness Audit by Ethnicity/Race and Sex. Significant differences in prevalence, significantly lower performance, or significantly higher O/E are bolded. 22
Epic EOL High Threshold in Hospital Medicine 22
Supplemental Table 13: Epic EOL High Threshold in Hospital Medicine: Reliability and Fairness Audit by Sex. Significant differences in prevalence, significantly lower performance, or significantly higher O/E are bolded. 23
Supplemental Table 14: Epic EOL High Threshold in Hospital Medicine: Reliability and Fairness Audit by Age. Significant differences in prevalence, significantly lower performance, or significantly higher O/E are bolded. 24
Supplemental Table 15: Epic EOL High Threshold in Hospital Medicine: Reliability and Fairness Audit by Ethnicity/Race. Significant differences in prevalence, significantly lower performance, or significantly higher O/E are bolded. 25

Supplemental Table 16: Epic EOL High Threshold in Hospital Medicine: Reliability and Fairness Audit by Ethnicity/Race and Sex. Significant differences in prevalence, significantly lower performance, or significantly higher O/E are bolded. 26
Stanford HM ACP in Hospital Medicine 26
Supplemental Table 17: Stanford HM ACP in Hospital Medicine: Reliability and Fairness Audit by Sex. Significant differences in prevalence, significantly lower performance, or significantly higher O/E are bolded. 26
Supplemental Table 18: Stanford HM ACP in Hospital Medicine: Reliability and Fairness Audit by Age. Significant differences in prevalence, significantly lower performance, or significantly higher O/E are bolded. 27
Supplemental Table 19: Stanford HM ACP in Hospital Medicine: Reliability and Fairness Audit by Ethnicity/Race. Significant differences in prevalence, significantly lower performance, or significantly higher O/E are bolded. 28
Supplemental Table 20: Stanford HM ACP in Hospital Medicine: Reliability and Fairness Audit by Ethnicity/Race and Sex. Significant differences in prevalence, significantly lower performance, or significantly higher O/E are bolded. 29
Clinical Decision Maker Survey Responses 30
Supplemental Table 21: Survey responses to "What are the first thoughts that came to your mind on seeing the results of the reliability and fairness audit?" 32
Supplemental Table 22: Survey responses to "Is there any other information you would want included in this audit to support your decision on whether to deploy a model? If so, what?" 33
Supplemental Table 23: Survey responses to "What are some key drivers to making these reliability and fairness audits standard practice?" 34
Supplemental Table 24: Survey responses to "What are some key barriers to making these reliability and fairness audits standard practice?" 35
Supplemental Table 25: Survey responses to "As a clinical decisionmaker, what pros do you see in using an AI model to support your work?" 35
Supplemental Table 26: Survey responses to "As a clinical decisionmaker, what cons do you see in using an AI model to support your work?" 36

### Supplemental Methods

#### *Survey Instrument*

### Reliability and Fairness Audit - Clinical Decision maker Survey

**** (not shared) Switch account

* Required

As a clinical decisionmaker, what pros do you see in using an AI model to support your work? *

Please check all that apply.

- [ ] Helps triage patients and identify who would benefit the most
- [ ] Reduces work for me
- [ ] Shared understanding of patients for our whole care team
- [ ] I do not see any pros to using an AI model to support my work.
- [ ] Other:

As a clinical decisionmaker, what cons do you see in using an AI model to support your work? *

Please check all that apply.

- [ ] I disagree with the model
- [ ] Loss of my decisionmaking autonomy
- [ ] Pressure to act even if I disagree with the model
- [ ] Lack of transparency of the model
- [ ] Takes effort to maintain
- [ ] I do not see any cons to using an AI model to support my work.
- [ ] Other:

What does it mean to you for a model to be reliable? *

Your answer

What does it mean to you for a model to be fair? *

Your answer

What are the first thoughts that came to your mind on seeing the results of the reliability and fairness audit? *

Your answer

For each component of the audit, would it affect your decision to deploy a model? *

| | Yes, this would affect my decision to deploy the model. | No, this would not affect my decision to deploy the model. | Prefer not to answer |
|---|---|---|---|
| Summary Statistics | ◯ | ◯ | ◯ |
| Performance | ◯ | ◯ | ◯ |
| Calibration | ◯ | ◯ | ◯ |
| Subgroup Performance | ◯ | ◯ | ◯ |
| Subgroup Calibration | ◯ | ◯ | ◯ |

Is there any other information you would want included in this audit to support your decision on whether to deploy a model? If so, what?

Your answer

How many minutes on average did it take you to answer the surprise question for one patient? *

Your answer

What are some key drivers to making these reliability and fairness audits standard practice? *

- [ ] Findings that AI models are not reliable
- [ ] Findings that AI models are not fair
- [ ] Academic medicine's push toward racial equity
- [ ] I do not see any key drivers to making reliability and fairness audits standard practice.
- [ ] Other:

What are some key barriers to making these reliability and fairness audits standard practice? *

- [ ] Lack of data access
- [ ] Poor data quality
- [ ] Poor demographic data quality
- [ ] Lack of knowledge about how to do an audit
- [ ] Lack of data science expertise in my practice setting
- [ ] The reliability of deployed AI models is not prioritized
- [ ] The fairness of deployed AI models is not prioritized
- [ ] Audits are not built into our incentives
- [ ] I do not see any barriers to making reliability and fairness audits standard practice.
- [ ] Other:

Please describe your role as a clinical decisionmaker in your setting. *

Your answer

Any other questions or comments related to this audit?

Your answer

Name *

Your answer

Setting *

- Inpatient Oncology
- Primary Care
- Hospital Medicine
- Other:

Submit Clear form

Never submit passwords through Google Forms.

This form was created inside of Stanford University. Report Abuse

Google Forms

### Supplemental Files:

**Supplemental File 1: Time and Requirements to Generate Reliability/Fairness Audit.**
This file is available as a Google Sheets file at https://docs.google.com/spreadsheets/d/12qKv8iguvnyO5WWT5vGt4M_Kwv6yRVpCs3Mv-3RVUl4/edit?usp=sharing

### Supplemental Figures:

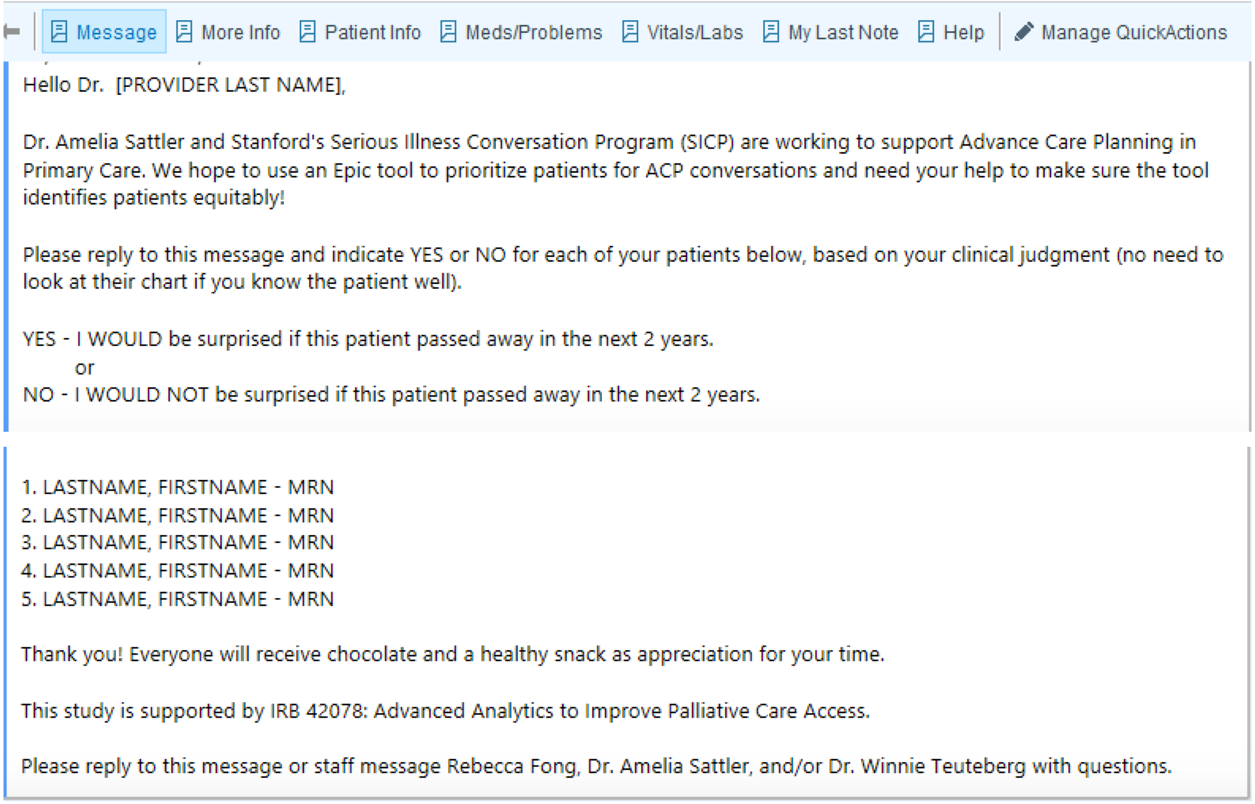
Message | More Info | Patient Info | Meds/Problems | Vitals/Labs | My Last Note | Help | Manage QuickActions

Hello Dr. [PROVIDER LAST NAME],

Dr. Amelia Sattler and Stanford's Serious Illness Conversation Program (SICP) are working to support Advance Care Planning in Primary Care. We hope to use an Epic tool to prioritize patients for ACP conversations and need your help to make sure the tool identifies patients equitably!

Please reply to this message and indicate YES or NO for each of your patients below, based on your clinical judgment (no need to look at their chart if you know the patient well).

YES - I WOULD be surprised if this patient passed away in the next 2 years.
or
NO - I WOULD NOT be surprised if this patient passed away in the next 2 years.

1. LASTNAME, FIRSTNAME - MRN
2. LASTNAME, FIRSTNAME - MRN
3. LASTNAME, FIRSTNAME - MRN
4. LASTNAME, FIRSTNAME - MRN
5. LASTNAME, FIRSTNAME - MRN

Thank you! Everyone will receive chocolate and a healthy snack as appreciation for your time.

This study is supported by IRB 42078: Advanced Analytics to Improve Palliative Care Access.

Please reply to this message or staff message Rebecca Fong, Dr. Amelia Sattler, and/or Dr. Winnie Teuteberg with questions.

**Supplemental Figure 1: Message to Primary Care clinicians soliciting surprise questions, staff message.**

Dear Dr. Wang,

Dr. Samantha Wang and Stanford's Serious Illness Conversation Program (SICP) are working to support Advance Care Planning in Hospital Medicine, using an AI model.

We need your help to make sure the tool identifies patients equitably!

Please reply to this message and indicate YES or NO for each of your patients below, based on your clinical judgment (no need to look at their chart if you know the patient well).

YES - I WOULD be surprised if this patient passed away in 1 year. (GOOD PROGNOSIS)

or

NO - I WOULD NOT be surprised if this patient passed away in 1 year. (POOR PROGNOSIS)

Please also mark an x by the 3 patients you would most prioritize for ACP.

| Name | MRN | Age | Bed | Team | Would I be surprised if pt passed away in 1 yr? | Top 3 for ACP Priority? |
|---|---|---|---|---|---|---|
| [redacted] | | | | MED UNIV 3 | | |
| | | | | MED UNIV 3 | | |
| | | | | MED UNIV 3 | | |
| | | | | MED UNIV 3 | | |
| | | | | MED UNIV 3 | | |
| | | | | MED UNIV 3 | | |
| | | | | MED UNIV 3 | | |
| | | | | MED UNIV 3 | | |
| | | | | MED UNIV 3 | | |
| | | | | MED UNIV 3 | | |
| | | | | MED UNIV 3 | | |
| | | | | MED UNIV 3 | | |

Thank you! Everyone will receive chocolate as appreciation for your time.

This study is supported by IRB 42078: Advanced Analytics to Improve Palliative Care Access.

Please reply to this message or staff message Jonathan Lu, Dr. Samantha Wang, and/or Dr. Winnie Teuteberg with questions.

**Supplemental Figure 2: Message to Hospital Medicine physicians soliciting surprise questions, email.**

### Supplemental Tables:

#### *Epic EOL Low Threshold in Primary Care*

| Group | Sample Size | Prevalence (Fraction) | Prevalence [95% CI] | Sensitivity (Fraction) | Sensitivity [95% CI] | Specificity (Fraction) | Specificity [95% CI] | Positive Predictive Value (Fraction) | Positive Predictive Value [95% CI] | O/E (Fraction) | O/E [95% CI] |
|---|---|---|---|---|---|---|---|---|---|---|---|
| Overall | 338 | 0.2 (68/338) | [0.16, 0.25] | 0.37 (25/68) | [0.26, 0.49] | 0.98 (265/270) | [0.97, 1.0] | 0.83 (25/30) | [0.7, 0.98] | 4.1 (68/16.4) | [3.3, 5.1] |
| Sex: Female | 201 | 0.19 (39/201) | [0.14, 0.26] | 0.38 (15/39) | [0.24, 0.53] | 0.98 (158/162) | [0.96, 1.0] | 0.79 (15/19) | [0.63, 0.99] | 4.0 (39/9.8) | [3.0, 5.3] |
| Sex: Male | 137 | 0.21 (29/137) | [0.15, 0.29] | 0.34 (10/29) | [0.15, 0.51] | 0.99 (107/108) | [0.98, 1.01] | 0.91 (10/11) | [0.82, 1.1] | 4.4 (29/6.6) | [3.2, 6.0] |

**Supplemental Table 1: Epic EOL Low Threshold in Primary Care: Reliability and Fairness Audit by Sex.** Significant differences in prevalence, significantly lower performance, or significantly higher O/E are bolded.

| Group | Sample Size | Prevalence (Fraction) | Prevalence [95% CI] | Sensitivity (Fraction) | Sensitivity [95% CI] | Specificity (Fraction) | Specificity [95% CI] | Positive Predictive Value (Fraction) | Positive Predictive Value [95% CI] | O/E (Fraction) | O/E [95% CI] |
|---|---|---|---|---|---|---|---|---|---|---|---|
| Overall | 338 | 0.2 (68/338) | [0.16, 0.25] | 0.37 (25/68) | [0.26, 0.49] | 0.98 (265/270) | [0.97, 1.0] | 0.83 (25/30) | [0.7, 0.98] | 4.1 (68/16.4) | [3.3, 5.1] |
| Age: (10, 20] | 3 | 0.0 (0/3) | [0, 0.71] | nan (0/0) | N/A | 1.0 (1/1) | [1.0, 1.0] | nan (0/0) | N/A | nan (0/0.0) | N/A |
| Age: (20, 30] | 27 | **0.0 (0/27)** | **[0, 0.13]** | nan (0/0) | N/A | 1.0 (1/1) | [1.0, 1.0] | nan (0/0) | N/A | nan (0/0.0) | N/A |
| Age: (30, 40] | 61 | **0.0 (0/61)** | **[0, 0.06]** | nan (0/0) | N/A | 1.0 (1/1) | [1.0, 1.0] | nan (0/0) | N/A | 0.0 (0/0.0) | N/A |
| Age: (40, 50] | 43 | 0.12 (5/43) | [0.04, 0.25] | 0.2 (1/5) | [-0.27, 0.4] | 1.0 (38/38) | [1.0, 1.0] | 1.0 (1/1) | [1.0, 1.0] | 11.1 (5/0.5) | [4.9, 25.3] |

| | | | | | | | | | | | |
|---|---|---|---|---|---|---|---|---|---|---|---|
| Age: (50, 60] | 48 | 0.04 (2/48) | [0.01, 0.14] | **0.0 (0/2)** | **[0.0, 0.0]** | 1.0 (46/46) | [1.0, 1.0] | nan (0/0) | N/A | 4.7 (2/0.4) | [1.2, 18.1] |
| Age: (60, 70] | 51 | 0.2 (10/51) | [0.1, 0.33] | **0.1 (1/10)** | **[-0.13, 0.2]** | 1.0 (41/41) | [1.0, 1.0] | 1.0 (1/1) | [1.0, 1.0] | **9.3 (10/1.1)** | **[5.3, 16.1]** |
| Age: (70, 80] | 51 | 0.29 (15/51) | [0.17, 0.44] | **0.07 (1/15)** | **[-0.09, 0.13]** | 0.97 (35/36) | [0.94, 1.03] | 0.5 (1/2) | [0.0, 1.0] | 6.3 (15/2.4) | [4.1, 9.6] |
| Age: (80, 90] | 33 | **0.55 (18/33)** | **[0.36, 0.72]** | 0.39 (7/18) | [0.15, 0.61] | 0.87 (13/15) | [0.73, 1.07] | 0.78 (7/9) | [0.56, 1.11] | 3.4 (18/5.3) | [2.5, 4.6] |
| Age: (90, 100] | 19 | **0.84 (16/19)** | **[0.6, 0.97]** | 0.81 (13/16) | [0.62, 1.0] | **0.33 (1/3)** | **[-0.33, 0.67]** | 0.87 (13/15) | [0.73, 1.05] | 2.7 (16/5.9) | [2.2, 3.3] |
| Age: (100, 110] | 2 | 1.0 (2/2) | [0.16, 1] | 1.0 (1/1) | [1.0, 1.0] | nan (0/0) | N/A | 1.0 (1/1) | [1.0, 1.0] | 2.4 (2/0.8) | [2.4, 2.4] |

**Supplemental Table 2: Epic EOL Low Threshold in Primary Care: Reliability and Fairness Audit by Age.** Significant differences in prevalence, significantly lower performance, or significantly higher O/E are bolded.

| Group | Sample Size | Prevalence (Fraction) | Prevalence [95% CI] | Sensitivity (Fraction) | Sensitivity [95% CI] | Specificity (Fraction) | Specificity [95% CI] | Positive Predictive Value (Fraction) | Positive Predictive Value [95% CI] | O/E (Fraction) | O/E [95% CI] |
|---|---|---|---|---|---|---|---|---|---|---|---|
| Overall | 338 | 0.2 (68/338) | [0.16, 0.25] | 0.37 (25/68) | [0.26, 0.49] | 0.98 (265/270) | [0.97, 1.0] | 0.83 (25/30) | [0.7, 0.98] | 4.1 (68/16.4) | [3.3, 5.1] |
| Ethnicity: Not Hispanic or Latino, Race: White | 154 | 0.25 (39/154) | [0.19, 0.33] | 0.44 (17/39) | [0.28, 0.59] | 0.97 (112/115) | [0.95, 1.01] | 0.85 (17/20) | [0.7, 1.01] | 4.0 (39/9.6) | [3.1, 5.3] |
| Ethnicity: Not Hispanic or Latino, Race: Asian | 103 | 0.19 (20/103) | [0.12, 0.28] | 0.3 (6/20) | [0.07, 0.49] | 0.99 (82/83) | [0.98, 1.01] | 0.86 (6/7) | [0.71, 1.21] | 4.7 (20/4.2) | [3.2, 7.0] |
| Ethnicity: Not Hispanic or Latino, Race: Other | 24 | 0.25 (6/24) | [0.1, 0.47] | 0.33 (2/6) | [-0.13, 0.67] | 0.94 (17/18) | [0.89, 1.08] | 0.67 (2/3) | [0.33, 1.33] | 3.1 (6/1.9) | [1.6, 6.2] |
| Ethnicity: Hispanic or Latino, Race: Other | 20 | 0.05 (1/20) | [0.0, 0.25] | **0.0 (0/1)** | **[0.0, 0.0]** | 1.0 (19/19) | [1.0, 1.0] | nan (0/0) | N/A | 4.2 (1/0.2) | [0.6, 28.1] |
| Ethnicity: Not Hispanic or Latino, Race: Black or | 9 | 0.0 (0/9) | [0, 0.34] | nan (0/0) | N/A | 1.0 (1/1) | [1.0, 1.0] | nan (0/0) | N/A | 0.0 | N/A |

| | | | | | | | | | | | |
|---|---|---|---|---|---|---|---|---|---|---|---|
| African American | | | | | | | | | | (0/0.2) | |
| Ethnicity: Hispanic or Latino, Race: White | 5 | 0.0 (0/5) | [0, 0.52] | nan (0/0) | N/A | 1.0 (1/1) | [1.0, 1.0] | nan (0/0) | N/A | 0.0 (0/0.0) | N/A |
| Ethnicity: Not Hispanic or Latino, Race: Native Hawaiian or Other Pacific Islander | 3 | 0.33 (1/3) | [0.01, 0.91] | **0.0 (0/1)** | **[0.0, 0.0]** | 1.0 (2/2) | [1.0, 1.0] | nan (0/0) | N/A | **50.0 (1/0.0)** | **[10.1, 247.7]** |
| Ethnicity: Not Hispanic or Latino, Race: Unknown | 3 | 0.0 (0/3) | [0, 0.71] | nan (0/0) | N/A | 1.0 (1/1) | [1.0, 1.0] | nan (0/0) | N/A | 0.0 (0/0.0) | N/A |
| Ethnicity: Hispanic or Latino, Race: Asian | 2 | 0.0 (0/2) | [0, 0.84] | nan (0/0) | N/A | 1.0 (1/1) | [1.0, 1.0] | nan (0/0) | N/A | nan (0/0.0) | N/A |
| Ethnicity: Not Hispanic or Latino, Race: American Indian or Alaska Native | 1 | 0.0 (0/1) | [0, 0.98] | nan (0/0) | N/A | 1.0 (1/1) | [1.0, 1.0] | nan (0/0) | N/A | 0.0 (0/0.0) | N/A |
| Ethnicity: Not Hispanic or Latino, Race: Patient Refused | 1 | 0.0 (0/1) | [0, 0.98] | nan (0/0) | N/A | 1.0 (1/1) | [1.0, 1.0] | nan (0/0) | N/A | nan (0/0.0) | N/A |

**Supplemental Table 3: Epic EOL Low Threshold in Primary Care: Reliability and Fairness Audit by Ethnicity/Race.** Significant differences in prevalence, significantly lower performance, or significantly higher O/E are bolded.

| Group | Sample Size | Prevalence (Fraction) | Prevalence [95% CI] | Sensitivity (Fraction) | Sensitivity [95% CI] | Specificity (Fraction) | Specificity [95% CI] | Positive Predictive Value (Fraction) | Positive Predictive Value [95% CI] | O/E (Fraction) | O/E [95% CI] |
|---|---|---|---|---|---|---|---|---|---|---|---|
| Overall | 338 | 0.2 (68/338) | [0.16, 0.25] | 0.37 (25/68) | [0.26, 0.49] | 0.98 (265/270) | [0.97, 1.0] | 0.83 (25/30) | [0.7, 0.98] | 4.1 (68/16.4) | [3.3, 5.1] |
| Ethnicity: Not Hispanic or Latino, Race: White, Sex: Female | 90 | 0.24 (22/90) | [0.16, 0.35] | 0.5 (11/22) | [0.29, 0.71] | 0.97 (66/68) | [0.94, 1.01] | 0.85 (11/13) | [0.69, 1.07] | 4.0 (22/5.5) | [2.8, 5.8] |
| Ethnicity: Not Hispanic or Latino, Race: White, Sex: Male | 64 | 0.27 (17/64) | [0.16, 0.39] | 0.35 (6/17) | [0.12, 0.56] | 0.98 (46/47) | [0.96, 1.03] | 0.86 (6/7) | [0.71, 1.14] | 4.1 (17/4.1) | [2.7, 6.2] |
| Ethnicity: Not Hispanic or Latino, Race: Asian, Sex: Female | 61 | 0.18 (11/61) | [0.09, 0.3] | 0.27 (3/11) | [-0.01, 0.55] | 0.98 (49/50) | [0.96, 1.02] | 0.75 (3/4) | [0.5, 1.5] | 4.2 (11/2.6) | [2.4, 7.1] |
| Ethnicity: Not Hispanic or Latino, Race: Asian, Sex: Male | 42 | 0.21 (9/42) | [0.1, 0.37] | 0.33 (3/9) | [0.0, 0.67] | 1.0 (33/33) | [1.0, 1.0] | 1.0 (3/3) | [1.0, 1.0] | 5.6 (9/1.6) | [3.2, 10.0] |

| | | | | | | | | | | | |
|---|---|---|---|---|---|---|---|---|---|---|---|
| Ethnicity: Not Hispanic or Latino, Race: Other, Sex: Female | 17 | 0.24 (4/17) | [0.07, 0.5] | 0.25 (1/4) | [-0.4, 0.5] | 0.92 (12/13) | [0.85, 1.1] | 0.5 (1/2) | [0.0, 1.0] | 3.0 (4/1.3) | [1.3, 7.0] |
| Ethnicity: Hispanic or Latino, Race: Other, Sex: Female | 11 | 0.0 (0/11) | [0, 0.28] | nan (0/0) | N/A | 1.0 (1/1) | [1.0, 1.0] | nan (0/0) | N/A | 0.0 (0/0.1) | N/A |
| Ethnicity: Hispanic or Latino, Race: Other, Sex: Male | 9 | 0.11 (1/9) | [0.0, 0.48] | **0.0 (0/1)** | **[0.0, 0.0]** | 1.0 (8/8) | [1.0, 1.0] | nan (0/0) | N/A | 7.1 (1/0.1) | [1.1, 45.3] |
| Ethnicity: Not Hispanic or Latino, Race: Other, Sex: Male | 7 | 0.29 (2/7) | [0.04, 0.71] | 0.5 (1/2) | [0.0, 1.0] | 1.0 (5/5) | [1.0, 1.0] | 1.0 (1/1) | [1.0, 1.0] | 3.4 (2/0.6) | [1.1, 11.1] |
| Ethnicity: Not Hispanic or Latino, Race: Black or African American, Sex: Female | 7 | 0.0 (0/7) | [0, 0.41] | nan (0/0) | N/A | 1.0 (1/1) | [1.0, 1.0] | nan (0/0) | N/A | 0.0 (0/0.2) | N/A |
| Ethnicity: Hispanic or Latino, Race: White, Sex: Male | 4 | 0.0 (0/4) | [0, 0.6] | nan (0/0) | N/A | 1.0 (1/1) | [1.0, 1.0] | nan (0/0) | N/A | 0.0 (0/0.0) | N/A |
| Ethnicity: Not Hispanic or Latino, Race: Native Hawaiian or Other Pacific Islander, Sex: Female | 3 | 0.33 (1/3) | [0.01, 0.91] | **0.0 (0/1)** | **[0.0, 0.0]** | 1.0 (2/2) | [1.0, 1.0] | nan (0/0) | N/A | **50.0 (1/0.0)** | **[10.1, 247.7]** |
| Ethnicity: Not Hispanic or Latino, Race: Black or African American, Sex: Male | 2 | 0.0 (0/2) | [0, 0.84] | nan (0/0) | N/A | 1.0 (1/1) | [1.0, 1.0] | nan (0/0) | N/A | 0.0 (0/0.0) | N/A |
| Ethnicity: Not Hispanic or Latino, Race: Unknown, Sex: Male | 2 | 0.0 (0/2) | [0, 0.84] | nan (0/0) | N/A | 1.0 (1/1) | [1.0, 1.0] | nan (0/0) | N/A | 0.0 (0/0.0) | N/A |
| Ethnicity: Hispanic or Latino, Race: Asian, Sex: Female | 2 | 0.0 (0/2) | [0, 0.84] | nan (0/0) | N/A | 1.0 (1/1) | [1.0, 1.0] | nan (0/0) | N/A | nan (0/0.0) | N/A |
| Ethnicity: Hispanic or Latino, Race: White, Sex: Female | 1 | 0.0 (0/1) | [0, 0.98] | nan (0/0) | N/A | 1.0 (1/1) | [1.0, 1.0] | nan (0/0) | N/A | nan (0/0.0) | N/A |
| Ethnicity: Not Hispanic or Latino, Race: Unknown, Sex: Female | 1 | 0.0 (0/1) | [0, 0.98] | nan (0/0) | N/A | 1.0 (1/1) | [1.0, 1.0] | nan (0/0) | N/A | nan (0/0.0) | N/A |
| Ethnicity: Not Hispanic or Latino, Race: American Indian or Alaska Native, Sex: Male | 1 | 0.0 (0/1) | [0, 0.98] | nan (0/0) | N/A | 1.0 (1/1) | [1.0, 1.0] | nan (0/0) | N/A | 0.0 (0/0.0) | N/A |
| Ethnicity: Not Hispanic or Latino, Race: Patient Refused, Sex: Male | 1 | 0.0 (0/1) | [0, 0.98] | nan (0/0) | N/A | 1.0 (1/1) | [1.0, 1.0] | nan (0/0) | N/A | nan (0/0.0) | N/A |

**Supplemental Table 4: Epic EOL Low Threshold in Primary Care: Reliability and Fairness Audit by Ethnicity/Race and Sex.** Significant differences in prevalence, significantly lower performance, or significantly higher O/E are bolded.

#### *Epic EOL High Threshold in Inpatient Oncology*

| Group | Sample Size | Prevalence (Fraction) | Prevalence [95% CI] | Sensitivity (Fraction) | Sensitivity [95% CI] | Specificity (Fraction) | Specificity [95% CI] | Positive Predictive Value (Fraction) | Positive Predictive Value [95% CI] | O/E (Fraction) | O/E [95% CI] |
|---|---|---|---|---|---|---|---|---|---|---|---|
| Overall | 150 | 0.7 (105/150) | [0.62, 0.77] | 0.27 (28/105) | [0.18, 0.34] | 0.91 (41/45) | [0.84, 1.0] | 0.88 (28/32) | [0.78, 1.01] | 3.0 (105/34.8) | [2.7, 3.4] |
| Sex: Female | 61 | 0.72 (44/61) | [0.59, 0.83] | 0.27 (12/44) | [0.14, 0.4] | 1.0 (17/17) | [1.0, 1.0] | 1.0 (12/12) | [1.0, 1.0] | 3.5 (44/12.7) | [3.0, 4.0] |
| Sex: Male | 89 | 0.69 (61/89) | [0.58, 0.78] | 0.26 (16/61) | [0.15, 0.37] | 0.86 (24/28) | [0.75, 0.99] | 0.8 (16/20) | [0.65, 1.0] | 2.8 (61/22.1) | [2.4, 3.2] |

**Supplemental Table 5: Epic EOL High Threshold in Inpatient Oncology: Reliability and Fairness Audit by Sex.** Prevalence, performance and calibration is presented for the overall cohort and for subgroups with significant differences in prevalence, significantly lower performance, or significantly higher O/E (bolded).

| Group | Sample Size | Prevalence (Fraction) | Prevalence [95% CI] | Sensitivity (Fraction) | Sensitivity [95% CI] | Specificity (Fraction) | Specificity [95% CI] | Positive Predictive Value (Fraction) | Positive Predictive Value [95% CI] | O/E (Fraction) | O/E [95% CI] |
|---|---|---|---|---|---|---|---|---|---|---|---|
| Overall | 150 | 0.7 (105/150) | [0.62, 0.77] | 0.27 (28/105) | [0.18, 0.34] | 0.91 (41/45) | [0.84, 1.0] | 0.88 (28/32) | [0.78, 1.01] | 3.0 (105/34.8) | [2.7, 3.4] |
| Age: (20, 30] | 13 | **0.23 (3/13)** | **[0.05, 0.54]** | **0.0 (0/3)** | **[0.0, 0.0]** | 1.0 (10/10) | [1.0, 1.0] | nan (0/0) | N/A | 5.0 (3/0.6) | [1.9, 13.5] |
| Age: (30, 40] | 14 | 0.57 (8/14) | [0.29, 0.82] | **0.0 (0/8)** | **[0.0, 0.0]** | 1.0 (6/6) | [1.0, 1.0] | nan (0/0) | N/A | **7.5 (8/1.1)** | **[4.8, 11.9]** |
| Age: (40, 50] | 14 | 0.57 (8/14) | [0.29, 0.82] | 0.12 (1/8) | [-0.15, 0.25] | 1.0 (6/6) | [1.0, 1.0] | 1.0 (1/1) | [1.0, 1.0] | 3.8 (8/2.1) | [2.4, 5.9] |
| Age: (50, 60] | 27 | 0.67 (18/27) | [0.46, 0.83] | 0.28 (5/18) | [0.06, 0.46] | 1.0 (9/9) | [1.0, 1.0] | 1.0 (5/5) | [1.0, 1.0] | 2.8 (18/6.5) | [2.1, 3.6] |

| | | | | | | | | | | | |
|---|---|---|---|---|---|---|---|---|---|---|---|
| Age: (60, 70] | 34 | 0.85 (29/34) | [0.69, 0.95] | 0.24 (7/29) | [0.07, 0.39] | **0.4 (2/5)** | **[-0.2, 0.8]** | 0.7 (7/10) | [0.4, 1.02] | 3.0 (29/9.8) | [2.6, 3.4] |
| Age: (70, 80] | 31 | 0.77 (24/31) | [0.59, 0.9] | 0.38 (9/24) | [0.18, 0.56] | 0.86 (6/7) | [0.71, 1.14] | 0.9 (9/10) | [0.8, 1.13] | 2.8 (24/8.4) | [2.4, 3.4] |
| Age: (80, 90] | 15 | 0.87 (13/15) | [0.6, 0.98] | 0.38 (5/13) | [0.13, 0.64] | 1.0 (2/2) | [1.0, 1.0] | 1.0 (5/5) | [1.0, 1.0] | 2.5 (13/5.2) | [2.0, 3.0] |
| Age: (90, 100] | 2 | 1.0 (2/2) | [0.16, 1] | 0.5 (1/2) | [0.0, 1.0] | nan (0/0) | N/A | 1.0 (1/1) | [1.0, 1.0] | 1.8 (2/1.1) | [1.8, 1.8] |

**Supplemental Table 6: Epic EOL High Threshold in Inpatient Oncology: Reliability and Fairness Audit by Age.** Significant differences in prevalence, significantly lower performance, or significantly higher O/E are bolded.

| Group | Sample Size | Prevalence (Fraction) | Prevalence [95% CI] | Sensitivity (Fraction) | Sensitivity [95% CI] | Specificity (Fraction) | Specificity [95% CI] | Positive Predictive Value (Fraction) | Positive Predictive Value [95% CI] | O/E (Fraction) | O/E [95% CI] |
|---|---|---|---|---|---|---|---|---|---|---|---|
| Overall | 150 | 0.7 (105/150) | [0.62, 0.77] | 0.27 (28/105) | [0.18, 0.34] | 0.91 (41/45) | [0.84, 1.0] | 0.88 (28/32) | [0.78, 1.01] | 3.0 (105/34.8) | [2.7, 3.4] |
| Ethnicity: Not Hispanic or Latino, Race: White | 57 | 0.67 (38/57) | [0.53, 0.79] | 0.24 (9/38) | [0.1, 0.37] | 0.84 (16/19) | [0.68, 1.03] | 0.75 (9/12) | [0.5, 1.04] | 2.7 (38/14.1) | [2.2, 3.2] |
| Ethnicity: Not Hispanic or Latino, Race: Asian | 38 | 0.82 (31/38) | [0.66, 0.92] | 0.39 (12/31) | [0.21, 0.56] | 1.0 (7/7) | [1.0, 1.0] | 1.0 (12/12) | [1.0, 1.0] | 2.6 (31/11.9) | [2.2, 3.0] |
| Ethnicity: Hispanic or Latino, Race: Other | 30 | 0.73 (22/30) | [0.54, 0.88] | **0.09 (2/22)** | **[-0.05, 0.18]** | 1.0 (8/8) | [1.0, 1.0] | 1.0 (2/2) | [1.0, 1.0] | **6.9 (22/3.2)** | **[5.6, 8.6]** |
| Ethnicity: Not Hispanic or Latino, Race: Other | 14 | 0.64 (9/14) | [0.35, 0.87] | 0.33 (3/9) | [0.0, 0.59] | 1.0 (5/5) | [1.0, 1.0] | 1.0 (3/3) | [1.0, 1.0] | 2.9 (9/3.1) | [2.0, 4.3] |
| Ethnicity: Not Hispanic or Latino, Race: Black or African American | 5 | 0.4 (2/5) | [0.05, 0.85] | 0.5 (1/2) | [0.0, 1.0] | 0.67 (2/3) | [0.33, 1.33] | 0.5 (1/2) | [0.0, 1.0] | 1.3 (2/1.5) | [0.4, 3.8] |
| Ethnicity: Hispanic or Latino, Race: White | 3 | 0.33 (1/3) | [0.01, 0.91] | **0.0 (0/1)** | **[0.0, 0.0]** | 1.0 (2/2) | [1.0, 1.0] | nan (0/0) | N/A | 2.9 (1/0.3) | [0.6, 14.6] |

| | | | | | | | | | | | |
|---|---|---|---|---|---|---|---|---|---|---|---|
| Ethnicity: Hispanic or Latino, Race: American Indian or Alaska Native | 1 | 0.0 (0/1) | [0, 0.98] | nan (0/0) | N/A | 1.0 (1/1) | [1.0, 1.0] | nan (0/0) | N/A | 0.0 (0/0.0) | N/A |
| Ethnicity: Not Hispanic or Latino, Race: American Indian or Alaska Native | 1 | 1.0 (1/1) | [0.03, 1] | 1.0 (1/1) | [1.0, 1.0] | nan (0/0) | N/A | 1.0 (1/1) | [1.0, 1.0] | 1.6 (1/0.6) | [1.6, 1.6] |

**Supplemental Table 7: Epic EOL High Threshold in Inpatient Oncology: Reliability and Fairness Audit by Ethnicity/Race.** Significant differences in prevalence, significantly lower performance, or significantly higher O/E are bolded.

| Group | Sample Size | Prevalence (Fraction) | Prevalence [95% CI] | Sensitivity (Fraction) | Sensitivity [95% CI] | Specificity (Fraction) | Specificity [95% CI] | Positive Predictive Value (Fraction) | Positive Predictive Value [95% CI] | O/E (Fraction) | O/E [95% CI] |
|---|---|---|---|---|---|---|---|---|---|---|---|
| Overall | 150 | 0.7 (105/150) | [0.62, 0.77] | 0.27 (28/105) | [0.18, 0.34] | 0.91 (41/45) | [0.84, 1.0] | 0.88 (28/32) | [0.78, 1.01] | 3.0 (105/34.8) | [2.7, 3.4] |
| Ethnicity: Not Hispanic or Latino, Race: White, Sex: Male | 32 | 0.69 (22/32) | [0.5, 0.84] | 0.23 (5/22) | [0.05, 0.38] | 0.7 (7/10) | [0.4, 1.02] | 0.62 (5/8) | [0.25, 1.0] | 2.7 (22/8.3) | [2.1, 3.4] |
| Ethnicity: Not Hispanic or Latino, Race: White, Sex: Female | 25 | 0.64 (16/25) | [0.43, 0.82] | 0.25 (4/16) | [0.03, 0.44] | 1.0 (9/9) | [1.0, 1.0] | 1.0 (4/4) | [1.0, 1.0] | 2.8 (16/5.8) | [2.1, 3.7] |
| Ethnicity: Not Hispanic or Latino, Race: Asian, Sex: Male | 23 | 0.83 (19/23) | [0.61, 0.95] | 0.37 (7/19) | [0.15, 0.58] | 1.0 (4/4) | [1.0, 1.0] | 1.0 (7/7) | [1.0, 1.0] | 2.4 (19/8.0) | [2.0, 2.9] |
| Ethnicity: Hispanic or Latino, Race: Other, Sex: Male | 17 | 0.76 (13/17) | [0.5, 0.93] | **0.0 (0/13)** | **[0.0, 0.0]** | 1.0 (4/4) | [1.0, 1.0] | nan (0/0) | N/A | **9.0 (13/1.4)** | **[6.9, 11.8]** |
| Ethnicity: Not Hispanic or Latino, Race: Asian, Sex: Female | 15 | 0.8 (12/15) | [0.52, 0.96] | 0.42 (5/12) | [0.13, 0.68] | 1.0 (3/3) | [1.0, 1.0] | 1.0 (5/5) | [1.0, 1.0] | 3.1 (12/3.9) | [2.4, 3.9] |
| Ethnicity: Hispanic or Latino, Race: Other, Sex: Female | 13 | 0.69 (9/13) | [0.39, 0.91] | 0.22 (2/9) | [-0.06, 0.44] | 1.0 (4/4) | [1.0, 1.0] | 1.0 (2/2) | [1.0, 1.0] | **5.2 (9/1.7)** | **[3.6, 7.4]** |
| Ethnicity: Not Hispanic or Latino, Race: Other, Sex: Male | 9 | 0.44 (4/9) | [0.14, 0.79] | 0.5 (2/4) | [0.0, 1.0] | 1.0 (5/5) | [1.0, 1.0] | 1.0 (2/2) | [1.0, 1.0] | 1.9 (4/2.1) | [0.9, 4.0] |
| Ethnicity: Not Hispanic or Latino, Race: Other, Sex: Female | 5 | 1.0 (5/5) | [0.48, 1] | 0.2 (1/5) | [-0.2, 0.4] | nan (0/0) | N/A | 1.0 (1/1) | [1.0, 1.0] | **4.9 (5/1.0)** | **[4.9, 4.9]** |
| Ethnicity: Not Hispanic or Latino, Race: Black or African American, Sex: Male | 3 | 0.33 (1/3) | [0.01, 0.91] | 1.0 (1/1) | [1.0, 1.0] | 0.5 (1/2) | [0.0, 1.0] | 0.5 (1/2) | [0.0, 1.0] | 0.8 (1/1.3) | [0.2, 3.8] |

| | | | | | | | | | | | |
|---|---|---|---|---|---|---|---|---|---|---|---|
| Ethnicity: Not Hispanic or Latino, Race: Black or African American, Sex: Female | 2 | 0.5 (1/2) | [0.01, 0.99] | **0.0 (0/1)** | **[0.0, 0.0]** | 1.0 (1/1) | [1.0, 1.0] | nan (0/0) | N/A | 4.2 (1/0.2) | [1.0, 16.7] |
| Ethnicity: Hispanic or Latino, Race: White, Sex: Male | 2 | 0.0 (0/2) | [0, 0.84] | nan (0/0) | N/A | 1.0 (1/1) | [1.0, 1.0] | nan (0/0) | N/A | 0.0 (0/0.3) | N/A |
| Ethnicity: Hispanic or Latino, Race: White, Sex: Female | 1 | 1.0 (1/1) | [0.03, 1] | **0.0 (0/1)** | **[0.0, 0.0]** | nan (0/0) | N/A | nan (0/0) | N/A | **inf (1/0.0)** | **[inf, inf]** |
| Ethnicity: Hispanic or Latino, Race: American Indian or Alaska Native, Sex: Male | 1 | 0.0 (0/1) | [0, 0.98] | nan (0/0) | N/A | 1.0 (1/1) | [1.0, 1.0] | nan (0/0) | N/A | 0.0 (0/0.0) | N/A |
| Ethnicity: Not Hispanic or Latino, Race: American Indian or Alaska Native, Sex: Male | 1 | 1.0 (1/1) | [0.03, 1] | 1.0 (1/1) | [1.0, 1.0] | nan (0/0) | N/A | 1.0 (1/1) | [1.0, 1.0] | 1.6 (1/0.6) | [1.6, 1.6] |

**Supplemental Table 8: Epic EOL High Threshold in Inpatient Oncology: Reliability and Fairness Audit by Ethnicity/Race and Sex.** Significant differences in prevalence, significantly lower performance, or significantly higher O/E are bolded.

#### *Stanford HM ACP in Inpatient Oncology*

| Group | Sample Size | Prevalence (Fraction) | Prevalence [95% CI] | Sensitivity (Fraction) | Sensitivity [95% CI] | Specificity (Fraction) | Specificity [95% CI] | Positive Predictive Value (Fraction) | Positive Predictive Value [95% CI] | O/E (Fraction) | O/E [95% CI] |
|---|---|---|---|---|---|---|---|---|---|---|---|
| Overall | 114 | 0.69 (79/114) | [0.6, 0.78] | 0.89 (70/79) | [0.82, 0.96] | 0.57 (20/35) | [0.4, 0.74] | 0.82 (70/85) | [0.74, 0.91] | 1.7 (79/46.2) | [1.5, 1.9] |
| Sex: Female | 48 | 0.67 (32/48) | [0.52, 0.8] | 0.91 (29/32) | [0.81, 1.01] | 0.5 (8/16) | [0.25, 0.75] | 0.78 (29/37) | [0.65, 0.91] | 1.6 (32/20.4) | [1.3, 1.9] |
| Sex: Male | 66 | 0.71 (47/66) | [0.59, 0.82] | 0.87 (41/47) | [0.78, 0.97] | 0.63 (12/19) | [0.43, 0.86] | 0.85 (41/48) | [0.77, 0.96] | 1.8 (47/25.7) | [1.6, 2.1] |

**Supplemental Table 9: Stanford HM ACP in Inpatient Oncology: Reliability and Fairness Audit by Sex.** Significant differences in prevalence, significantly lower performance, or significantly higher O/E are bolded.

| Group | Sample Size | Prevalence (Fraction) | Prevalence [95% CI] | Sensitivity (Fraction) | Sensitivity [95% CI] | Specificity (Fraction) | Specificity [95% CI] | Positive Predictive Value (Fraction) | Positive Predictive Value [95% CI] | O/E (Fraction) | O/E [95% CI] |
|---|---|---|---|---|---|---|---|---|---|---|---|
| Overall | 114 | 0.69 (79/114) | [0.6, 0.78] | 0.89 (70/79) | [0.82, 0.96] | 0.57 (20/35) | [0.4, 0.74] | 0.82 (70/85) | [0.74, 0.91] | 1.7 (79/46.2) | [1.5, 1.9] |
| Age: (20, 30] | 11 | 0.27 (3/11) | [0.06, 0.61] | 1.0 (3/3) | [1.0, 1.0] | 0.88 (7/8) | [0.75, 1.15] | 0.75 (3/4) | [0.5, 1.5] | 1.1 (3/2.7) | [0.4, 2.9] |
| Age: (30, 40] | 12 | 0.5 (6/12) | [0.21, 0.79] | 0.83 (5/6) | [0.67, 1.17] | 0.5 (3/6) | [0.0, 1.0] | 0.62 (5/8) | [0.25, 1.0] | 1.5 (6/4.0) | [0.8, 2.6] |
| Age: (40, 50] | 11 | 0.55 (6/11) | [0.23, 0.83] | 0.83 (5/6) | [0.67, 1.17] | **0.2 (1/5)** | **[-0.2, 0.4]** | 0.56 (5/9) | [0.24, 0.89] | 1.5 (6/4.0) | [0.9, 2.6] |
| Age: (50, 60] | 22 | 0.64 (14/22) | [0.41, 0.83] | 0.93 (13/14) | [0.86, 1.08] | 0.75 (6/8) | [0.5, 1.1] | 0.87 (13/15) | [0.73, 1.07] | 1.8 (14/7.8) | [1.3, 2.5] |
| Age: (60, 70] | 25 | 0.92 (23/25) | [0.74, 0.99] | 0.96 (22/23) | [0.91, 1.05] | 0.5 (1/2) | [0.0, 1.0] | 0.96 (22/23) | [0.91, 1.05] | 1.8 (23/12.6) | [1.6, 2.1] |
| Age: (70, 80] | 20 | 0.8 (16/20) | [0.56, 0.94] | 0.75 (12/16) | [0.56, 0.97] | 0.5 (2/4) | [0.0, 1.0] | 0.86 (12/14) | [0.71, 1.08] | 1.9 (16/8.3) | [1.5, 2.4] |
| Age: (80, 90] | 12 | 0.83 (10/12) | [0.52, 0.98] | 0.9 (9/10) | [0.8, 1.1] | **0.0 (0/2)** | **[0.0, 0.0]** | 0.82 (9/11) | [0.64, 1.05] | 1.6 (10/6.2) | [1.3, 2.1] |
| Age: (90, 100] | 1 | 1.0 (1/1) | [0.03, 1] | 1.0 (1/1) | [1.0, 1.0] | nan (0/0) | N/A | 1.0 (1/1) | [1.0, 1.0] | 1.6 (1/0.6) | [1.6, 1.6] |

**Supplemental Table 10: Stanford HM ACP in Inpatient Oncology: Reliability and Fairness Audit by Age.** Significant differences in prevalence, significantly lower performance, or significantly higher O/E are bolded.

| Group | Sample Size | Prevalence (Fraction) | Prevalence [95% CI] | Sensitivity (Fraction) | Sensitivity [95% CI] | Specificity (Fraction) | Specificity [95% CI] | Positive Predictive Value (Fraction) | Positive Predictive Value [95% CI] | O/E (Fraction) | O/E [95% CI] |
|---|---|---|---|---|---|---|---|---|---|---|---|
| Overall | 114 | 0.69 (79/114) | [0.6, 0.78] | 0.89 (70/79) | [0.82, 0.96] | 0.57 (20/35) | [0.4, 0.74] | 0.82 (70/85) | [0.74, 0.91] | 1.7 (79/46.2) | [1.5, 1.9] |

| | | | | | | | | | | | |
|---|---|---|---|---|---|---|---|---|---|---|---|
| Ethnicity: Not Hispanic or Latino, Race: White | 35 | 0.66 (23/35) | [0.48, 0.81] | 0.96 (22/23) | [0.91, 1.06] | 0.5 (6/12) | [0.22, 0.78] | 0.79 (22/28) | [0.64, 0.94] | 1.7 (23/13.7) | [1.3, 2.1] |
| Ethnicity: Not Hispanic or Latino, Race: Asian | 31 | 0.81 (25/31) | [0.63, 0.93] | 0.92 (23/25) | [0.84, 1.04] | 0.67 (4/6) | [0.33, 1.08] | 0.92 (23/25) | [0.84, 1.04] | 1.7 (25/14.8) | [1.4, 2.0] |
| Ethnicity: Hispanic or Latino, Race: Other | 26 | 0.73 (19/26) | [0.52, 0.88] | 0.84 (16/19) | [0.68, 1.03] | 0.71 (5/7) | [0.43, 1.1] | 0.89 (16/18) | [0.78, 1.04] | 1.8 (19/10.3) | [1.5, 2.3] |
| Ethnicity: Not Hispanic or Latino, Race: Other | 13 | 0.62 (8/13) | [0.32, 0.86] | 0.88 (7/8) | [0.75, 1.15] | 0.6 (3/5) | [0.2, 1.06] | 0.78 (7/9) | [0.56, 1.06] | 1.8 (8/4.5) | [1.1, 2.7] |
| Ethnicity: Hispanic or Latino, Race: White | 3 | 0.33 (1/3) | [0.01, 0.91] | 1.0 (1/1) | [1.0, 1.0] | **0.0 (0/2)** | **[0.0, 0.0]** | **0.33 (1/3)** | **[-0.33, 0.67]** | 0.8 (1/1.3) | [0.2, 3.9] |
| Ethnicity: Not Hispanic or Latino, Race: Black or African American | 3 | 0.33 (1/3) | [0.01, 0.91] | **0.0 (0/1)** | **[0.0, 0.0]** | 0.5 (1/2) | [0.0, 1.0] | **0.0 (0/1)** | **[0.0, 0.0]** | 1.8 (1/0.6) | [0.4, 8.8] |
| Ethnicity: Hispanic or Latino, Race: American Indian or Alaska Native | 1 | 0.0 (0/1) | [0, 0.98] | nan (0/0) | N/A | 1.0 (1/1) | [1.0, 1.0] | nan (0/0) | N/A | 0.0 (0/0.1) | N/A |
| Ethnicity: Not Hispanic or Latino, Race: American Indian or Alaska Native | 1 | 1.0 (1/1) | [0.03, 1] | **0.0 (0/1)** | **[0.0, 0.0]** | nan (0/0) | N/A | nan (0/0) | N/A | **4.1 (1/0.2)** | **[4.1, 4.1]** |

**Supplemental Table 11: Stanford HM ACP in Inpatient Oncology: Reliability and Fairness Audit by Ethnicity/Race.** Significant differences in prevalence, significantly lower performance, or significantly higher O/E are bolded.

| Group | Sample Size | Prevalence (Fraction) | Prevalence [95% CI] | Sensitivity (Fraction) | Sensitivity [95% CI] | Specificity (Fraction) | Specificity [95% CI] | Positive Predictive Value (Fraction) | Positive Predictive Value [95% CI] | O/E (Fraction) | O/E [95% CI] |
|---|---|---|---|---|---|---|---|---|---|---|---|
| Overall | 114 | 0.69 (79/114) | [0.6, 0.78] | 0.89 (70/79) | [0.82, 0.96] | 0.57 (20/35) | [0.4, 0.74] | 0.82 (70/85) | [0.74, 0.91] | 1.7 (79/46.2) | [1.5, 1.9] |
| Ethnicity: Not Hispanic or Latino, Race: Asian, Sex: Male | 19 | 0.84 (16/19) | [0.6, 0.97] | 0.94 (15/16) | [0.88, 1.08] | 0.67 (2/3) | [0.33, 1.33] | 0.94 (15/16) | [0.88, 1.08] | 1.9 (16/8.4) | [1.6, 2.3] |
| Ethnicity: Not Hispanic or Latino, Race: White, Sex: Male | 18 | 0.78 (14/18) | [0.52, 0.94] | 0.93 (13/14) | [0.86, 1.09] | 0.75 (3/4) | [0.5, 1.3] | 0.93 (13/14) | [0.86, 1.07] | 1.9 (14/7.4) | [1.5, 2.4] |

| | | | | | | | | | | | |
|---|---|---|---|---|---|---|---|---|---|---|---|
| Ethnicity: Not Hispanic or Latino, Race: White, Sex: Female | 17 | 0.53 (9/17) | [0.28, 0.77] | 1.0 (9/9) | [1.0, 1.0] | 0.38 (3/8) | [-0.03, 0.75] | 0.64 (9/14) | [0.38, 0.9] | 1.4 (9/6.3) | [0.9, 2.2] |
| Ethnicity: Hispanic or Latino, Race: Other, Sex: Male | 15 | 0.8 (12/15) | [0.52, 0.96] | 0.83 (10/12) | [0.67, 1.07] | 0.67 (2/3) | [0.33, 1.33] | 0.91 (10/11) | [0.82, 1.12] | 1.9 (12/6.2) | [1.5, 2.5] |
| Ethnicity: Not Hispanic or Latino, Race: Asian, Sex: Female | 12 | 0.75 (9/12) | [0.43, 0.95] | 0.89 (8/9) | [0.78, 1.11] | 0.67 (2/3) | [0.33, 1.33] | 0.89 (8/9) | [0.78, 1.11] | 1.4 (9/6.4) | [1.0, 1.9] |
| Ethnicity: Hispanic or Latino, Race: Other, Sex: Female | 11 | 0.64 (7/11) | [0.31, 0.89] | 0.86 (6/7) | [0.71, 1.16] | 0.75 (3/4) | [0.5, 1.25] | 0.86 (6/7) | [0.71, 1.21] | 1.7 (7/4.2) | [1.1, 2.6] |
| Ethnicity: Not Hispanic or Latino, Race: Other, Sex: Male | 8 | 0.38 (3/8) | [0.09, 0.76] | 0.67 (2/3) | [0.33, 1.33] | 0.6 (3/5) | [0.2, 1.2] | 0.5 (2/4) | [0.0, 1.0] | 1.4 (3/2.1) | [0.6, 3.5] |
| Ethnicity: Not Hispanic or Latino, Race: Other, Sex: Female | 5 | 1.0 (5/5) | [0.48, 1] | 1.0 (1/1) | [1.0, 1.0] | nan (0/0) | N/A | 1.0 (1/1) | [1.0, 1.0] | **2.0 (5/2.4)** | **[2.0, 2.0]** |
| Ethnicity: Hispanic or Latino, Race: White, Sex: Male | 2 | 0.0 (0/2) | [0, 0.84] | nan (0/0) | N/A | **0.0 (0/2)** | **[0.0, 0.0]** | **0.0 (0/2)** | **[0.0, 0.0]** | 0.0 (0/0.5) | N/A |
| Ethnicity: Not Hispanic or Latino, Race: Black or African American, Sex: Female | 2 | 0.5 (1/2) | [0.01, 0.99] | **0.0 (0/1)** | **[0.0, 0.0]** | **0.0 (0/1)** | **[0.0, 0.0]** | **0.0 (0/1)** | **[0.0, 0.0]** | 2.6 (1/0.4) | [0.7, 10.6] |
| Ethnicity: Hispanic or Latino, Race: White, Sex: Female | 1 | 1.0 (1/1) | [0.03, 1] | 1.0 (1/1) | [1.0, 1.0] | nan (0/0) | N/A | 1.0 (1/1) | [1.0, 1.0] | 1.4 (1/0.7) | [1.4, 1.4] |
| Ethnicity: Not Hispanic or Latino, Race: Black or African American, Sex: Male | 1 | 0.0 (0/1) | [0, 0.98] | nan (0/0) | N/A | 1.0 (1/1) | [1.0, 1.0] | nan (0/0) | N/A | 0.0 (0/0.2) | N/A |
| Ethnicity: Hispanic or Latino, Race: American Indian or Alaska Native, Sex: Male | 1 | 0.0 (0/1) | [0, 0.98] | nan (0/0) | N/A | 1.0 (1/1) | [1.0, 1.0] | nan (0/0) | N/A | 0.0 (0/0.1) | N/A |
| Ethnicity: Not Hispanic or Latino, Race: American Indian or Alaska Native, Sex: Male | 1 | 1.0 (1/1) | [0.03, 1] | **0.0 (0/1)** | **[0.0, 0.0]** | nan (0/0) | N/A | nan (0/0) | N/A | **4.1 (1/0.2)** | **[4.1, 4.1]** |

**Supplemental Table 12: Stanford HM ACP in Inpatient Oncology: Reliability and Fairness Audit by Ethnicity/Race and Sex.** Significant differences in prevalence, significantly lower performance, or significantly higher O/E are bolded.

#### *Epic EOL High Threshold in Hospital Medicine*

| Group | Sample Size | Prevalence (Fraction) | Prevalence [95% CI] | Sensitivity (Fraction) | Sensitivity [95% CI] | Specificity (Fraction) | Specificity [95% CI] | Positive Predictive Value (Fraction) | Positive Predictive Value [95% CI] | O/E (Fraction) | O/E [95% CI] |
|---|---|---|---|---|---|---|---|---|---|---|---|
| Overall | 305 | 0.44 (133/305) | [0.38, 0.49] | 0.2 (26/133) | [0.12, 0.26] | 0.95 (164/172) | [0.92, 0.99] | 0.76 (26/34) | [0.63, 0.91] | 2.5 (133/53.2) | [2.2, 2.8] |
| Sex: Female | 140 | 0.47 (66/140) | [0.39, 0.56] | 0.26 (17/66) | [0.14, 0.36] | 0.95 (70/74) | [0.9, 1.0] | 0.81 (17/21) | [0.67, 0.99] | 2.6 (66/25.2) | [2.2, 3.1] |
| Sex: Male | 165 | 0.41 (67/165) | [0.33, 0.49] | 0.13 (9/67) | [0.05, 0.21] | 0.96 (94/98) | [0.93, 1.01] | 0.69 (9/13) | [0.45, 0.97] | 2.4 (67/28.0) | [2.0, 2.9] |

**Supplemental Table 13: Epic EOL High Threshold in Hospital Medicine: Reliability and Fairness Audit by Sex.** Significant differences in prevalence, significantly lower performance, or significantly higher O/E are bolded.

| Group | Sample Size | Prevalence (Fraction) | Prevalence [95% CI] | Sensitivity (Fraction) | Sensitivity [95% CI] | Specificity (Fraction) | Specificity [95% CI] | Positive Predictive Value (Fraction) | Positive Predictive Value [95% CI] | O/E (Fraction) | O/E [95% CI] |
|---|---|---|---|---|---|---|---|---|---|---|---|
| Overall | 305 | 0.44 (133/305) | [0.38, 0.49] | 0.2 (26/133) | [0.12, 0.26] | 0.95 (164/172) | [0.92, 0.99] | 0.76 (26/34) | [0.63, 0.91] | 2.5 (133/53.2) | [2.2, 2.8] |
| Age: (10, 20] | 3 | 0.33 (1/3) | [0.01, 0.91] | **0.0 (0/1)** | **[0.0, 0.0]** | 1.0 (2/2) | [1.0, 1.0] | nan (0/0) | N/A | **inf (1/0.0)** | **[inf, inf]** |
| Age: (20, 30] | 24 | **0.12 (3/24)** | **[0.03, 0.32]** | **0.0 (0/3)** | **[0.0, 0.0]** | 1.0 (21/21) | [1.0, 1.0] | nan (0/0) | N/A | 4.5 (3/0.7) | [1.6, 12.9] |
| Age: (30, 40] | 40 | **0.15 (6/40)** | **[0.06, 0.3]** | **0.0 (0/6)** | **[0.0, 0.0]** | 1.0 (34/34) | [1.0, 1.0] | nan (0/0) | N/A | 4.7 (6/1.3) | [2.2, 9.7] |
| Age: (40, 50] | 12 | 0.5 (6/12) | [0.21, 0.79] | 0.17 (1/6) | [-0.17, 0.33] | 1.0 (6/6) | [1.0, 1.0] | 1.0 (1/1) | [1.0, 1.0] | 3.6 (6/1.7) | [2.0, 6.3] |
| Age: (50, 60] | 40 | 0.28 (11/40) | [0.15, 0.44] | **0.0 (0/11)** | **[0.0, 0.0]** | 1.0 (29/29) | [1.0, 1.0] | nan (0/0) | N/A | 3.6 (11/3.1) | [2.2, 5.9] |
| Age: (60, 70] | 72 | 0.44 (32/72) | [0.33, 0.57] | 0.19 (6/32) | [0.04, 0.31] | 0.92 (37/40) | [0.85, 1.01] | 0.67 (6/9) | [0.33, 1.0] | 2.2 (32/14.7) | [1.7, 2.8] |
| Age: (70, 80] | 62 | 0.5 (31/62) | [0.37, 0.63] | 0.32 (10/31) | [0.16, 0.48] | 0.9 (28/31) | [0.81, 1.02] | 0.77 (10/13) | [0.54, 1.04] | 2.2 (31/14.2) | [1.7, 2.8] |

| | | | | | | | | | | | |
|---|---|---|---|---|---|---|---|---|---|---|---|
| Age: (80, 90] | 34 | **0.76 (26/34)** | **[0.59, 0.89]** | 0.19 (5/26) | [0.02, 0.34] | 0.88 (7/8) | [0.75, 1.18] | 0.83 (5/6) | [0.67, 1.17] | 2.6 (26/10.0) | [2.2, 3.1] |
| Age: (90, 100] | 18 | **0.94 (17/18)** | **[0.73, 1.0]** | 0.24 (4/17) | [0.03, 0.41] | **0.0 (0/1)** | **[0.0, 0.0]** | 0.8 (4/5) | [0.6, 1.27] | 2.2 (17/7.6) | [2.0, 2.5] |

**Supplemental Table 14: Epic EOL High Threshold in Hospital Medicine: Reliability and Fairness Audit by Age.** Significant differences in prevalence, significantly lower performance, or significantly higher O/E are bolded.

| Group | Sample Size | Prevalence (Fraction) | Prevalence [95% CI] | Sensitivity (Fraction) | Sensitivity [95% CI] | Specificity (Fraction) | Specificity [95% CI] | Positive Predictive Value (Fraction) | Positive Predictive Value [95% CI] | O/E (Fraction) | O/E [95% CI] |
|---|---|---|---|---|---|---|---|---|---|---|---|
| Overall | 305 | 0.44 (133/305) | [0.38, 0.49] | 0.2 (26/133) | [0.12, 0.26] | 0.95 (164/172) | [0.92, 0.99] | 0.76 (26/34) | [0.63, 0.91] | 2.5 (133/53.2) | [2.2, 2.8] |
| Ethnicity: Not Hispanic or Latino, Race: White | 145 | 0.44 (64/145) | [0.36, 0.53] | 0.09 (6/64) | [0.01, 0.16] | 0.95 (77/81) | [0.91, 1.0] | 0.6 (6/10) | [0.27, 0.95] | 3.1 (64/20.4) | [2.6, 3.8] |
| Ethnicity: Hispanic or Latino, Race: Other | 44 | **0.18 (8/44)** | **[0.08, 0.33]** | 0.12 (1/8) | [-0.17, 0.25] | 0.94 (34/36) | [0.89, 1.02] | 0.33 (1/3) | [-0.33, 0.67] | 2.0 (8/4.0) | [1.1, 3.7] |
| Ethnicity: Not Hispanic or Latino, Race: Asian | 37 | **0.68 (25/37)** | **[0.5, 0.82]** | 0.32 (8/25) | [0.12, 0.52] | 1.0 (12/12) | [1.0, 1.0] | 1.0 (8/8) | [1.0, 1.0] | 2.1 (25/12.2) | [1.6, 2.6] |
| Ethnicity: Not Hispanic or Latino, Race: Black or African American | 35 | 0.54 (19/35) | [0.37, 0.71] | 0.47 (9/19) | [0.26, 0.7] | 0.88 (14/16) | [0.75, 1.06] | 0.82 (9/11) | [0.64, 1.08] | 1.6 (19/12.2) | [1.2, 2.1] |
| Ethnicity: Not Hispanic or Latino, Race: Other | 16 | 0.5 (8/16) | [0.25, 0.75] | 0.25 (2/8) | [-0.1, 0.5] | 1.0 (8/8) | [1.0, 1.0] | 1.0 (2/2) | [1.0, 1.0] | 4.0 (8/2.0) | [2.4, 6.5] |
| Ethnicity: Hispanic or Latino, Race: White | 13 | 0.23 (3/13) | [0.05, 0.54] | **0.0 (0/3)** | **[0.0, 0.0]** | 1.0 (10/10) | [1.0, 1.0] | nan (0/0) | N/A | 4.4 (3/0.7) | [1.6, 11.9] |
| Ethnicity: Not Hispanic or Latino, Race: Native Hawaiian or Other Pacific Islander | 10 | 0.4 (4/10) | [0.12, 0.74] | **0.0 (0/4)** | **[0.0, 0.0]** | 1.0 (6/6) | [1.0, 1.0] | nan (0/0) | N/A | 3.0 (4/1.3) | [1.4, 6.4] |
| Ethnicity: Not Hispanic or Latino, Race: American Indian or Alaska Native | 1 | 0.0 (0/1) | [0, 0.98] | nan (0/0) | N/A | 1.0 (1/1) | [1.0, 1.0] | nan (0/0) | N/A | 0.0 (0/0.2) | N/A |

**Supplemental Table 15: Epic EOL High Threshold in Hospital Medicine: Reliability and Fairness Audit by Ethnicity/Race.** Significant differences in prevalence, significantly lower performance, or significantly higher O/E are bolded.

| Group | Sample Size | Prevalence (Fraction) | Prevalence [95% CI] | Sensitivity (Fraction) | Sensitivity [95% CI] | Specificity (Fraction) | Specificity [95% CI] | Positive Predictive Value (Fraction) | Positive Predictive Value [95% CI] | O/E (Fraction) | O/E [95% CI] |
|---|---|---|---|---|---|---|---|---|---|---|---|
| Overall | 305 | 0.44 (133/305) | [0.38, 0.49] | 0.2 (26/133) | [0.12, 0.26] | 0.95 (164/172) | [0.92, 0.99] | 0.76 (26/34) | [0.63, 0.91] | 2.5 (133/53.2) | [2.2, 2.8] |
| Ethnicity: Not Hispanic or Latino, Race: White, Sex: Male | 81 | 0.38 (31/81) | [0.28, 0.5] | 0.06 (2/31) | [-0.04, 0.13] | 0.94 (47/50) | [0.88, 1.01] | 0.4 (2/5) | [-0.2, 0.8] | 2.7 (31/11.4) | [2.1, 3.6] |
| Ethnicity: Not Hispanic or Latino, Race: White, Sex: Female | 64 | 0.52 (33/64) | [0.39, 0.64] | 0.12 (4/33) | [0.01, 0.22] | 0.97 (30/31) | [0.94, 1.04] | 0.8 (4/5) | [0.6, 1.27] | **3.7 (33/9.0)** | **[2.9, 4.6]** |
| Ethnicity: Hispanic or Latino, Race: Other, Sex: Female | 22 | 0.23 (5/22) | [0.08, 0.45] | 0.2 (1/5) | [-0.27, 0.4] | 0.88 (15/17) | [0.76, 1.05] | 0.33 (1/3) | [-0.33, 0.67] | 1.5 (5/3.4) | [0.7, 3.2] |
| Ethnicity: Hispanic or Latino, Race: Other, Sex: Male | 22 | **0.14 (3/22)** | **[0.03, 0.35]** | **0.0 (0/3)** | **[0.0, 0.0]** | 1.0 (19/19) | [1.0, 1.0] | nan (0/0) | N/A | 4.3 (3/0.7) | [1.5, 12.4] |
| Ethnicity: Not Hispanic or Latino, Race: Asian, Sex: Male | 21 | 0.71 (15/21) | [0.48, 0.89] | 0.13 (2/15) | [-0.07, 0.27] | 1.0 (6/6) | [1.0, 1.0] | 1.0 (2/2) | [1.0, 1.0] | 2.1 (15/7.3) | [1.6, 2.7] |
| Ethnicity: Not Hispanic or Latino, Race: Black or African American, Sex: Male | 20 | 0.45 (9/20) | [0.23, 0.68] | 0.56 (5/9) | [0.24, 0.91] | 0.91 (10/11) | [0.82, 1.15] | 0.83 (5/6) | [0.67, 1.22] | 1.3 (9/6.8) | [0.8, 2.2] |
| Ethnicity: Not Hispanic or Latino, Race: Asian, Sex: Female | 16 | 0.62 (10/16) | [0.35, 0.85] | 0.6 (6/10) | [0.3, 0.91] | 1.0 (6/6) | [1.0, 1.0] | 1.0 (6/6) | [1.0, 1.0] | 2.0 (10/4.9) | [1.4, 3.0] |
| Ethnicity: Not Hispanic or Latino, Race: Black or African American, Sex: Female | 15 | 0.67 (10/15) | [0.38, 0.88] | 0.4 (4/10) | [0.09, 0.7] | 0.8 (4/5) | [0.6, 1.27] | 0.8 (4/5) | [0.6, 1.27] | 1.9 (10/5.4) | [1.3, 2.7] |
| Ethnicity: Not Hispanic or Latino, Race: Other, Sex: Male | 9 | 0.56 (5/9) | [0.21, 0.86] | **0.0 (0/5)** | **[0.0, 0.0]** | 1.0 (4/4) | [1.0, 1.0] | nan (0/0) | N/A | **13.9 (5/0.4)** | **[7.7, 24.9]** |
| Ethnicity: Not Hispanic or Latino, Race: Other, Sex: Female | 7 | 0.43 (3/7) | [0.1, 0.82] | 0.67 (2/3) | [0.33, 1.33] | 1.0 (4/4) | [1.0, 1.0] | 1.0 (2/2) | [1.0, 1.0] | 1.8 (3/1.7) | [0.8, 4.3] |
| Ethnicity: Hispanic or Latino, Race: White, Sex: Male | 7 | 0.14 (1/7) | [0.0, 0.58] | **0.0 (0/1)** | **[0.0, 0.0]** | 1.0 (6/6) | [1.0, 1.0] | nan (0/0) | N/A | 10.0 | [1.6, |

| | | | | | | | | | | | |
|---|---|---|---|---|---|---|---|---|---|---|---|
| | | | | | | | | | | (1/0.1) | 61.4] |
| Ethnicity: Hispanic or Latino, Race: White, Sex: Female | 6 | 0.33 (2/6) | [0.04, 0.78] | **0.0 (0/2)** | **[0.0, 0.0]** | 1.0 (4/4) | [1.0, 1.0] | nan (0/0) | N/A | 3.4 (2/0.6) | [1.1, 10.7] |
| Ethnicity: Not Hispanic or Latino, Race: Native Hawaiian or Other Pacific Islander, Sex: Female | 6 | 0.17 (1/6) | [0.0, 0.64] | **0.0 (0/1)** | **[0.0, 0.0]** | 1.0 (5/5) | [1.0, 1.0] | nan (0/0) | N/A | 7.7 (1/0.1) | [1.3, 46.0] |
| Ethnicity: Not Hispanic or Latino, Race: Native Hawaiian or Other Pacific Islander, Sex: Male | 4 | 0.75 (3/4) | [0.19, 0.99] | **0.0 (0/3)** | **[0.0, 0.0]** | 1.0 (1/1) | [1.0, 1.0] | nan (0/0) | N/A | 2.5 (3/1.2) | [1.4, 4.4] |
| Ethnicity: Not Hispanic or Latino, Race: American Indian or Alaska Native, Sex: Male | 1 | 0.0 (0/1) | [0, 0.98] | nan (0/0) | N/A | 1.0 (1/1) | [1.0, 1.0] | nan (0/0) | N/A | 0.0 (0/0.2) | N/A |

**Supplemental Table 16: Epic EOL High Threshold in Hospital Medicine: Reliability and Fairness Audit by Ethnicity/Race and Sex.** Significant differences in prevalence, significantly lower performance, or significantly higher O/E are bolded.

#### *Stanford HM ACP in Hospital Medicine*

| Group | Sample Size | Prevalence (Fraction) | Prevalence [95% CI] | Sensitivity (Fraction) | Sensitivity [95% CI] | Specificity (Fraction) | Specificity [95% CI] | Positive Predictive Value (Fraction) | Positive Predictive Value [95% CI] | O/E (Fraction) | O/E [95% CI] |
|---|---|---|---|---|---|---|---|---|---|---|---|
| Overall | 225 | 0.44 (99/225) | [0.37, 0.51] | 0.69 (68/99) | [0.6, 0.78] | 0.87 (109/126) | [0.81, 0.93] | 0.8 (68/85) | [0.72, 0.89] | 1.5 (99/65.2) | [1.3, 1.8] |
| Sex: Female | 114 | 0.45 (51/114) | [0.35, 0.54] | 0.73 (37/51) | [0.61, 0.86] | 0.83 (52/63) | [0.73, 0.92] | 0.77 (37/48) | [0.66, 0.9] | 1.4 (51/35.7) | [1.2, 1.7] |
| Sex: Male | 111 | 0.43 (48/111) | [0.34, 0.53] | 0.65 (31/48) | [0.51, 0.78] | 0.9 (57/63) | [0.84, 0.98] | 0.84 (31/37) | [0.73, 0.96] | 1.6 (48/29.5) | [1.3, 2.0] |

**Supplemental Table 17: Stanford HM ACP in Hospital Medicine: Reliability and Fairness Audit by Sex.** Significant differences in prevalence, significantly lower performance, or significantly higher O/E are bolded.

| Group | Sample Size | Prevalence (Fraction) | Prevalence [95% CI] | Sensitivity (Fraction) | Sensitivity [95% CI] | Specificity (Fraction) | Specificity [95% CI] | Positive Predictive Value (Fraction) | Positive Predictive Value [95% CI] | O/E (Fraction) | O/E [95% CI] |
|---|---|---|---|---|---|---|---|---|---|---|---|

| | | | | | | | | | | | |
|---|---|---|---|---|---|---|---|---|---|---|---|
| Overall | 225 | 0.44 (99/225) | [0.37, 0.51] | 0.69 (68/99) | [0.6, 0.78] | 0.87 (109/126) | [0.81, 0.93] | 0.8 (68/85) | [0.72, 0.89] | 1.5 (99/65.2) | [1.3, 1.8] |
| Age: (10, 20] | 3 | 0.33 (1/3) | [0.01, 0.91] | **0.0 (0/1)** | **[0.0, 0.0]** | 1.0 (2/2) | [1.0, 1.0] | nan (0/0) | N/A | 2.4 (1/0.4) | [0.5, 12.1] |
| Age: (20, 30] | 13 | 0.15 (2/13) | [0.02, 0.45] | 0.5 (1/2) | [0.0, 1.0] | 1.0 (11/11) | [1.0, 1.0] | 1.0 (1/1) | [1.0, 1.0] | 1.0 (2/2.0) | [0.3, 3.7] |
| Age: (30, 40] | 28 | **0.11 (3/28)** | **[0.02, 0.28]** | **0.0 (0/3)** | **[0.0, 0.0]** | 1.0 (25/25) | [1.0, 1.0] | nan (0/0) | N/A | 0.8 (3/3.7) | [0.3, 2.3] |
| Age: (40, 50] | 15 | 0.4 (6/15) | [0.16, 0.68] | 0.33 (2/6) | [-0.08, 0.67] | 1.0 (9/9) | [1.0, 1.0] | 1.0 (2/2) | [1.0, 1.0] | 1.8 (6/3.4) | [1.0, 3.3] |
| Age: (50, 60] | 25 | 0.24 (6/25) | [0.09, 0.45] | **0.17 (1/6)** | **[-0.17, 0.33]** | 1.0 (19/19) | [1.0, 1.0] | 1.0 (1/1) | [1.0, 1.0] | 1.5 (6/4.1) | [0.7, 2.9] |
| Age: (60, 70] | 50 | 0.36 (18/50) | [0.23, 0.51] | 0.78 (14/18) | [0.6, 0.98] | 0.84 (27/32) | [0.73, 0.98] | 0.74 (14/19) | [0.56, 0.95] | 1.2 (18/14.9) | [0.8, 1.8] |
| Age: (70, 80] | 48 | 0.56 (27/48) | [0.41, 0.71] | 0.81 (22/27) | [0.67, 0.99] | **0.57 (12/21)** | **[0.37, 0.78]** | 0.71 (22/31) | [0.56, 0.88] | 1.3 (27/20.2) | [1.0, 1.7] |
| Age: (80, 90] | 30 | **0.8 (24/30)** | **[0.61, 0.92]** | 0.75 (18/24) | [0.59, 0.93] | 0.5 (3/6) | [0.0, 1.0] | 0.86 (18/21) | [0.71, 1.01] | 2.1 (24/11.7) | [1.7, 2.5] |
| Age: (90, 100] | 13 | **0.92 (12/13)** | **[0.64, 1.0]** | 0.83 (10/12) | [0.67, 1.05] | 1.0 (1/1) | [1.0, 1.0] | 1.0 (10/10) | [1.0, 1.0] | **2.5 (12/4.9)** | **[2.1, 2.9]** |

**Supplemental Table 18: Stanford HM ACP in Hospital Medicine: Reliability and Fairness Audit by Age.** Significant differences in prevalence, significantly lower performance, or significantly higher O/E are bolded.

| Group | Sample Size | Prevalence (Fraction) | Prevalence [95% CI] | Sensitivity (Fraction) | Sensitivity [95% CI] | Specificity (Fraction) | Specificity [95% CI] | Positive Predictive Value (Fraction) | Positive Predictive Value [95% CI] | O/E (Fraction) | O/E [95% CI] |
|---|---|---|---|---|---|---|---|---|---|---|---|
| Overall | 225 | 0.44 (99/225) | [0.37, 0.51] | 0.69 (68/99) | [0.6, 0.78] | 0.87 (109/126) | [0.81, 0.93] | 0.8 (68/85) | [0.72, 0.89] | 1.5 (99/65.2) | [1.3, 1.8] |
| Ethnicity: Not Hispanic or Latino, Race: White | 81 | 0.44 (36/81) | [0.33, 0.56] | 0.67 (24/36) | [0.51, 0.81] | 0.91 (41/45) | [0.84, 1.0] | 0.86 (24/28) | [0.74, 1.0] | 1.8 | [1.4, |

| | | | | | | | | | | | |
|---|---|---|---|---|---|---|---|---|---|---|---|
| | | | | | | | | | | (36/20.2 ) | 2.3] |
| Ethnicity: Hispanic or Latino, Race: Other | 38 | **0.16 (6/38)** | **[0.06, 0.31]** | 0.33 (2/6) | [-0.13, 0.67] | 0.84 (27/32) | [0.72, 0.96] | **0.29 (2/7)** | **[-0.1, 0.57]** | 0.9 (6/7.0) | [0.4, 1.8] |
| Ethnicity: Not Hispanic or Latino, Race: Asian | 37 | **0.7 (26/37)** | **[0.53, 0.84]** | 0.73 (19/26) | [0.58, 0.91] | 0.91 (10/11) | [0.82, 1.13] | 0.95 (19/20) | [0.9, 1.07] | 1.6 (26/16.1 ) | [1.3, 2.0] |
| Ethnicity: Not Hispanic or Latino, Race: Black or African American | 21 | 0.43 (9/21) | [0.22, 0.66] | 0.67 (6/9) | [0.33, 1.03] | 0.83 (10/12) | [0.67, 1.05] | 0.75 (6/8) | [0.5, 1.1] | 1.5 (9/6.1) | [0.9, 2.4] |
| Ethnicity: Hispanic or Latino, Race: White | 12 | 0.33 (4/12) | [0.1, 0.65] | 0.75 (3/4) | [0.5, 1.5] | 0.88 (7/8) | [0.75, 1.15] | 0.75 (3/4) | [0.5, 1.26] | 1.5 (4/2.7) | [0.7, 3.3] |
| Ethnicity: Not Hispanic or Latino, Race: Native Hawaiian or Other Pacific Islander | 10 | 0.4 (4/10) | [0.12, 0.74] | 0.75 (3/4) | [0.5, 1.25] | 1.0 (6/6) | [1.0, 1.0] | 1.0 (3/3) | [1.0, 1.0] | 1.8 (4/2.3) | [0.8, 3.8] |
| Ethnicity: Not Hispanic or Latino, Race: Other | 10 | 0.5 (5/10) | [0.19, 0.81] | 0.4 (2/5) | [-0.2, 0.8] | 1.0 (5/5) | [1.0, 1.0] | 1.0 (2/2) | [1.0, 1.0] | 1.4 (5/3.5) | [0.8, 2.6] |

**Supplemental Table 19: Stanford HM ACP in Hospital Medicine: Reliability and Fairness Audit by Ethnicity/Race.** Significant differences in prevalence, significantly lower performance, or significantly higher O/E are bolded.

| Group | Sample Size | Prevalence (Fraction) | Prevalence [95% CI] | Sensitivity (Fraction) | Sensitivity [95% CI] | Specificity (Fraction) | Specificity [95% CI] | Positive Predictive Value (Fraction) | Positive Predictive Value [95% CI] | O/E (Fraction) | O/E [95% CI] |
|---|---|---|---|---|---|---|---|---|---|---|---|
| Overall | 225 | 0.44 (99/225) | [0.37, 0.51] | 0.69 (68/99) | [0.6, 0.78] | 0.87 (109/126) | [0.81, 0.93] | 0.8 (68/85) | [0.72, 0.89] | 1.5 (99/65.2 ) | [1.3, 1.8] |
| Ethnicity: Not Hispanic or Latino, Race: White, Sex: Female | 43 | 0.53 (23/43) | [0.38, 0.69] | 0.65 (15/23) | [0.47, 0.85] | 0.85 (17/20) | [0.7, 1.05] | 0.83 (15/18) | [0.67, 1.01] | 1.8 (23/12.5 ) | [1.4, 2.4] |
| Ethnicity: Not Hispanic or Latino, Race: White, Sex: Male | 38 | 0.34 (13/38) | [0.2, 0.51] | 0.69 (9/13) | [0.46, 0.96] | 0.96 (24/25) | [0.92, 1.06] | 0.9 (9/10) | [0.8, 1.13] | 1.7 (13/7.7) | [1.1, 2.6] |
| Ethnicity: Hispanic or Latino, Race: Other, Sex: Female | 22 | 0.18 (4/22) | [0.05, 0.4] | 0.5 (2/4) | [0.0, 1.0] | 0.83 (15/18) | [0.67, 1.0] | 0.4 (2/5) | [-0.2, 0.8] | 0.8 (4/4.8) | [0.3, 2.0] |

| | | | | | | | | | | | |
|---|---|---|---|---|---|---|---|---|---|---|---|
| Ethnicity: Not Hispanic or Latino, Race: Asian, Sex: Male | 21 | **0.81 (17/21)** | **[0.58, 0.95]** | 0.65 (11/17) | [0.42, 0.87] | 0.75 (3/4) | [0.5, 1.25] | 0.92 (11/12) | [0.83, 1.12] | 1.8 (17/9.6) | [1.4, 2.2] |
| Ethnicity: Hispanic or Latino, Race: Other, Sex: Male | 16 | 0.12 (2/16) | [0.02, 0.38] | **0.0 (0/2)** | **[0.0, 0.0]** | 0.86 (12/14) | [0.71, 1.05] | **0.0 (0/2)** | **[0.0, 0.0]** | 0.9 (2/2.2) | [0.3, 3.4] |
| Ethnicity: Not Hispanic or Latino, Race: Asian, Sex: Female | 16 | 0.56 (9/16) | [0.3, 0.8] | 0.89 (8/9) | [0.78, 1.14] | 1.0 (7/7) | [1.0, 1.0] | 1.0 (8/8) | [1.0, 1.0] | 1.4 (9/6.4) | [0.9, 2.2] |
| Ethnicity: Not Hispanic or Latino, Race: Black or African American, Sex: Female | 12 | 0.67 (8/12) | [0.35, 0.9] | 0.75 (6/8) | [0.5, 1.07] | 0.75 (3/4) | [0.5, 1.3] | 0.86 (6/7) | [0.71, 1.14] | 1.7 (8/4.8) | [1.1, 2.5] |
| Ethnicity: Not Hispanic or Latino, Race: Black or African American, Sex: Male | 9 | 0.11 (1/9) | [0.0, 0.48] | **0.0 (0/1)** | **[0.0, 0.0]** | 0.88 (7/8) | [0.75, 1.12] | **0.0 (0/1)** | **[0.0, 0.0]** | 0.8 (1/1.3) | [0.1, 5.0] |
| Ethnicity: Hispanic or Latino, Race: White, Sex: Male | 7 | 0.29 (2/7) | [0.04, 0.71] | 0.5 (1/2) | [0.0, 1.0] | 1.0 (5/5) | [1.0, 1.0] | 1.0 (1/1) | [1.0, 1.0] | 2.0 (2/1.0) | [0.6, 6.3] |
| Ethnicity: Not Hispanic or Latino, Race: Native Hawaiian or Other Pacific Islander, Sex: Female | 6 | 0.17 (1/6) | [0.0, 0.64] | **0.0 (0/1)** | **[0.0, 0.0]** | 1.0 (5/5) | [1.0, 1.0] | nan (0/0) | N/A | 0.8 (1/1.2) | [0.1, 5.0] |
| Ethnicity: Not Hispanic or Latino, Race: Other, Sex: Male | 6 | 0.5 (3/6) | [0.12, 0.88] | **0.0 (0/3)** | **[0.0, 0.0]** | 1.0 (3/3) | [1.0, 1.0] | nan (0/0) | N/A | 2.3 (3/1.3) | [1.0, 5.0] |
| Ethnicity: Hispanic or Latino, Race: White, Sex: Female | 5 | 0.4 (2/5) | [0.05, 0.85] | 1.0 (2/2) | [1.0, 1.0] | 0.67 (2/3) | [0.33, 1.33] | 0.67 (2/3) | [0.33, 1.33] | 1.2 (2/1.7) | [0.4, 3.4] |
| Ethnicity: Not Hispanic or Latino, Race: Native Hawaiian or Other Pacific Islander, Sex: Male | 4 | 0.75 (3/4) | [0.19, 0.99] | 1.0 (3/3) | [1.0, 1.0] | 1.0 (1/1) | [1.0, 1.0] | 1.0 (3/3) | [1.0, 1.0] | 2.9 (3/1.1) | [1.6, 5.0] |
| Ethnicity: Not Hispanic or Latino, Race: Other, Sex: Female | 4 | 0.5 (2/4) | [0.07, 0.93] | 1.0 (2/2) | [1.0, 1.0] | 1.0 (2/2) | [1.0, 1.0] | 1.0 (2/2) | [1.0, 1.0] | 0.9 (2/2.2) | [0.3, 2.4] |

**Supplemental Table 20: Stanford HM ACP in Hospital Medicine: Reliability and Fairness Audit by Ethnicity/Race and Sex.** Significant differences in prevalence, significantly lower performance, or significantly higher O/E are bolded.

*Clinical Decision Maker Survey Responses*

| Theme | Example Response | Response Count |
|---|---|---|
| Primary Care: Excitement | "I was impressed that the PPV and specificity were so high. Very encouraging and exciting! I think it will work well for our intended purpose!" | 2 |
| Primary Care: TrustToUseForPurpose | "The results are really exciting! They make sense clinically and I appreciate that in the context of implementation, I would be able to trust the flag as being accurate/helpful at guiding my decision to have more focused, in-depth ACP discussions with the patient and their family/caregivers. " | 2 |
| Inpatient Oncology: Low Sample Size | "Number too small " | 2 |
| Inpatient Oncology: Results Depend On Threshold | "Reliability depends on the model threshold picked, There may be some signals of differences based on age and race/ethnicity groups, but I wonder if this is in part limited by low power." | 2 |
| Inpatient Oncology: EOL Underpredicted Death Risk | "Epic end of life index underpredicted death risk for cancer patients (~30 out of 150 medonc inpatients being flagged at high risk of death is way too low!). ACP model results looked pretty good. Comparison to physicians' predictions and not actual death data limits conclusions since we don't know how good the clinicians' performance is, whether their results are biased for certain ethnic groups etc." | 1 |
| Inpatient Oncology: Reassured that Reliable/Fair | "It’s reassuring that there are these safeguards to ensure that models are reliable and fair. " | 1 |

| | | |
|---|---|---|
| Safeguards Exist | | |
| Inpatient Oncology: Low Sample Size: Affects Detecting Differences By Race | "Reliability depends on the model threshold picked, There may be some signals of differences based on age and race/ethnicity groups, but I wonder if this is in part limited by low power." | 1 |
| Inpatient Oncology: ACP Not Specific At Threshold | "It was not surprising that the ACP model was not specific and made me wonder if we might adjust that threshold " | 1 |
| Hospital Medicine: Interesting | "Interesting!" | 3 |
| Hospital Medicine: Interesting: Clinician Difference By Race | "I found it was interesting how the model and clinicians agreed at the extremes of populations. It was also interesting to see that as a clinician, our predictions are different across races (ie. LatinX more likely to be surprised and Asians less likely). " | 1 |
| Hospital Medicine: Interesting: Sensitivity Difference By Race | "interesting that there was a difference in sensitivity by race " | 1 |

| | | |
|---|---|---|
| Hospital Medicine: Interesting: Models and Clinicians Agree at Extremes Of Populations | "I found it was interesting how the model and clinicians agreed at the extremes of populations. It was also interesting to see that as a clinician, our predictions are different across races (ie. LatinX more likely to be surprised and Asians less likely). " | 1 |

**Supplemental Table 21: Survey responses to "What are the first thoughts that came to your mind on seeing the results of the reliability and fairness audit?"**

| Theme | Example Response | Response Count |
|---|---|---|
| More reliable race data in EHR | "I wish Epic had more reliable race data" | 2 |
| Link subgroup analysis with population demographics ("Model Performance for subgroup X (X's % of patient population") | "I would like to know how the subgroup analysis - particularly the race/ethnicity analyses - relate to our population demographics as a whole. For example, the model is less likely to identify X type of patients, these patients make up X% of your patient population. " | 1 |
| Model's ranking of important patients | "Some statistic to describe rank order" | 1 |
| Broader audit in collaboration with other centers | "Broader effort with other centers " | 1 |

| | | |
|---|---|---|
| Subgroup analysis by clinical characteristic (cancer type, performance status, etc.) | "Performance for patients with varying clinical characteristics (cancer type, performance status, etc.). Performance for patients newer to Stanford system or more established." | 1 |
| Subgroup analysis by patient's time with health system (newer or more established) | "Performance for patients with varying clinical characteristics (cancer type, performance status, etc.). Performance for patients newer to Stanford system or more established." | 1 |

**Supplemental Table 22: Survey responses to "Is there any other information you would want included in this audit to support your decision on whether to deploy a model? If so, what?"**

| **Drivers to make these reliability and fairness audits standard practice** | **Responses** |
|---|---|
| Findings that AI models are not fair | 10 |
| Findings that AI models are not reliable | 9 |
| Academic medicine's push toward racial equity | 9 |

**Supplemental Table 23: Survey responses to “What are some key drivers to making these reliability and fairness audits standard practice?”**

| **Barriers to make these reliability and fairness audits standard practice** | **Responses** |
|---|---|
| Poor demographic data quality | 8 |
| Poor data quality | 6 |
| Lack of data access | 5 |
| Audits are not built into our incentives | 4 |
| Lack of knowledge about how to do an audit | 3 |
| The reliability of deployed AI models is not prioritized | 3 |
| The fairness of deployed AI models is not prioritized | 3 |
| Lack of data science expertise in my practice setting | 2 |
| Other: I don't see us as designing them, but if teams want to engage providers in helping with these audits, I think the most significant barrier is the time, but if there is incentive/appreciation/protected time to do the audit, I can't think of any other barriers | 1 |
| Other: Death data | 1 |

| | |
|---|---|
| I do not see any barriers to making reliability and fairness audits standard practice. | 0 |

**Supplemental Table 24: Survey responses to "What are some key barriers to making these reliability and fairness audits standard practice?"**

| **Pros in using AI to support my work** | **Responses** |
|---|---|
| Helps triage patients and identify who would benefit the most | 10 |
| Shared understanding of patients for our whole care team | 9 |
| Reduces work for me | 3 |
| I do not see any pros to using an AI model to support my work. | 0 |

**Supplemental Table 25: Survey responses to "As a clinical decisionmaker, what pros do you see in using an AI model to support your work?"**

| **Cons in using AI to support my work** | **Responses** |
|---|---|
| Lack of transparency of the model | 5 |
| Takes effort to maintain | 4 |
| I disagree with the model | 3 |
| Loss of my decisionmaking autonomy | 2 |
| Pressure to act even if I disagree with the model | 1 |

| | |
|---|---|
| Other: Worry that the model may miss some patients who might benefit | 1 |
| Other: The HM model although is more sensitive- so many patients flag. Is it possible to risk stratify who is highest risk (using green, yellow, red) like the Epic AI models | 0 |
| I do not see any cons to using an AI model to support my work. | 1 |

**Supplemental Table 26: Survey responses to "As a clinical decisionmaker, what cons do you see in using an AI model to support your work?"**
